# Levothyroxine treatment during pregnancy: a metabolomics study

**DOI:** 10.1101/2025.04.04.25325273

**Authors:** Olli Kärkkäinen, Heidi Sahlman, Leea Keski-Nisula, Jaana Rysä

**Affiliations:** School of Pharmacy, University of Eastern Finland, Finland; Institute of Clinical Medicine, School of Medicine, University of Eastern Finland, Finland; Department of Obstetrics and Gynecology, Kuopio University Hospital, Finland

**Keywords:** metabolomics, NMR, pregnancy, levothyroxine, hypothyroidism

## Abstract

**Background:** While levothyroxine is one of the most extensively prescribed drugs during pregnancy, the possible effects of levothyroxine on the metabolome are not well known. Our aim was to determine levothyroxine treatment-associated changes in the metabolite profile of umbilical cord serum after birth, as well as in maternal serum samples collected at different stages of pregnancy, and link these to the health of the newborn.

**Methods:** The study cohort, 118 levothyroxine-treated and 118 healthy control pregnancies, was collected from Kuopio University Hospital, Finland, during the years 2013–2017. Serum metabolite profiles were determined with nuclear magnetic resonance-based metabolomics from 1) umbilical cord blood samples, 2) samples collected during the 1^st^ trimester and 3) during delivery from the pregnant women. There was no difference in demographic characteristics between study groups including gestational age.

**Results:** There was a negative correlation between cord blood thyroid stimulating hormone (CBTSH) concentrations and Apgar scores at the 1-minute and 5-minute time-points in levothyroxine-treated pregnancies. Furthermore, the concentrations of cord serum metabolites linked with anaerobic glycolysis, e.g., lactate, citrate and glycerol, as well as all measured amino acids were negatively associated with Apgar scores. Furthermore, cord serum concentrations of lactate (β = 0.50, p < 0.0001), glycerol (β = 0.41, p < 0.0001) and alanine (β = 0.34, p = 0.0005) were positively correlated with CBTSH concentrations in the levothyroxine-treated pregnancies. No differences in the 1^st^ trimester samples were observed between the groups. In the during delivery samples, there was small but significant decrease in cholesteryl esters, cholesterol and phospholipids in small very low-density lipoprotein in the levothyroxine-treated pregnancies.

**Conclusions:** In the levothyroxine-treated pregnancies, the alterations detected in the cord serum concentrations of metabolites linked to fetal hypoxia and muscle degradation could explain the association between CBTSH and the health of the newborn measured via Apgar scores.

## Introduction

Hypothyroidism, i.e. an insufficiency of circulating thyroid hormones, is a common condition with a prevalence of 5% in the population. During pregnancy, overt hypothyroidism (OH) and subclinical hypothyroidism (SCH) are diagnosed in approximately 0.4 to 0.9% (OH) and 3 to 8.5% (SCH) of pregnant women, but it is believed that these conditions are more common than generally acknowledged ^1–3^. Approximately 2% to 15% of fertile aged women are consuming levothyroxine (LT4) which is the most commonly used medication to treat hypothyroidism and also one of the drugs most frequently prescribed during pregnancy ^4–6^. Women diagnosed with hypothyroidism are invariably treated with LT4 and women using LT4 prior pregnancy will need a 20% to 50% increase in their doses at the onset of pregnancy ^7^. LT4 replacement therapy is considered a safe and effective treatment to minimize maternal and fetal harm.

Adequate levels of thyroid hormones are essential for both the maintenance of pregnancy as well as fetal development ^8^. In order to meet the increased physiological demands of the growing fetus, the maternal production of thyroid hormones is augmented. The fetus is completely dependent on maternal thyroid hormone supply until the onset of fetal thyroid function in mid-gestation but a significant transfer of thyroid hormones from the mother to the fetus still occurs during the second half of gestation ^8^. Hypothyroidism during pregnancy has been associated with pregnancy complications such as increased risks of premature birth, low birth weight, pregnancy loss as well as gestational diabetes and pre-eclampsia ^4,5,9,10^. In addition to the adverse effects during pregnancy, hypothyroidism may also have long-term consequences for the developing fetus since adequate levels of thyroid hormones are needed for fetal brain development ^8^.

Metabolomics is a tool that can be used to study various biological processes, including pregnancy and its complications ^11^. Metabolomics involves the comprehensive analysis of the small molecules (metabolites) present in biological fluids or tissues, such as blood. Metabolomics can provide insights into the biochemical pathways and networks that underlie various physiological and pathological processes that occur during pregnancy. Previous studies analyzing maternal and cord blood samples have shown that the maternal metabolite profile is altered not only as gestation progresses, but also in maternal pregnancy disorders as well as changes due to chemical exposures during pregnancy ^12–16^. Even though LT4 is widely used during pregnancy, little is known about its impact on the metabolome during pregnancy. A case-control study on gestational hypothyroidism has suggested that hypothyroidism is associated with increased cord blood concentrations of creatinine and O-phosphocholine, and decreased cord blood levels of citric acid, tyrosine, and carnitine ^17^. However, as far as we are aware, no longitudinal metabolomics studies have been published on the effects of LT4 treatment during pregnancy on the circulating metabolome of pregnant women or on umbilical cord blood metabolite profiles.

Consequently, the aim of this study was to evaluate whether LT4 treatment during pregnancy is associated with changes in metabolome. Our hypothesis was that LT4 treatment would be associated with altered lipid and energy metabolism. In this study, we utilized nuclear magnetic resonance (NMR) based metabolite profiling to determine whether the maternal serum metabolite profile differs at different stages of pregnancy, or in cord serum, in response to LT4 prescribed for the treatment of hypothyroidism.

## Patients and methods

### Subjects

The study cohort was collected from the Kuopio Birth Cohort (Kubico) ^18^ from mother –child pairs born in Kuopio University Hospital (KUH) during the years 2013–2017. All pregnant women who were expected to give birth in KUH in the Finnish county Northern Savo were invited to participate in KuBiCo. Participation in the study was voluntary and the women gave informed written consent at the time of recruitment. The study was approved by the Research Ethics Committee of Hospital District of Central Finland in Jyväskylä, Finland 15.11.2011 (18U/2011).

The following demographic and clinical information were collected from the KUH Birth Register from each woman in the KuBiCo trial along with details of her ongoing pregnancy (maternal age, weight and height, parity, gravidity, medication in hospital), maternal diseases, and the delivery (mode and gestational age at delivery, birth weight, Apgar scores, fetal sex, and number of fetuses, cord blood TSH value (CBTSH)). The self-reporting part of the register contained also information about maternal use of over-the-counter and prescription drugs, as well as information about smoking and dietary habits. Information about the most common pregnancy complications, gestational diabetes (GDM) and preeclampsia, was also collected. The diagnostic code (ICD-10) used for GDM was O24.4; for preeclampsia, the ICD-10 codes were mild to moderate preeclampsia (O13 and O14.0), severe preeclampsia (O14.1) or unspecified preeclampsia (O14.9) or eclamptic seizures (O15.0, O15.1, O15.2 and O15.9).

Study population consisted of women treated with LT4 medication in the KuBiCo cohort. We included all LT4 treated Kubico women who were pregnant with one child and had provided a blood sample. A total of 118 women participating in KuBiCo were using LT4 during pregnancy and thus 118 control women not using LT4 were included in the study. Controls were randomly selected and not specifically matched to LT4 treated women. Inclusion criteria for controls were singleton pregnancies and not using LT4 medication. The following diagnostic criteria were used for hypothyroidism; TSH value > 4.2 mU/l and T4 value < 11 pmol/l ^19,20^. For subclinical hypothyroidism, the diagnostic criteria were TSH value > 2.5 mU/l in the first trimester and TSH value > 3 mU/l in the second and third trimester and normal T4 value ^5^. Pregnant women diagnosed with overt or subclinical hypothyroidism were treated with LT4. Information about the LT4 treatment was collected from the KUH birth register. Information about the duration of LT4 dosing was available in 95.8% of women (113/118); 74.3% of these women received LT4 medication during either throughout pregnancy (50.4%) or had started medication during first trimester (23.9 %), 19.5% had started medication during second and 6.2% during third trimester. Of the women treated with LT4, dose was reported from 81.4% of users. Dose varied between 25 µg and 700 µg per day, 100 µg being the most common dose. 89.2% of LT4 users used dose 50 to 200 µg/day. One woman had subclinical hypothyroidism and she was treated with 12.5 µg levothyroxine for 3 times / week. Information about the serum levels of TSH and T4 during pregnancy were obtained from hospital’s medical records.

### Blood sampling

All women had a venous blood sample drawn by professional laboratory staff during the 1^st^ trimester. Maternal venous blood sample collected during the delivery and an umbilical cord vein blood sample were taken by midwives. Blood samples were centrifuged by the laboratory staff and serum was collected. Samples were stored at −80 °C until analysis. CBTSH levels and pregnant women’s TSH and LT4 concentrations were measured with accredited electrochemiluminescence immunoassay (ECLIA) methods on a Roche Cobas® e 601 automated analyzer (Roche Diagnostics) by Eastern Finland Laboratory Centre Joint Authority Enterprise (ISLAB) (a laboratory center for the Kuopio University Hospital). Free thyroxine was measured using FT4 kit (Roche Cat# 11731297, RRID:AB_2936841) or Elecsys FT4 II kit (Roche Cat# 06437281, RRID:AB_2924686) and TSH was measured using Elecsys® TSH Kit (Roche Cat# 11731459, RRID:AB_2756377).

### NMR Metabolite profiling analysis

Maternal serum samples collected during the 1^st^ trimester (between weeks 9 to 12) and during delivery as well as umbilical cord samples were used as a matrix for metabolite profiling analysis. A total of 216 serum samples were taken during the 1^st^ trimester (98 cases, 118 controls), 236 serum samples taken during delivery (118 cases, 118 controls) and 236 samples from umbilical cord serum (118 cases, 118 controls).

The NMR based metabolite profiling method (Nightingale Health Plc. Helsinki, Finland), which has been described in detail earlier ^21,22^, was used to measure metabolites from the samples. The analysis platform consisted of individual metabolites as well as ratios between different measurements, representing a total of 250 variables.

### Statistical analysis

Pearson’s Chi-squared test and Welch’s t-test were used to compare differences in the background variables between the groups. The influence of confounding factors (Apgar scores, maternal age, BMI, parity and smoking status) to newborn CBTSH values was analyzed by linear regression analysis.

Principal component analysis (PCA) was used to analyze the overall variance in the data. Because variables from the metabolite profiling analysis correlate with each other, the number of components needed to explain 95% of variance in the metabolomics data (here 17 components) was used to correct the α level to account for multiple tests and thus the new α was set to 0.003. Variables with p-values between 0.05 and 0.003 were considered to be trends.

Paired samples t-test and Cohen’s d effect sizes were used to investigate metabolites that had changed during pregnancy (1^st^ trimester vs. at delivery samples) and between mothers’ samples and the cord serum samples. Furthermore, Welch’s t-test and Cohen’s d effect sizes were used to investigate which metabolites were altered in each sample type separately. Moreover, repeated measures ANOVA was used to investigate the effect of LT4 treatment on the change seen in the metabolite profile between 1^st^ trimester samples and specimens collected during delivery. Linear regression was used to calculate correlations between metabolite levels and background variables for each study group separately. Significance of the difference between correlation coefficients of the two groups were assessed with the Fisher’s z-transformation of correlations test.

## Results

There was no difference in background characteristics between study groups including gestational age. In the case-control comparison between LT4 treatment, there were no significant differences in the metabolomics variables between the study groups at 1^st^ trimester samples nor in the cord serum samples (Figure 1). However, we observed significant differences in the metabolomics variables between the study groups in the samples collected during delivery (Figure 1, Supplementary Table 1). After correction for multiple testing, cholesteryl esters in small very low-density lipoprotein VLDL (p = 0.0016, d = −0.42), cholesterol in small VLDL (p = 0.0025, d = −0.40), free cholesterol in very small VLDL (p = 0.0027, d = −0.39), and phospholipids in very small VLDL (p = 0.0030, d = −0.39) were significantly decreased in the samples of LT4-treated women during delivery when compared to the samples of controls. Furthermore, many measured variables related to small or very small VLDL particles had p-values between the multiple testing corrected α level (0.003) and 0.05, indicating that the lipid content of the small and very small VLDL particles did differ between the study groups (Figure 1).

**Figure 1:**
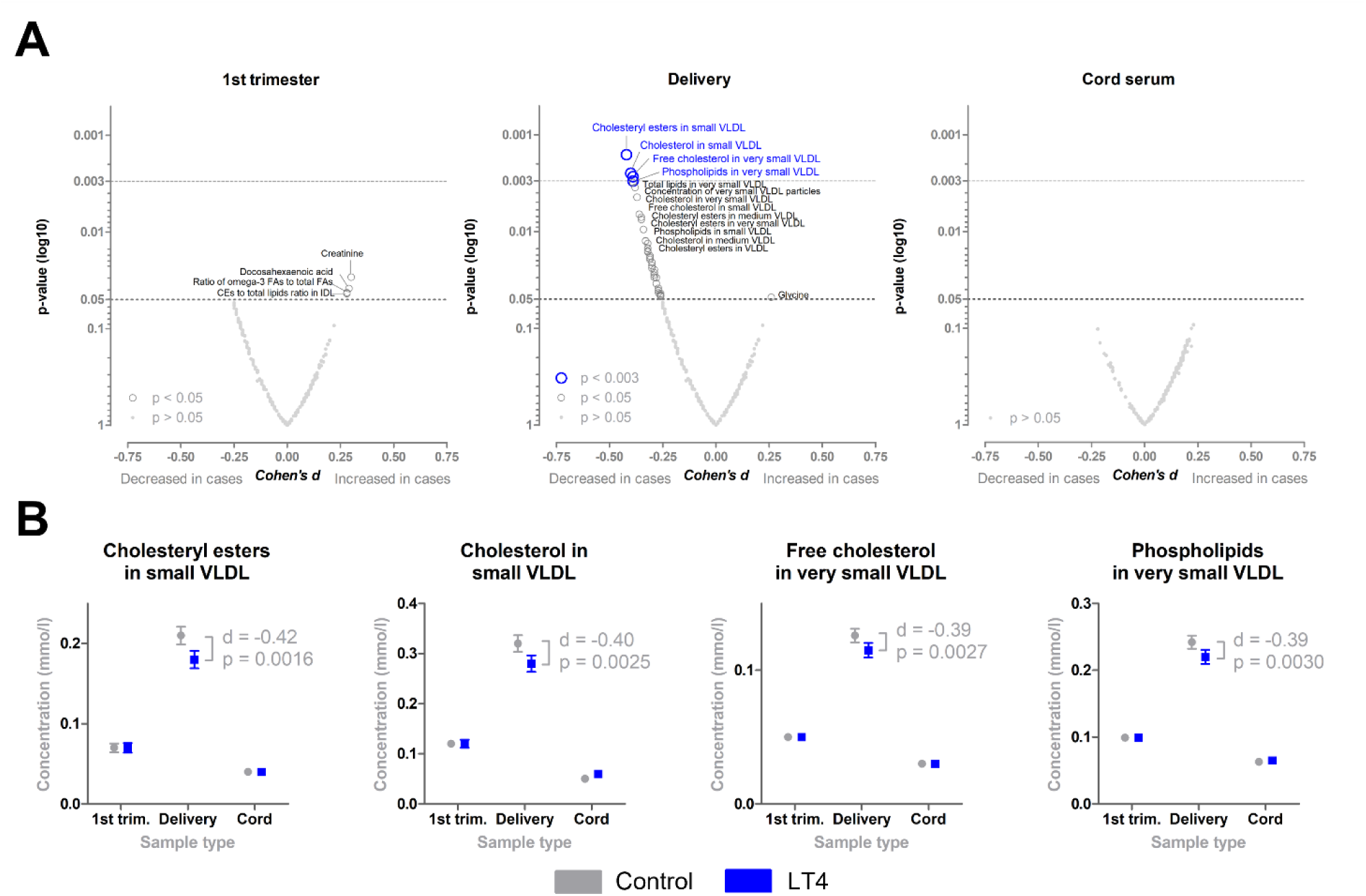
Levothyroxine exposure during pregnancy is associated with an altered serum metabolite profile in samples collected during delivery from the mother, but not in samples collected during the 1^st^ trimester of pregnancy nor in cord serum samples. Volcano plots showing Cohen’s d effect sizes and p-values (in log10 format) from Welch’s t-test (A). Cholesteryl esters in small very low-density lipoprotein (VLDL), cholesterol in small VLDL, free cholesterol in very small VLDL, and phospholipids in very small VLDL were significantly decreased in the levothyroxine-exposed pregnancies when compared to the controls in the samples collected during delivery from the mothers (B). Variables were considered to be significantly altered when the p-value was below the multiple testing corrected α level (0.003). Variables with p-values between 0.05 and 0.003 were considered to be trends.

There were significant differences in the metabolite profiles between the sample types in the PCA (Figure S1, Table S1). The metabolite profile of the serum samples from pregnant women changed dramatically during pregnancy, with almost all measured variables displaying a significant difference between the sample types (Figure 2). Similarly, there was a large difference between the serum samples taken during delivery from the women and cord serum samples, with most variables revealing significant differences. When comparing samples collected during delivery to samples collected during the 1^st^ trimester, there were 234 significantly (p < 0.003, multiple testing corrected α level, paired samples t-test) altered variables. There were 49 significantly decreased variables (Figure S2), and 185 significantly increased variables (Figure S3) in the serum samples taken during delivery, when compared to samples collected during the 1^st^ trimester. This indicates that the serum metabolite profile of the pregnant woman undergoes major changes during pregnancy (Figure 2).

**Figure 2:**
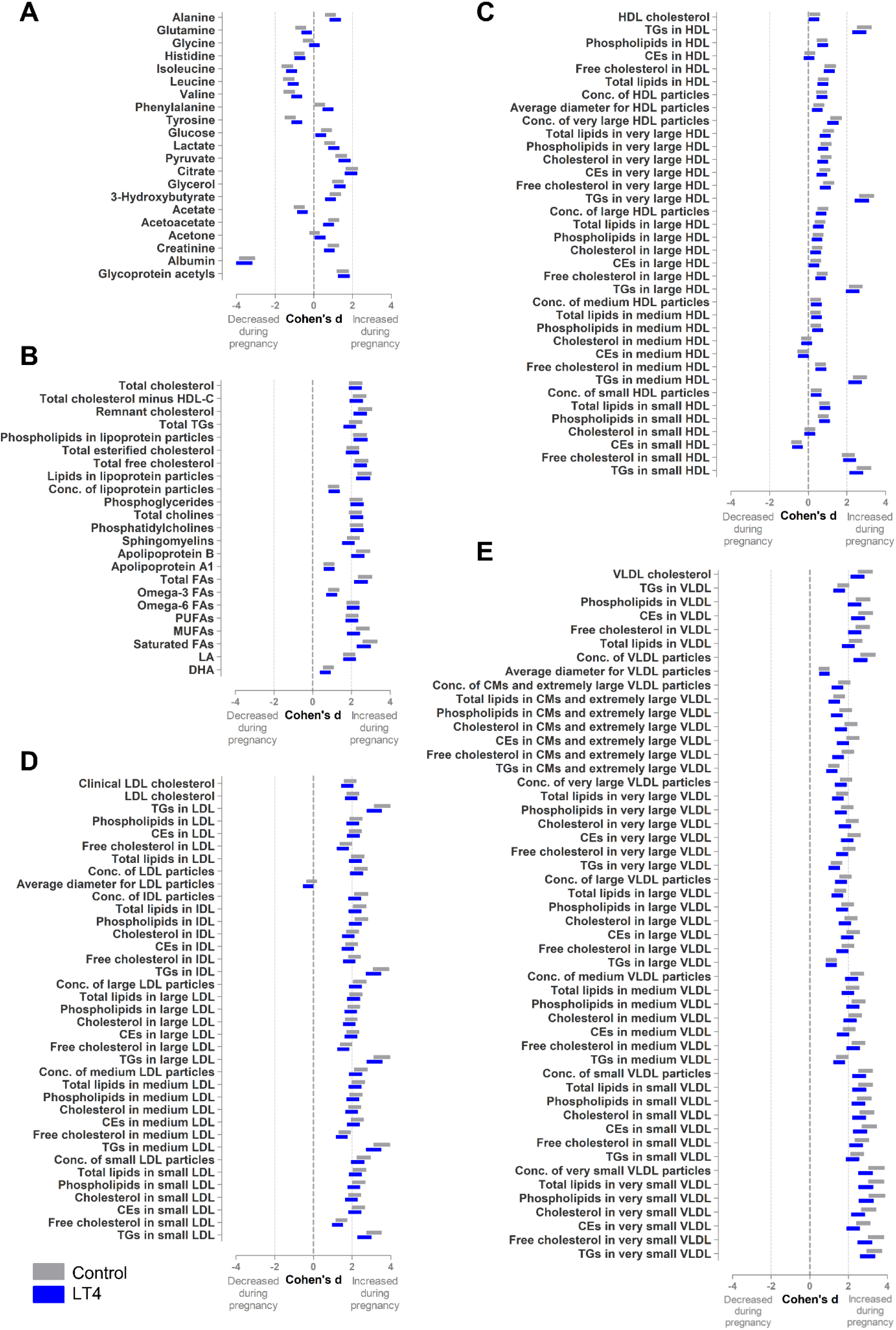
Pregnancy alters concentrations of most of the measured metabolites. When comparing samples collected during 1^st^ trimester of pregnancy and samples collected during delivery, the metabolite profile of the samples had changed dramatically. Floating bars show 95% confidence interval for Cohen’s d value. For most variables, there were no significant differences in the level of change during pregnancy between the study groups. However, there was a significant difference in the level of change during pregnancy between the levothyroxine (LT4) treated pregnancies and controls in the concentration of cholesteryl esters in small VLDL (p = 0.0022, Cohen’s d for difference in change during pregnancy = −0.42). Legend: CE, cholesteryl ester; CM, chylomicron; DHA, docosahexaenoic acid; FA, fatty acid; HDL, high-density lipoprotein; LA, linoleic acid; MUFA, monounsaturated fatty acid; PUFA, polyunsaturated fatty acid; LDL, low-density lipoprotein; VLDL, very low-density lipoprotein; TG, triglyceride.

There were no significant differences in the background variables between the study groups, with the exception of the higher CBTSH concentrations in the newborns born to women treated with LT4 (mean 14.94 mU/L, n=118) as compared to newborn CBTSH concentrations in controls (mean 11.54 mU/L, n=118) (Table 1, Figure 3). Furthermore, the CBTSH concentration correlated negatively with the Apgar scores evaluated at both the 1- and 5-minutes time-points (standardized β = −0.28, p = 0.0032; and β = −0.27, p = 0.0044, respectively) (Figure 3) in the LT4 group, but not in the control pregnancies (standardized β = −0.11, p = 0.2718; and β = −0.07, p = 0.4140, respectively). These negative correlations remained significant when background characteristics of the pregnant women (age, BMI, parity and smoking status) were included in the linear models (β = - 0.29, p = 0.0032; and β = −0.26, p = 0.0087, respectively). However, the difference in the correlation coefficients was not significantly different between the study groups (z = 1.38, p = 0.1676; and z = 1.51, p = 0.1310, respectively). There were no significant correlations between CBTSH concentrations and birth weight of the child (cases β = 0.16, p = 0.0872; controls (β = − 0.09, p = 0.3375).

**Figure 3:**
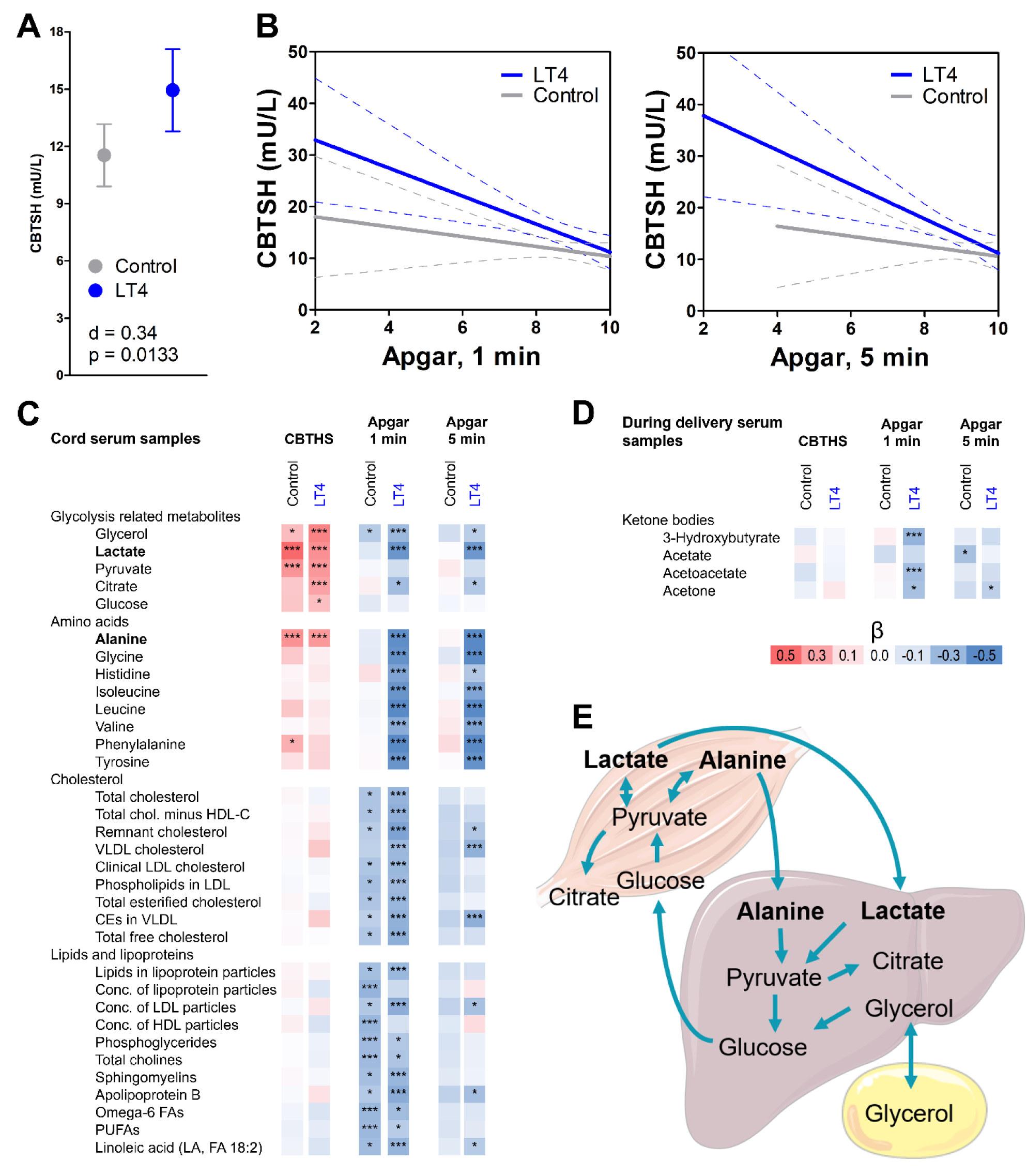
Cord blood concentration of thyroid stimulating hormone correlates negatively with Apgar score in levothyroxine treated pregnancies. Levothyroxine (LT4) treated pregnancies had higher cord blood concentrations of thyroid stimulating hormone (CBTSH) when compared to the controls (A, Cohen’s d = 0.34, p = 0.0133, group means and 95% confidence intervals are shown). CBTSH concentrations correlated negatively with Apgar scores measured at both 1 minute and 5 minutes after birth in the LT4 group (standardized β = −0.28, p = 0.0032, and β = −0.27, p = 0.0044, respectively), but not in the controls (β = −0.10, p = 0.2718, and β = −0.08, p = 0.4140, respectively) (B). When the background characteristics of the pregnant women (age, body mass index, parity, and smoking status) were included in the linear models, the negative correlation between CBTSH and Apgar scores remained statistically significant in the LT4 group (1 min: β = −0.29, p = 0.0032; and 5 min: β = −0.26, p = 0.0087). In the LT4 group, significant (p < 0.003, ***) positive correlations were observed between CBTSH and glycerol, lactate, pyruvate, citrate and alanine (C, standardized β values are shown). Furthermore, in the LT4 group, the cord serum levels of glycerol, lactate and alanine, as well as most of the other measured amino acids, correlated negatively with Apgar scores, a phenomenon which was not observed in the controls. Furthermore, in both study groups, there were significant negative correlations between Apgar scores at 1 minute and cord serum concentrations of cholesterol, lipids, and lipoproteins. In the serum samples collected from the mothers during delivery, ketones 3-hydroxybutyrate and acetoacetate correlated negatively with Apgar scores at 1 minute in the LT4 treated pregnancies, but not in the control pregnancies (D). The positive correlation between TSH and cord serum lactate (Cori’s cycle, lactic acid cycle) indicates that anaerobic glycolysis is increased in the muscles (E). High cord serum alanine concentrations correlating with high concentration of TSH are evidence of muscle protein being degraded to generate more pyruvate and glucose (in the glucose-alanine cycle, the Cahill cycle). The fact that both lactate and alanine display a significant negative correlation to Apgar scores in the LT4 exposed children could indicate the possible presence of hypoxia and muscle degradation associated with CBTSH. *, p = 0.05 – 0.003 (trend); ***, p < 0.003 (significant at the multiple testing corrected α level).

**Table 1:** Background characteristics of the pregnant women and their newborns.

| Variable | Controls<br>(n = 118) | Levothyroxine users<br>(n = 118) | p-value |
| --- | --- | --- | --- |
| **Maternal characteristics** |  |  |  |
| Age (years, mean, ±SD) | 31.2 ±5.3 | 31.0 ±5.4 | 0.7859<sup>a</sup> |
| BMI before pregnancy (kg/m<sup>2</sup>, mean ±SD) | 25.8 ±5.7 | 26.0 ±6.2 | 0.8261<sup>a</sup> |
| Self-reported tobacco smoking (n, %) |  |  | 0.9141<sup>b</sup> |
| not smoking | 92 (78%) | 94 (79.7%) |  |
| quit smoking before pregnancy | 3 (2.5%) | 2 (1.7%) |  |
| continued smoking during pregnancy | 21 (17.8%) | 19 (16.1%) |  |
| smoking status unknown | 2 (1.7%) | 3 (2.5%) |  |
| Parity (mean ±SD) | 0.9 ±1.0 | 0.7 ±1.0 | 0.0731<sup>a</sup> |
| Gestational weeks (GWs) | 39.6 ±1.5 | 39.1 ±2.1 | 0.732<sup>b</sup> |
| GWs (excluding caesarean sections)<sup>1</sup> | 39.7 ±1.3 | 39.5 ±1.4 | |
| Gestational diabetes mellitus | 28 (24%) | 17 (14%) | 0.0683<sup>b</sup> |
| Caesarean section (number, %) | 10 (9%) | 16 (14%) | 0.2581<sup>b</sup> |
| Preeclampsia | 3 (2.5%) | 1 (0.8%) | 0.3132<sup>b</sup> |
| TSH value mU/L (mean±SD)<sup>2</sup> | NA | 3.4 ±2.9 | |
| T4 value pmol/L (mean±SD)<sup>3</sup> | NA | 13.1 ±2.0 | |
| **Newborn characteristics** |  |  |  |
| Birth weight (grams, mean ±SD) | 3531 ±471 | 3454 ±547 | 0.2542<sup>a</sup> |
| Head circumference (cm, mean ±SD) | 35.30 ±1.46 | 35.13 ±1.57 | 0.3985<sup>a</sup> |
| Placental weight (grams, mean ±SD) | 578.5 ±113.1 | 578.1 ±113.3 | 0.9802<sup>a</sup> |
| Sex of the child (female / male) | 52 / 64 | 46 / 68 | 0.4924<sup>b</sup> |
| Apgar scores, 1 min (mean ±SD) | 8.8 ±0.9 | 8.6 ±1.2 | 0.3017<sup>a</sup> |
| Apgar scores, 5 min (mean ±SD) | 9.0 ±0.7 | 8.9 ±0.9 | 0.2273<sup>a</sup> |
| Cord blood TSH (mU/L, mean ±SD) | 11.54 ±8.67 | 14.94 ±11.35 | 0.0133<sup>a</sup> |

<sup>a</sup>, Welch's t-test; <sup>b</sup>, Chi-squared test; n, number; SD, standard deviation; TSH, thyroid stimulating hormone, Apgar = Appearance, Pulse, Grimace, Activity, and Respiration score. <sup>1</sup>n=100 (controls) and n= 99 (cases) <sup>2</sup>n=100 (controls) and n= 99 (cases) <sup>3</sup>n=116, <sup>3</sup>n=115, NA= not available

We performed correlation analyses to investigate if this negative correlation between CBTSH values in newborns born to LT4 treated women and Apgar scores could be linked to altered metabolic processes. In the cord serum samples, the metabolites associated with glycolysis were positively correlated with CBTSH concentrations in both controls and LT4 treated pregnancies (Figure 3). Of these, cord serum concentrations of lactic acid, alanine and citrate were also significantly negatively associated with Apgar scores (at 1 and 5 minutes) in the LT4 treated pregnancies, but not in the control pregnancies. There were significant differences between the groups in correlation coefficients (Apgar 1 min *vs.* lactic acid z = 2.90, p = 0.0037; Apgar 1 min *vs.* alanine z = 3.43, p = 0.0006; and Apgar 1 min *vs.* citrate z = 2.56, p = 0.0105; and Apgar 5 min *vs.* lactic acid z = 3.58, p = 0.0003; Apgar 5 min *vs*. alanine z = 4.73, p < 0.0001; and Apgar 5 min *vs*. citrate z = 1.93, p = 0.0536). Furthermore, cord serum concentrations of most of the measured amino acids correlated negatively with Apgar scores (both 1 and 5 minutes) only in the LT4 treated pregnancies (Figure 3). In both study groups, metabolomics variables related to cholesterol, lipids and lipoproteins correlated negatively with Apgar scores at 1 minute after birth (Figure 1). All correlations between metabolite variables measured from umbilical cord serum samples and variables from the newborn are shown in Table S2.

As a post-hoc analysis, we investigated the relative contributions of the key metabolites (metabolites that had significant correlations to both CBTSH and Apgar scores, i.e., lactate and alanine) in mediating the correlation between CBTSH and Apgar score in the LT4 treated pregnancies. We used linear regressions models, where the Apgar score at 1 min or 5 min was the dependent variable, and CBTSH and lactate or alanine were covariates. In the LT4 treated pregnancies, we found 0.22 mmol/L and 0.18 mmol/L reduction in cord serum concentration of lactate (β = −0.40, p < 0.0001, and β = −0.40, p < 0.0001) and 4.7 mmol/L and 4.1 mmol/L reduction in cord serum concentrations of alanine (β = −0.48, p < 0.0001, and β = −0.53, p < 0.0001) for every 1-unit reduction in Apgar score at 1 min and 5 min after birth, respectively, when CBTSH was also a covariate (β = −0.13, p = 0.1735; β = −0.12, p = 0.2143; β = −0.12, p = 0.2112; and β = −0.09, p = 0.3176, respectively).

Furthermore, also as a post-hoc analysis, we removed samples from pregnancy conditions known to affect the metabolome, i.e. pre-eclampsia ^12^, GDM ^23^, from the regression analyses (total of 18 cases removed). The key correlations remained significant between CBTSH levels and cord serum concentrations of lactate (β = 0.43, p < 0.0001) and alanine (β = 0.29, p = 0.0076) in the LT4 treated pregnancies. Moreover, also the negative correlations between Apgar scores at 1 min and 5 min and cord serum concentrations of lactate (β = −0.46, p < 0.0001; β = −0.47, p < 0.0001, respectively) and alanine (β = −0.48, p < 0.0001; β = −0.55, p < 0.0001) remained significant in the LT4 treated pregnancies.

Furthermore, to investigate the possible effect of mode of transport, as a post-hoc analysis we removed samples from pregnancies with Caesarean section from the regression analyses (total of 16 cases removed). Again, the correlations between CBTSH levels and cord serum concentrations of lactate (β = 0.44, p < 0.0001) and alanine (β = 0.29, p = 0.0070) remained significant in the LT4 treated pregnancies. Moreover, also the negative correlations between Apgar scores at 1 min and 5 min and cord serum concentrations of lactate (β = −0.25, p = 0.0167; β = −0.27, p = 0.0085, respectively) and alanine (β = −0.24, p = 0.0223; β = −0.22, p = 0.0345) had p-values below 0.05 in the LT4 treated pregnancies after exclusion samples from Caesarean section births.

Furthermore, in the serum samples collected during delivery from the pregnant women, ketone bodies 3-hydroxybutyrate and acetoacetate significantly correlated negatively with Apgar scores at 1 minute after birth in the LT4 treated pregnancies, but not in control pregnancies (Figure 3, z = 2.81, p = 0.0050, and z = 2.33, p = 0.0198, respectively). All correlations between metabolite variables measured from during delivery serum samples and variables from the newborns are shown in Table S3.

Since placental weight could contribute to the altered fetal glycolysis, protein catabolism, and Apgar score at delivery, we analyzed whether there is any correlation between maternal or cord blood TSH and placental weight in LT4 treated pregnancies. Placenta weight did not correlate with the CBTSH or the TSH concentration of the pregnant women (β = −0.09, p = 0.3336, β = 0.06, p = 0.3295, respectively). However, there was a negative trend between the placenta weight and the pregnant women’s T4 concentration (β = −0.21, p = 0.0243). Moreover, there were no significant correlations between the CBTSH and the TSH (β = −0.09, p = 0.3396) or T4 (β = −0.12, p = 0.2197) concentrations of the pregnant women. Furthermore, maternal serum TSH or T4 concentrations did not correlate with Apgar scores measured 1 min (β = 0.11, p = 0.2395, β = 0.05, p = 0.6160, respectively) and 5 min (β = 0.10, p = 0.3035, β = 0.04, p = 0.6712, respectively) after birth.

Moreover, in the serum samples collected from the women during the 1^st^ trimester of the pregnancy (98 cases, 118 controls), there were significant negative correlations between Apgar scores at 1 min after birth and concentrations of phosphoglycerides (β = −0.31, p = 0.0024), phospholipids in lipoprotein particles (β = −0.31, p = 0.0025), and total cholines (β = −0.31, p = 0.0021, Table S3). All correlations between metabolite variables measured from 1^st^ trimester serum samples and variables from the newborn are shown in Table S4.

## Discussion

LT4 treatment is commonly used to treat both overt and subclinical hypothyroidism during pregnancy. However, there is only one report examining the impact of LT4 treatment on cord blood metabolite profile ^17^. To our knowledge, this is the first study to investigate the longitudinal effects of LT4 treatment on the metabolite profile during pregnancy. Our main findings were the associations between altered energy and amino acid metabolism, CBTSH concentrations and Apgar scores in newborn babies from LT4-treated pregnancies.

In the present study, newborn CBTSH concentrations were negatively correlated with Apgar scores (at 1 and 5 minutes) of the newborns in LT4 treated pregnancies, but not in the babies born to the control women. Previously, CBTSH levels have been reported to be higher in newborns with lower Apgar scores (<5) ^24^. Additionally, the CBTSH concentration has been observed to be elevated in cases with a higher gestational age at birth and with increasing maternal age as well as with the history of maternal hypothyroidism ^25^. There are preliminary reports that both maternal hypo- and hyperthyroidism might be associated with lower Apgar scores ^26,27^. In contrast, LT4 treatment has previously been associated with a reduced risk of low (<7) Apgar scores in women with subclinical hypothyroidism ^28^. It could be that the relationship between CBTSH and Apgar scores is an inverted U shape, where both very high and very low CBTSH concentrations could be associated with low Apgar scores. It should be noted that the maternal TSH and T4 levels did not correlate with CBTSH levels in LT4 treated pregnancies, in line with previous literature ^29^. LT4 treatment has not been shown to affect gestational length, nor was such an effect seen in this study.

The NMR metabolomics analysis revealed that low Apgar scores were associated with changes in the cord serum metabolic profile in LT4-treated pregnancies. There were significant positive correlations between CBTSH and lactate and alanine levels, but when analyzed with regression analyses where both CBTSH and cord serum lactate or alanine concentrations were both covariates, significant negative correlations were only seen between Apgar scores and lactate and alanine in the LT4 treated pregnancies, showing that levels of these metabolites contribute relatively more in explaining the Apgar scores than CBTSH. High lactate and alanine levels indicate increased anaerobic glycolysis and protein degradation in muscles in LT4 treated pregnancies with low Apgar values. Moreover, the increased protein degradation could also explain the significant negative correlations between Apgar scores and cord serum concentrations of most of the measured amino acids in the LT4 treated pregnancies. Muscle tissue is important in regulating energy consumption and glucose homeostasis, and it well-known that thyroid hormones can influence the metabolic properties of muscles ^30^. It has been shown that LT4 exposure increases anaerobic glycolysis and lactate formation in skeletal muscle cells *in vitro* ^31–33^. Furthermore, in an experiment conducted in rats, LT4 also increased glucose production from alanine ^32^. These effects can explain the positive correlation between CBTSH and lactate and alanine levels observed in both control and LT4 treated pregnancies. However, it is unclear why an increase in the CBTSH levels would negatively correlate with low Apgar scores only in the children of LT4-treated women. This could be linked to the observations that high LT4 levels stimulate the maturation of muscle cells ^34–36^, which might allow the effects of LT4 to induce anaerobic glycolysis and muscle protein degradation to be more prominent in LT4 treated pregnancies. This alteration in the energy metabolism seen in the cord serum samples, might also be linked to the association observed between low Apgar scores and high concentrations of ketone bodies in the maternal serum samples taken during delivery in the LT4 treated pregnancies. It could be that the hypothalamic-pituitary-thyroid axis is more responsive in the LT4 treated pregnancies, which could explain the CBTSH and lactate levels, which both rise during delivery, and that is reflected in the Apgar scores of the baby.

In the present study, we also observed that extensive changes occurred in the serum metabolite profile during pregnancy. This is in line with previous longitudinal metabolomics studies, where the circulating metabolome of the women exhibited very large alterations during pregnancy ^11,13,14^. It is well known that pregnancy triggers many physiological changes, including major alterations in lipid metabolism ^37,38^. Our results of metabolomics analysis also revealed increases in nearly all of the markers related to cholesterol metabolism during pregnancy, which is in line with previous reports of increases in plasma cholesterol levels and triglyceride levels ^37,38^. These elevated levels ensure that adequate amounts of cholesterols and lipids are available for normal fetal development including steroid production in placenta ^39,40^.

A reduced increase was observed in the concentrations of cholesteryl esters in small VLDL, cholesterol in small VLDL, free cholesterol in very small VLDL, and phospholipids in very small VLDL in the LT4-treated pregnancies when compared to controls (Figure 1). Thyroid hormones have an important role in the regulation of cholesterol biosynthesis and thus a thyroid dysfunction can influence lipid profiles ^41,42^. Due to the reduced activity of LDL receptors and decreased catabolism of LDL and intermediate-density lipoproteins (IDL), patients with overt hypothyroidism and subclinical hypothyroidism exhibit elevated levels of total cholesterol, LDL and VLDL. Additionally, a reduction of hepatic lipase, an enzyme whose function is stimulated by thyroid hormones, would decrease the catabolism of HDL2 and thus increase the levels of HDL in cases of overt or subclinical hypothyroidism. In the non-pregnant population, LT4 therapy has been associated with decreased cholesterol, lipoprotein and lipid levels ^43^.

Furthermore, significant associations were observed between low Apgar scores at 1 minute and high levels of cord serum metabolomics variables associated with cholesterol, lipids and lipoproteins in both LT4-treated pregnancies and control pregnancies. This is in line with the previous literature, where high cholesterol and lipid levels in the cord blood or in newborn babies have been associated with low Apgar scores ^44–46^. These types of alterations in cholesterol and lipid metabolism are encountered in pregnancy complications like gestational diabetes mellitus (GDM) ^47^, which has also been linked to lower Apgar scores ^48^. However, it should be noted that in the present study, there were no significant alterations in the cord blood metabolite levels when GDM pregnancies were compared to non-GDM pregnancies (data not shown). GDM has been associated with an increased risk of long term complications, e.g., the metabolic syndrome, in both the pregnant women and newborns ^49,50^. Therefore, further studies are needed to investigate if the high cholesterol and lipid levels in the cord blood would be associated with an increased risk of metabolic complications or neurocognitive development after birth.

One strength of this study is that it included samples collected from 1^st^ trimester and during delivery from the pregnant woman as well as cord blood samples. This allowed us to compare longitudinal changes in the maternal circulating metabolite profile during pregnancy and also differences between the fetal and maternal compartments during pregnancy. Since hypothyroidism during pregnancy is invariably treated, it was impossible to enroll women with untreated hypothyroidism into the study. Therefore, separating the effect of LT4 treatment for hypothyroidism with the disease of hypothyroidism was not possible. However, LT4 is the most commonly used medication for treatment of hypothyroidism. In our study, the dose was not uniform in all the women and in some women the dose was adjusted during pregnancy. There are no data available on the etiology of hypothyroidism, but other than autoimmune conditions are rare in the age group of pregnant women, and of these the majority have Hashimoto disease. However, it cannot ruled out that autoimmune diseases could modify metabolomics profile irrespectively of thyroid hormone levels.

### Conclusions

In this study, nearly all measured cholesterol and lipoprotein variables were increased during pregnancy in the maternal samples. Furthermore, LT4 treatment might cause small disturbances in this process, observed here as a reduced increase in the concentrations of cholesteryl esters, cholesterol and phospholipids in (very) small VLDL during pregnancy. Moreover, the associations between the cord serum metabolite profile and Apgar scores of the newborn differed between LT4 treated pregnancies and control pregnancies in those metabolites associated with energy metabolism and amino acid metabolism. These results indicate that high CBTSH concentrations are associated with alterations in the cord serum levels of the metabolites which have been linked with hypoxia and muscle degradation. These changes might explain the negative correlation between the CBTSH level and Apgar scores in LT4-treated pregnancies.

## Acknowledgements

This paper belongs to the studies carried out by the Kuopio Birth Cohort consortium (www.KuBiCo.fi) and we thank our colleagues who are responsible for the design and conduct of the KuBiCo. We thank Ms. Pirjo Hänninen, Ms. Sonja Holopainen and MSc. Essi Järvelä for expert laboratory assistance, Ewen MacDonald for proof-reading, and the staff of the Department of Obstetrics and Gynecology in Kuopio University Hospital.

## Declaration of Interest statement

Heidi Sahlman, Leea Keski-Nisula, and Jaana Rysä report no conflict of interest. Olli Kärkkäinen is affiliated with a metabolomics analysis service company Afekta Technologies Ltd (not used here).

## Author Contributions statement

Olli Kärkkäinen, Heidi Sahlman, Leea Keski-Nisula and Jaana Rysä contributed to the conception and design of the study; Olli Kärkkäinen, Leea Keski-Nisula and Heidi Sahlman contributed analysis and interpretation of the data; Olli Kärkkäinen and Heidi Sahlman drafted the paper, all the authors have critically reviewed and approved the final manuscript.

## Data Availability statement

The data used in this study are not publicly available because they contain identification information. However, some parts of the data may be requested from the corresponding author (JR) upon reasonable request.

## Financial Support

This work was supported by the European Union’s Horizon 2020 research and innovation programme under grant agreement No 825762 and by a grant from the Avohoidon tutkimussäätiö.

## Notes

### Competing Interest Statement

HS, LKN, and JR report no conflict of interest. OK is affiliated with a metabolomics analysis service company Afekta Technologies Ltd (not used here).

### Funding Statement

This work was supported by the European Union Horizon 2020 research and innovation programme under grant agreement No 825762 and by a grant from the Avohoidon tutkimussäätiö.

### Author Declarations

The Research Ethics Committee of Hospital District of Central Finland gave ethical approval for this work.

