## Supplementary material for "Levothyroxine treatment during pregnancy: a metabolomics study": Table S1

Supplementary table 1: NMR metabolomics results

| | Case-control comparisons | | | | | | | | | | | | | | | | | | Change during pregnancy | | | |
|---|---|---|---|---|---|---|---|---|---|---|---|---|---|---|---|---|---|---|---|---|---|---|
| | 1st trimester | | | | | | Delivery | | | | | | Cord serum | | | | | | Delivery vs. 1st trimester | | | |
| | LT4 | | Control | | | | LT4 | | Control | | | | LT4 | | Control | | | | All subjects | | LT4 vs. controls | |
| Metabolite abbreviation | Mean | SD | Mean | SD | p | d | Mean | SD | Mean | SD | p | d | Mean | SD | Mean | SD | p | d | p | d | p | d |
| Total cholesterol | 4.80 | 0.69 | 4.68 | 0.93 | 2.93E-01 | 0.14 | 6.97 | 1.29 | 7.27 | 1.40 | 8.54E-02 | -0.23 | 1.88 | 0.41 | 1.84 | 0.46 | 4.59E-01 | 0.10 | 8.31E-76 | 2.18 | 0.0418 | -0.28 |
| Total cholesterol minus HDL-C | 3.02 | 0.63 | 2.95 | 0.77 | 4.67E-01 | 0.10 | 5.09 | 1.21 | 5.41 | 1.27 | 4.60E-02 | -0.26 | 1.11 | 0.30 | 1.06 | 0.33 | 2.22E-01 | 0.17 | 6.44E-80 | 2.32 | 0.0275 | -0.30 |
| Remnant cholesterol | 1.37 | 0.32 | 1.34 | 0.38 | 5.42E-01 | 0.08 | 2.54 | 0.64 | 2.73 | 0.65 | 2.46E-02 | -0.29 | 0.56 | 0.14 | 0.53 | 0.16 | 2.23E-01 | 0.17 | 1.34E-86 | 2.56 | 0.0182 | -0.32 |
| VLDL cholesterol | 0.52 | 0.19 | 0.51 | 0.20 | 9.03E-01 | 0.02 | 1.26 | 0.42 | 1.37 | 0.39 | 4.02E-02 | -0.27 | 0.23 | 0.09 | 0.22 | 0.09 | 1.66E-01 | 0.19 | 6.58E-89 | 2.65 | 0.0502 | -0.27 |
| Clinical LDL cholesterol | 2.30 | 0.52 | 2.23 | 0.63 | 3.73E-01 | 0.12 | 3.62 | 0.99 | 3.86 | 1.09 | 8.16E-02 | -0.23 | 0.70 | 0.25 | 0.66 | 0.28 | 2.92E-01 | 0.14 | 1.12E-61 | 1.82 | 0.0370 | -0.29 |
| LDL cholesterol | 1.65 | 0.32 | 1.61 | 0.40 | 4.12E-01 | 0.11 | 2.54 | 0.60 | 2.68 | 0.65 | 9.78E-02 | -0.22 | 0.56 | 0.16 | 0.53 | 0.18 | 2.43E-01 | 0.16 | 7.41E-69 | 1.99 | 0.0519 | -0.27 |
| HDL cholesterol | 1.78 | 0.29 | 1.73 | 0.31 | 2.56E-01 | 0.16 | 1.88 | 0.42 | 1.86 | 0.44 | 7.05E-01 | 0.05 | 0.77 | 0.18 | 0.78 | 0.18 | 7.36E-01 | -0.05 | 9.70E-06 | 0.31 | 0.7463 | -0.04 |
| Total TGs | 1.19 | 0.48 | 1.19 | 0.53 | 9.95E-01 | 0.00 | 2.82 | 1.23 | 2.83 | 0.95 | 9.52E-01 | -0.01 | 0.50 | 0.19 | 0.48 | 0.18 | 3.81E-01 | 0.12 | 2.67E-69 | 2.04 | 0.7977 | 0.04 |
| TGs in VLDL | 0.73 | 0.40 | 0.74 | 0.44 | 9.59E-01 | -0.01 | 1.86 | 1.08 | 1.81 | 0.81 | 6.89E-01 | 0.05 | 0.28 | 0.14 | 0.27 | 0.13 | 3.89E-01 | 0.12 | 5.57E-53 | 1.60 | 0.4226 | 0.11 |
| TGs in LDL | 0.16 | 0.04 | 0.16 | 0.04 | 8.53E-01 | 0.03 | 0.36 | 0.09 | 0.38 | 0.08 | 2.98E-02 | -0.28 | 0.08 | 0.02 | 0.08 | 0.02 | 5.08E-01 | 0.09 | 3.77E-99 | 3.33 | 0.0311 | -0.29 |
| TGs in HDL | 0.18 | 0.05 | 0.18 | 0.06 | 8.34E-01 | 0.03 | 0.36 | 0.09 | 0.37 | 0.08 | 1.33E-01 | -0.20 | 0.07 | 0.02 | 0.07 | 0.02 | 4.65E-01 | 0.10 | 3.64E-90 | 2.75 | 0.1618 | -0.19 |
| Phospholipids in lipoprotein particles | 3.22 | 0.41 | 3.16 | 0.54 | 3.22E-01 | 0.14 | 4.53 | 0.65 | 4.63 | 0.67 | 2.21E-01 | -0.16 | 1.47 | 0.22 | 1.45 | 0.25 | 5.81E-01 | 0.07 | 2.65E-84 | 2.44 | 0.1521 | -0.20 |
| Phospholipids in VLDL | 0.35 | 0.14 | 0.35 | 0.15 | 9.91E-01 | 0.00 | 0.92 | 0.35 | 0.97 | 0.30 | 2.10E-01 | -0.16 | 0.15 | 0.07 | 0.14 | 0.06 | 2.13E-01 | 0.17 | 3.48E-85 | 2.52 | 0.2959 | -0.14 |
| Phospholipids in LDL | 0.56 | 0.10 | 0.55 | 0.12 | 4.05E-01 | 0.11 | 0.86 | 0.19 | 0.92 | 0.21 | 4.28E-02 | -0.27 | 0.24 | 0.05 | 0.24 | 0.05 | 2.70E-01 | 0.15 | 1.89E-72 | 2.10 | 0.0203 | -0.32 |
| Phospholipids in HDL | 2.03 | 0.30 | 1.98 | 0.34 | 2.97E-01 | 0.14 | 2.29 | 0.42 | 2.26 | 0.44 | 5.43E-01 | 0.08 | 0.93 | 0.16 | 0.93 | 0.16 | 8.28E-01 | -0.03 | 6.31E-19 | 0.72 | 0.9607 | -0.01 |
| Total esterified cholesterol | 3.54 | 0.49 | 3.45 | 0.67 | 2.70E-01 | 0.15 | 4.98 | 0.92 | 5.18 | 1.00 | 1.05E-01 | -0.21 | 1.37 | 0.31 | 1.34 | 0.34 | 4.94E-01 | 0.09 | 6.66E-71 | 2.04 | 0.0490 | -0.27 |
| CEs in VLDL | 0.31 | 0.11 | 0.31 | 0.12 | 8.84E-01 | 0.02 | 0.73 | 0.23 | 0.80 | 0.23 | 1.25E-02 | -0.33 | 0.14 | 0.05 | 0.14 | 0.05 | 1.52E-01 | 0.19 | 2.95E-88 | 2.67 | 0.0148 | -0.34 |
| CEs in LDL | 1.20 | 0.25 | 1.17 | 0.30 | 4.52E-01 | 0.10 | 1.91 | 0.44 | 2.01 | 0.48 | 1.03E-01 | -0.21 | 0.39 | 0.12 | 0.37 | 0.13 | 2.19E-01 | 0.17 | 1.39E-73 | 2.11 | 0.0608 | -0.26 |
| CEs in HDL | 1.37 | 0.22 | 1.34 | 0.24 | 2.47E-01 | 0.16 | 1.38 | 0.33 | 1.36 | 0.34 | 5.91E-01 | 0.07 | 0.61 | 0.13 | 0.61 | 0.13 | 7.10E-01 | -0.05 | 3.81E-01 | 0.06 | 0.8628 | -0.02 |
| Total free cholesterol | 1.27 | 0.20 | 1.24 | 0.26 | 3.64E-01 | 0.12 | 1.99 | 0.39 | 2.09 | 0.41 | 5.77E-02 | -0.25 | 0.51 | 0.11 | 0.50 | 0.12 | 3.75E-01 | 0.12 | 6.20E-85 | 2.47 | 0.0322 | -0.29 |
| Free cholesterol in VLDL | 0.21 | 0.08 | 0.21 | 0.09 | 9.32E-01 | 0.01 | 0.53 | 0.20 | 0.57 | 0.17 | 1.61E-01 | -0.18 | 0.09 | 0.04 | 0.08 | 0.04 | 1.96E-01 | 0.18 | 1.76E-85 | 2.51 | 0.2156 | -0.17 |
| Free cholesterol in LDL | 0.45 | 0.08 | 0.44 | 0.10 | 3.24E-01 | 0.14 | 0.64 | 0.16 | 0.67 | 0.18 | 9.85E-02 | -0.22 | 0.17 | 0.05 | 0.16 | 0.05 | 3.36E-01 | 0.13 | 1.69E-52 | 1.59 | 0.0394 | -0.28 |
| Free cholesterol in HDL | 0.41 | 0.07 | 0.40 | 0.08 | 3.03E-01 | 0.14 | 0.50 | 0.10 | 0.50 | 0.11 | 8.67E-01 | -0.02 | 0.16 | 0.04 | 0.16 | 0.04 | 8.25E-01 | -0.03 | 9.16E-37 | 1.10 | 0.4330 | -0.11 |
| Lipids in lipoprotein particles | 9.21 | 1.38 | 9.03 | 1.78 | 4.04E-01 | 0.11 | 14.31 | 2.55 | 14.73 | 2.49 | 2.05E-01 | -0.17 | 3.85 | 0.69 | 3.77 | 0.76 | 3.95E-01 | 0.12 | 4.27E-91 | 2.62 | 0.1595 | -0.19 |
| Total lipids in VLDL | 1.60 | 0.70 | 1.60 | 0.75 | 9.96E-01 | 0.00 | 4.03 | 1.75 | 4.15 | 1.40 | 5.89E-01 | -0.07 | 0.67 | 0.29 | 0.62 | 0.27 | 2.45E-01 | 0.16 | 1.63E-74 | 2.16 | 0.8073 | -0.03 |
| Total lipids in LDL | 2.37 | 0.45 | 2.32 | 0.55 | 4.25E-01 | 0.11 | 3.77 | 0.84 | 3.98 | 0.90 | 6.13E-02 | -0.25 | 0.88 | 0.22 | 0.84 | 0.25 | 2.47E-01 | 0.16 | 2.58E-76 | 2.21 | 0.0322 | -0.29 |
| Total lipids in HDL | 3.99 | 0.61 | 3.89 | 0.67 | 2.82E-01 | 0.15 | 4.53 | 0.83 | 4.49 | 0.89 | 7.29E-01 | 0.05 | 1.77 | 0.33 | 1.78 | 0.34 | 8.13E-01 | -0.03 | 4.20E-21 | 0.76 | 0.7614 | -0.04 |
| Conc. of lipoprotein particles | 0.02 | 0.00 | 0.02 | 0.00 | 1.60E-01 | 0.19 | 0.02 | 0.00 | 0.02 | 0.00 | 8.93E-01 | 0.02 | 0.01 | 0.00 | 0.01 | 0.00 | 8.07E-01 | 0.03 | 1.47E-33 | 1.08 | 0.5856 | -0.07 |
| Conc. of VLDL particles | 0.00 | 0.00 | 0.00 | 0.00 | 8.88E-01 | 0.02 | 0.00 | 0.00 | 0.00 | 0.00 | 4.56E-02 | -0.26 | 0.00 | 0.00 | 0.00 | 0.00 | 2.22E-01 | 0.17 | 2.37E-92 | 2.79 | 0.0585 | -0.26 |
| Conc. of LDL particles | 0.00 | 0.00 | 0.00 | 0.00 | 6.99E-01 | 0.05 | 0.00 | 0.00 | 0.00 | 0.00 | 2.96E-02 | -0.29 | 0.00 | 0.00 | 0.00 | 0.00 | 1.40E-01 | 0.20 | 6.97E-80 | 2.33 | 0.0241 | -0.31 |
| Conc. of HDL particles | 0.02 | 0.00 | 0.02 | 0.00 | 1.59E-01 | 0.19 | 0.02 | 0.00 | 0.02 | 0.00 | 5.53E-01 | 0.08 | 0.01 | 0.00 | 0.01 | 0.00 | 9.19E-01 | 0.01 | 2.86E-17 | 0.69 | 0.8475 | -0.03 |
| Average diameter for VLDL particles | 37.57 | 1.18 | 37.55 | 1.28 | 9.25E-01 | 0.01 | 38.54 | 1.42 | 38.39 | 1.04 | 3.29E-01 | 0.13 | 36.02 | 1.05 | 35.84 | 1.11 | 2.18E-01 | 0.17 | 1.62E-22 | 0.73 | 0.6078 | 0.07 |
| Average diameter for LDL particles | 23.95 | 0.07 | 23.96 | 0.07 | 4.27E-01 | -0.11 | 23.93 | 0.08 | 23.95 | 0.07 | 3.43E-02 | -0.28 | 23.90 | 0.05 | 23.90 | 0.05 | 7.75E-01 | 0.04 | 1.24E-02 | -0.19 | 0.9394 | 0.01 |
| Average diameter for HDL particles | 9.90 | 0.19 | 9.89 | 0.18 | 7.73E-01 | 0.04 | 9.99 | 0.20 | 10.00 | 0.21 | 7.65E-01 | -0.04 | 9.61 | 0.22 | 9.62 | 0.23 | 6.71E-01 | -0.06 | 5.96E-14 | 0.50 | 0.9934 | 0.00 |
| Phosphoglycerides | 2.77 | 0.38 | 2.71 | 0.49 | 3.17E-01 | 0.14 | 3.83 | 0.55 | 3.89 | 0.57 | 4.20E-01 | -0.11 | 1.09 | 0.19 | 1.08 | 0.20 | 6.53E-01 | 0.06 | 3.13E-78 | 2.25 | 0.2911 | -0.14 |
| Ratio of TGs to phosphoglycerides | 0.42 | 0.14 | 0.43 | 0.15 | 7.24E-01 | -0.05 | 0.73 | 0.29 | 0.73 | 0.22 | 9.80E-01 | 0.00 | 0.46 | 0.16 | 0.45 | 0.16 | 6.00E-01 | 0.07 | 5.66E-54 | 1.50 | 0.5952 | 0.07 |
| Total cholines | 3.04 | 0.37 | 2.98 | 0.49 | 3.09E-01 | 0.14 | 4.09 | 0.55 | 4.17 | 0.59 | 2.79E-01 | -0.14 | 1.30 | 0.20 | 1.29 | 0.22 | 7.17E-01 | 0.05 | 5.88E-77 | 2.21 | 0.1983 | -0.18 |
| Phosphatidylcholines | 2.69 | 0.38 | 2.63 | 0.48 | 3.47E-01 | 0.13 | 3.74 | 0.54 | 3.81 | 0.57 | 3.44E-01 | -0.12 | 1.03 | 0.17 | 1.01 | 0.18 | 5.85E-01 | 0.07 | 7.24E-78 | 2.26 | 0.2411 | -0.16 |
| Sphingomyelins | 0.50 | 0.06 | 0.49 | 0.08 | 1.74E-01 | 0.19 | 0.66 | 0.12 | 0.68 | 0.11 | 9.87E-02 | -0.22 | 0.25 | 0.04 | 0.25 | 0.05 | 5.44E-01 | 0.08 | 1.31E-63 | 1.95 | 0.0394 | -0.28 |
| Apolipoprotein B | 0.73 | 0.15 | 0.72 | 0.17 | 6.55E-01 | 0.06 | 1.27 | 0.31 | 1.37 | 0.33 | 2.37E-02 | -0.30 | 0.40 | 0.07 | 0.39 | 0.07 | 1.49E-01 | 0.20 | 1.52E-83 | 2.45 | 0.0184 | -0.32 |
| Apolipoprotein A1 | 1.82 | 0.22 | 1.78 | 0.26 | 2.28E-01 | 0.17 | 2.05 | 0.32 | 2.03 | 0.34 | 6.44E-01 | 0.06 | 0.89 | 0.12 | 0.90 | 0.13 | 9.13E-01 | -0.01 | 2.39E-23 | 0.84 | 0.8122 | -0.03 |
| Ratio of apolipoprotein B to apolipoprotein A1 | 0.41 | 0.10 | 0.41 | 0.09 | 9.75E-01 | 0.00 | 0.64 | 0.20 | 0.69 | 0.21 | 4.51E-02 | -0.26 | 0.45 | 0.08 | 0.43 | 0.07 | 9.19E-02 | 0.23 | 6.95E-58 | 1.70 | 0.0450 | -0.27 |
| Total FAs | 12.73 | 2.17 | 12.50 | 2.69 | 4.87E-01 | 0.09 | 20.61 | 4.17 | 20.90 | 3.52 | 5.64E-01 | -0.08 | 5.20 | 0.93 | 5.09 | 0.99 | 4.28E-01 | 0.11 | 7.23E-92 | 2.58 | 0.5439 | -0.08 |
| Degree of unsaturation | 1.37 | 0.06 | 1.36 | 0.06 | 6.97E-02 | 0.25 | 1.26 | 0.09 | 1.25 | 0.07 | 2.87E-01 | 0.14 | 1.37 | 0.08 | 1.38 | 0.08 | 7.22E-01 | -0.05 | 3.79E-56 | -1.55 | 0.9374 | 0.01 |
| Omega-3 FAs | 0.68 | 0.19 | 0.63 | 0.19 | 5.65E-02 | 0.26 | 0.91 | 0.26 | 0.88 | 0.27 | 4.59E-01 | 0.10 | 0.31 | 0.14 | 0.29 | 0.15 | 4.85E-01 | 0.09 | 2.25E-36 | 1.04 | 0.8959 | -0.02 |
| Omega-6 FAs | 4.80 | 0.56 | 4.71 | 0.76 | 3.38E-01 | 0.13 | 6.47 | 1.04 | 6.59 | 1.04 | 3.94E-01 | -0.11 | 2.17 | 0.26 | 2.16 | 0.29 | 8.63E-01 | 0.02 | 4.43E-73 | 2.07 | 0.3871 | -0.12 |
| PUFAs | 5.48 | 0.70 | 5.35 | 0.90 | 2.13E-01 | 0.17 | 7.38 | 1.18 | 7.47 | 1.18 | 5.59E-01 | -0.08 | 2.47 | 0.35 | 2.45 | 0.37 | 6.80E-01 | 0.06 | 1.52E-72 | 2.02 | 0.4306 | -0.11 |
| MUFAs | 2.89 | 0.72 | 2.84 | 0.84 | 6.74E-01 | 0.06 | 5.47 | 1.73 | 5.54 | 1.25 | 7.06E-01 | -0.05 | 0.79 | 0.31 | 0.75 | 0.28 | 3.83E-01 | 0.12 | 3.99E-80 | 2.31 | 0.7765 | -0.04 |
| Saturated FAs | 4.36 | 0.86 | 4.31 | 1.06 | 7.04E-01 | 0.05 | 7.76 | 1.72 | 7.89 | 1.36 | 5.34E-01 | -0.08 | 1.94 | 0.37 | 1.89 | 0.40 | 3.64E-01 | 0.12 | 5.08E-94 | 2.77 | 0.5611 | -0.08 |
| LA | 4.01 | 0.55 | 3.96 | 0.75 | 5.58E-01 | 0.08 | 5.52 | 1.04 | 5.67 | 1.07 | 2.77E-01 | -0.14 | 1.14 | 0.24 | 1.14 | 0.28 | 9.75E-01 | 0.00 | 1.43E-65 | 1.88 | 0.3386 | -0.13 |
| DHA | 0.34 | 0.06 | 0.32 | 0.06 | 3.84E-02 | 0.29 | 0.39 | 0.10 | 0.39 | 0.10 | 7.98E-01 | 0.03 | 0.18 | 0.06 | 0.17 | 0.06 | 5.08E-01 | 0.09 | 2.05E-20 | 0.74 | 0.4592 | -0.10 |
| Ratio of omega-3 FAs to total FAs | 5.34 | 1.12 | 5.05 | 0.95 | 4.24E-02 | 0.28 | 4.40 | 0.95 | 4.21 | 1.11 | 1.62E-01 | 0.18 | 5.72 | 1.82 | 5.56 | 2.06 | 5.54E-01 | 0.08 | 2.02E-31 | -0.84 | 0.6777 | -0.06 |
| Ratio of omega-6 FAs to total FAs | 38.07 | 2.75 | 38.25 | 3.26 | 6.60E-01 | -0.06 | 31.93 | 5.09 | 31.67 | 2.67 | 6.11E-01 | 0.07 | 42.34 | 4.67 | 43.06 | 4.43 | 2.47E-01 | -0.16 | 8.83E-60 | -1.80 | 0.4976 | 0.09 |
| Ratio of PUFA to total FAs | 43.41 | 2.79 | 43.30 | 3.31 | 7.87E-01 | 0.04 | 36.34 | 5.16 | 35.88 | 2.87 | 4.01E-01 | 0.11 | 48.06 | 3.79 | 48.62 | 3.43 | 2.55E-01 | -0.15 | 2.01E-67 | -2.00 | 0.5685 | 0.08 |
| Ratio of MUFAs to total FAs | 22.45 | 2.11 | 22.36 | 2.44 | 7.87E-01 | 0.04 | 26.09 | 3.74 | 26.34 | 2.33 | 5.28E-01 | -0.08 | 14.72 | 3.94 | 14.39 | 3.25 | 5.01E-01 | 0.09 | 1.48E-45 | 1.41 | 0.6637 | -0.06 |
| Ratio of saturated FAs to total FAs | 34.14 | 1.57 | 34.34 | 1.67 | 3.76E-01 | -0.12 | 37.57 | 2.45 | 37.78 | 1.81 | 4.74E-01 | -0.09 | 37.22 | 1.70 | 36.99 | 1.78 | 3.33E-01 | 0.13 | 6.96E-55 | 1.81 | 0.8813 | -0.02 |
| Ratio of LA to total FAs | 31.78 | 2.82 | 31.93 | 2.97 | 6.92E-01 | -0.05 | 27.21 | 5.06 | 27.17 | 2.93 | 9.53E-01 | 0.01 | 22.05 | 3.39 | 22.36 | 3.65 | 5.16E-01 | -0.09 | 1.04E-44 | -1.33 | 0.7033 | 0.05 |
| Ratio of DHA to total FAs | 2.67 | 0.46 | 2.59 | 0.43 | 1.66E-01 | 0.19 | 1.93 | 0.47 | 1.87 | 0.47 | 3.02E-01 | 0.13 | 3.37 | 0.67 | 3.33 | 0.79 | 6.79E-01 | 0.06 | 1.89E-64 | -1.59 | 0.8156 | -0.03 |
| Ratio of PUFAs to MUFAs | 1.96 | 0.28 | 1.99 | 0.58 | 5.88E-01 | -0.08 | 1.39 | 0.26 | 1.38 | 0.22 | 6.48E-01 | 0.06 | 3.57 | 1.23 | 3.60 | 1.05 | 8.39E-01 | -0.03 | 1.65E-48 | -1.67 | 0.4771 | 0.10 |
| Ratio of omega-6 FAs to omega-3 FAs | 7.46 | 1.79 | 7.93 | 2.25 | 9.15E-02 | -0.23 | 7.62 | 2.24 | 8.07 | 2.39 | 1.38E-01 | -0.19 | 8.57 | 4.81 | 9.14 | 4.49 | 3.66E-01 | -0.12 | 3.43E-01 | 0.06 | 0.9534 | 0.01 |
| Alanine | 0.40 | 0.06 | 0.41 | 0.07 | 1.29E-01 | -0.21 | 0.48 | 0.09 | 0.48 | 0.08 | 5.59E-01 | 0.08 | 0.56 | 0.12 | 0.56 | 0.11 | 8.87E-01 | 0.02 | 7.58E-24 | 0.98 | 0.1156 | 0.22 |
| Glutamine | 0.54 | 0.05 | 0.55 | 0.06 | 2.14E-01 | -0.17 | 0.52 | 0.10 | 0.51 | 0.08 | 4.18E-01 | 0.11 | 0.63 | 0.11 | 0.63 | 0.08 | 8.05E-01 | -0.03 | 1.00E-08 | -0.52 | 0.1994 | 0.18 |
| Glycine | 0.21 | 0.04 | 0.21 | 0.06 | 4.86E-01 | -0.10 | 0.21 | 0.05 | 0.20 | 0.05 | 4.76E-02 | 0.26 | 0.28 | 0.07 | 0.27 | 0.06 | 5.84E-01 | 0.07 | 7.54E-02 | -0.15 | 0.0042 | 0.39 |
| Histidine | 0.10 | 0.01 | 0.10 | 0.01 | 9.98E-01 | 0.00 | 0.09 | 0.02 | 0.09 | 0.01 | 9.74E-01 | 0.00 | 0.12 | 0.02 | 0.11 | 0.01 | 6.83E-01 | 0.06 | 1.13E-17 | -0.74 | 0.9980 | 0.00 |
| Total Conc. of BCAA | 0.42 | 0.10 | 0.43 | 0.10 | 3.89E-01 | -0.12 | 0.33 | 0.08 | 0.32 | 0.06 | 4.37E-01 | 0.10 | 0.44 | 0.08 | 0.44 | 0.07 | 8.85E-01 | 0.02 | 4.81E-29 | -1.19 | 0.1849 | 0.18 |

Supplementary table 1: NMR metabolomics results

| | Case-control comparisons | | | | | | | | | | | | | | | | | | Change during pregnancy | | | |
|---|---|---|---|---|---|---|---|---|---|---|---|---|---|---|---|---|---|---|---|---|---|---|
| | 1st trimester | | | | | | Delivery | | | | | | Cord serum | | | | | | Delivery vs. 1st trimester | | | |
| | LT4 | | Control | | | | LT4 | | Control | | | | LT4 | | Control | | | | All subjects | | LT4 vs. controls | |
| Metabolite abbreviation | Mean | SD | Mean | SD | p | d | Mean | SD | Mean | SD | p | d | Mean | SD | Mean | SD | p | d | p | d | p | d |
| Isoleucine | 0.06 | 0.02 | 0.06 | 0.02 | 6.89E-01 | -0.05 | 0.04 | 0.01 | 0.04 | 0.01 | 5.12E-01 | 0.09 | 0.06 | 0.02 | 0.06 | 0.01 | 9.53E-01 | -0.01 | 6.21E-30 | -1.26 | 0.4182 | 0.11 |
| Leucine | 0.12 | 0.04 | 0.13 | 0.04 | 4.91E-01 | -0.09 | 0.09 | 0.03 | 0.09 | 0.02 | 3.43E-01 | 0.12 | 0.12 | 0.03 | 0.12 | 0.02 | 5.64E-01 | 0.08 | 3.13E-27 | -1.19 | 0.1993 | 0.18 |
| Valine | 0.24 | 0.05 | 0.25 | 0.05 | 2.57E-01 | -0.16 | 0.20 | 0.04 | 0.20 | 0.03 | 5.30E-01 | 0.08 | 0.25 | 0.04 | 0.25 | 0.04 | 9.17E-01 | -0.01 | 2.47E-26 | -1.07 | 0.1489 | 0.20 |
| Phenylalanine | 0.08 | 0.01 | 0.08 | 0.01 | 1.23E-01 | -0.21 | 0.09 | 0.02 | 0.08 | 0.02 | 1.44E-01 | 0.19 | 0.09 | 0.02 | 0.09 | 0.01 | 9.34E-01 | 0.01 | 3.54E-09 | 0.51 | 0.0125 | 0.34 |
| Tyrosine | 0.07 | 0.02 | 0.07 | 0.02 | 3.72E-01 | -0.12 | 0.05 | 0.01 | 0.05 | 0.01 | 3.51E-01 | 0.12 | 0.07 | 0.02 | 0.07 | 0.01 | 5.48E-01 | 0.08 | 3.84E-24 | -1.05 | 0.1125 | 0.22 |
| Glucose | 5.24 | 1.55 | 4.98 | 0.79 | 1.31E-01 | 0.22 | 5.82 | 1.69 | 5.60 | 1.14 | 2.40E-01 | 0.16 | 5.16 | 1.58 | 4.92 | 1.18 | 2.08E-01 | 0.17 | 7.69E-07 | 0.46 | 0.8981 | 0.02 |
| Lactate | 2.40 | 0.69 | 2.57 | 0.74 | 8.45E-02 | -0.24 | 3.36 | 1.16 | 3.32 | 1.08 | 7.77E-01 | 0.04 | 6.85 | 2.24 | 6.87 | 2.06 | 9.47E-01 | -0.01 | 8.59E-18 | 0.92 | 0.2184 | 0.17 |
| Pyruvate | 0.06 | 0.03 | 0.06 | 0.04 | 3.06E-01 | -0.14 | 0.13 | 0.06 | 0.13 | 0.06 | 5.23E-01 | 0.08 | 0.16 | 0.06 | 0.16 | 0.06 | 7.67E-01 | -0.04 | 1.75E-36 | 1.49 | 0.2268 | 0.17 |
| Citrate | 0.06 | 0.01 | 0.06 | 0.01 | 9.76E-01 | 0.00 | 0.08 | 0.01 | 0.08 | 0.01 | 5.47E-01 | 0.08 | 0.05 | 0.01 | 0.05 | 0.01 | 8.94E-01 | 0.02 | 4.56E-61 | 1.95 | 0.4585 | 0.10 |
| Glycerol | 0.11 | 0.04 | 0.10 | 0.04 | 5.25E-01 | 0.09 | 0.17 | 0.05 | 0.16 | 0.05 | 3.93E-01 | 0.11 | 0.09 | 0.03 | 0.09 | 0.03 | 5.02E-01 | 0.09 | 6.23E-36 | 1.30 | 0.4836 | 0.10 |
| 3-Hydroxybutyrate | 0.04 | 0.04 | 0.04 | 0.04 | 8.14E-01 | 0.03 | 0.15 | 0.21 | 0.15 | 0.15 | 8.92E-01 | 0.02 | 0.19 | 0.21 | 0.16 | 0.12 | 2.40E-01 | 0.16 | 3.94E-15 | 0.97 | 0.7062 | 0.05 |
| Acetate | 0.04 | 0.02 | 0.04 | 0.02 | 8.91E-01 | 0.02 | 0.03 | 0.02 | 0.03 | 0.01 | 5.71E-01 | 0.07 | 0.03 | 0.01 | 0.03 | 0.01 | 5.61E-01 | 0.08 | 2.25E-14 | -0.66 | 0.6469 | 0.06 |
| Acetoacetate | 0.02 | 0.02 | 0.02 | 0.02 | 9.43E-01 | 0.01 | 0.06 | 0.07 | 0.05 | 0.04 | 6.12E-01 | 0.07 | 0.04 | 0.04 | 0.03 | 0.03 | 2.65E-01 | 0.15 | 2.96E-13 | 0.85 | 0.4928 | 0.10 |
| Acetone | 0.02 | 0.00 | 0.02 | 0.01 | 8.82E-02 | -0.23 | 0.02 | 0.02 | 0.02 | 0.01 | 2.20E-01 | 0.17 | 0.03 | 0.02 | 0.03 | 0.01 | 3.09E-01 | 0.14 | 9.62E-02 | 0.20 | 0.1008 | 0.24 |
| Creatinine | 59.08 | 7.34 | 56.88 | 7.24 | 2.93E-02 | 0.30 | 67.48 | 13.43 | 65.27 | 9.41 | 1.45E-01 | 0.19 | 59.34 | 12.00 | 57.49 | 11.77 | 2.59E-01 | 0.16 | 3.70E-28 | 0.89 | 0.4372 | 0.11 |
| Albumin | 39.61 | 3.33 | 39.98 | 3.30 | 4.15E-01 | -0.11 | 29.07 | 2.54 | 29.37 | 2.86 | 4.09E-01 | -0.11 | 37.83 | 3.77 | 38.41 | 3.48 | 2.45E-01 | -0.16 | 2.88E-114 | -3.52 | 0.9900 | 0.00 |
| Glycoprotein acetyls | 0.87 | 0.10 | 0.86 | 0.12 | 3.40E-01 | 0.13 | 1.04 | 0.12 | 1.03 | 0.11 | 5.37E-01 | 0.08 | 0.44 | 0.06 | 0.44 | 0.06 | 9.49E-01 | -0.01 | 4.22E-58 | 1.51 | 0.9500 | 0.01 |
| Conc. of CMs and extremely large VLDL particles | 0.00 | 0.00 | 0.00 | 0.00 | 7.55E-01 | -0.04 | 0.00 | 0.00 | 0.00 | 0.00 | 4.12E-01 | 0.11 | 0.00 | 0.00 | 0.00 | 0.00 | 3.49E-01 | 0.13 | 3.18E-45 | 1.54 | 0.2284 | 0.17 |
| Total lipids in CMs and extremely large VLDL | 0.07 | 0.13 | 0.07 | 0.13 | 7.85E-01 | -0.04 | 0.44 | 0.46 | 0.39 | 0.29 | 2.89E-01 | 0.14 | 0.04 | 0.04 | 0.03 | 0.03 | 3.81E-01 | 0.12 | 1.72E-36 | 1.34 | 0.1519 | 0.20 |
| Phospholipids in CMs and extremely large VLDL | 0.01 | 0.02 | 0.01 | 0.02 | 8.86E-01 | -0.02 | 0.07 | 0.07 | 0.06 | 0.04 | 4.80E-01 | 0.09 | 0.01 | 0.01 | 0.01 | 0.01 | 3.56E-01 | 0.13 | 1.84E-43 | 1.55 | 0.3245 | 0.14 |
| Cholesterol in CMs and extremely large VLDL | 0.02 | 0.02 | 0.02 | 0.02 | 8.94E-01 | -0.02 | 0.10 | 0.08 | 0.10 | 0.05 | 7.86E-01 | 0.04 | 0.01 | 0.01 | 0.01 | 0.01 | 2.95E-01 | 0.14 | 2.73E-55 | 1.80 | 0.5385 | 0.09 |
| CEs in CMs and extremely large VLDL | 0.01 | 0.01 | 0.01 | 0.01 | 9.04E-01 | -0.02 | 0.06 | 0.04 | 0.06 | 0.03 | 9.95E-01 | 0.00 | 0.01 | 0.01 | 0.00 | 0.01 | 2.49E-01 | 0.16 | 5.35E-61 | 1.92 | 0.7107 | 0.05 |
| Free cholesterol in CMs and extremely large VLDL | 0.01 | 0.01 | 0.01 | 0.01 | 8.83E-01 | -0.02 | 0.04 | 0.04 | 0.04 | 0.02 | 5.63E-01 | 0.08 | 0.00 | 0.00 | 0.00 | 0.00 | 3.76E-01 | 0.12 | 1.13E-47 | 1.64 | 0.3826 | 0.12 |
| TGs in CMs and extremely large VLDL | 0.04 | 0.09 | 0.05 | 0.09 | 7.42E-01 | -0.05 | 0.27 | 0.31 | 0.23 | 0.20 | 1.86E-01 | 0.18 | 0.02 | 0.02 | 0.02 | 0.02 | 4.96E-01 | 0.09 | 2.69E-29 | 1.15 | 0.0886 | 0.24 |
| Conc. of very large VLDL particles | 0.00 | 0.00 | 0.00 | 0.00 | 9.02E-01 | -0.02 | 0.00 | 0.00 | 0.00 | 0.00 | 7.39E-01 | 0.04 | 0.00 | 0.00 | 0.00 | 0.00 | 3.47E-01 | 0.13 | 5.38E-58 | 1.71 | 0.4600 | 0.10 |
| Total lipids in very large VLDL | 0.12 | 0.11 | 0.13 | 0.12 | 8.95E-01 | -0.02 | 0.42 | 0.29 | 0.40 | 0.21 | 5.93E-01 | 0.07 | 0.04 | 0.03 | 0.04 | 0.03 | 3.57E-01 | 0.13 | 1.63E-51 | 1.53 | 0.3391 | 0.13 |
| Phospholipids in very large VLDL | 0.02 | 0.02 | 0.02 | 0.02 | 8.84E-01 | -0.02 | 0.08 | 0.06 | 0.08 | 0.04 | 7.82E-01 | 0.04 | 0.01 | 0.01 | 0.01 | 0.01 | 3.25E-01 | 0.13 | 1.01E-57 | 1.73 | 0.4920 | 0.10 |
| Cholesterol in very large VLDL | 0.03 | 0.02 | 0.03 | 0.02 | 9.82E-01 | 0.00 | 0.10 | 0.05 | 0.10 | 0.04 | 6.36E-01 | -0.06 | 0.01 | 0.01 | 0.01 | 0.01 | 2.18E-01 | 0.17 | 5.84E-69 | 1.99 | 0.8769 | -0.02 |
| CEs in very large VLDL | 0.02 | 0.01 | 0.02 | 0.01 | 9.46E-01 | 0.01 | 0.05 | 0.03 | 0.06 | 0.02 | 2.82E-01 | -0.14 | 0.01 | 0.01 | 0.01 | 0.01 | 1.76E-01 | 0.18 | 6.55E-74 | 2.10 | 0.3905 | -0.12 |
| Free cholesterol in very large VLDL | 0.01 | 0.01 | 0.01 | 0.01 | 9.09E-01 | -0.02 | 0.05 | 0.03 | 0.05 | 0.02 | 9.24E-01 | 0.01 | 0.00 | 0.00 | 0.00 | 0.00 | 2.89E-01 | 0.14 | 4.33E-61 | 1.81 | 0.6222 | 0.07 |
| TGs in very large VLDL | 0.07 | 0.07 | 0.08 | 0.08 | 8.75E-01 | -0.02 | 0.24 | 0.19 | 0.22 | 0.13 | 3.63E-01 | 0.12 | 0.02 | 0.02 | 0.02 | 0.02 | 4.74E-01 | 0.10 | 7.33E-42 | 1.30 | 0.1772 | 0.19 |
| Conc. of large VLDL particles | 0.00 | 0.00 | 0.00 | 0.00 | 9.98E-01 | 0.00 | 0.00 | 0.00 | 0.00 | 0.00 | 9.42E-01 | 0.01 | 0.00 | 0.00 | 0.00 | 0.00 | 2.91E-01 | 0.14 | 1.05E-58 | 1.71 | 0.6556 | 0.06 |
| Total lipids in large VLDL | 0.24 | 0.16 | 0.24 | 0.17 | 1.00E+00 | 0.00 | 0.63 | 0.39 | 0.62 | 0.31 | 7.42E-01 | 0.04 | 0.08 | 0.05 | 0.08 | 0.05 | 3.30E-01 | 0.13 | 1.25E-49 | 1.48 | 0.4592 | 0.10 |
| Phospholipids in large VLDL | 0.05 | 0.03 | 0.05 | 0.03 | 9.23E-01 | -0.01 | 0.14 | 0.08 | 0.14 | 0.07 | 9.12E-01 | -0.01 | 0.02 | 0.01 | 0.01 | 0.01 | 3.19E-01 | 0.14 | 1.12E-61 | 1.79 | 0.7840 | 0.04 |
| Cholesterol in large VLDL | 0.06 | 0.04 | 0.06 | 0.04 | 9.50E-01 | -0.01 | 0.18 | 0.09 | 0.19 | 0.08 | 4.27E-01 | -0.10 | 0.02 | 0.02 | 0.02 | 0.02 | 2.70E-01 | 0.15 | 9.44E-69 | 1.96 | 0.6440 | -0.06 |
| CEs in large VLDL | 0.03 | 0.02 | 0.03 | 0.02 | 9.40E-01 | -0.01 | 0.09 | 0.04 | 0.10 | 0.04 | 1.61E-01 | -0.18 | 0.01 | 0.01 | 0.01 | 0.01 | 2.53E-01 | 0.15 | 1.94E-72 | 2.08 | 0.2698 | -0.15 |
| Free cholesterol in large VLDL | 0.03 | 0.02 | 0.03 | 0.02 | 9.61E-01 | -0.01 | 0.09 | 0.05 | 0.09 | 0.04 | 8.26E-01 | -0.03 | 0.01 | 0.01 | 0.01 | 0.01 | 2.96E-01 | 0.14 | 5.41E-63 | 1.81 | 0.8812 | 0.02 |
| TGs in large VLDL | 0.14 | 0.09 | 0.13 | 0.10 | 9.51E-01 | 0.01 | 0.31 | 0.22 | 0.28 | 0.17 | 3.36E-01 | 0.13 | 0.04 | 0.03 | 0.04 | 0.02 | 4.13E-01 | 0.11 | 2.10E-33 | 1.10 | 0.1666 | 0.19 |
| Conc. of medium VLDL particles | 0.00 | 0.00 | 0.00 | 0.00 | 8.14E-01 | 0.03 | 0.00 | 0.00 | 0.00 | 0.00 | 1.17E-01 | -0.20 | 0.00 | 0.00 | 0.00 | 0.00 | 1.58E-01 | 0.19 | 8.55E-81 | 2.29 | 0.1464 | -0.20 |
| Total lipids in medium VLDL | 0.46 | 0.17 | 0.45 | 0.20 | 8.35E-01 | 0.03 | 1.01 | 0.38 | 1.06 | 0.35 | 2.60E-01 | -0.15 | 0.14 | 0.07 | 0.13 | 0.07 | 1.92E-01 | 0.18 | 2.56E-74 | 2.08 | 0.3537 | -0.13 |
| Phospholipids in medium VLDL | 0.10 | 0.04 | 0.10 | 0.04 | 8.24E-01 | 0.03 | 0.23 | 0.08 | 0.25 | 0.08 | 8.13E-02 | -0.23 | 0.02 | 0.02 | 0.02 | 0.02 | 1.58E-01 | 0.19 | 4.42E-83 | 2.36 | 0.0980 | -0.23 |
| Cholesterol in medium VLDL | 0.12 | 0.04 | 0.12 | 0.05 | 7.65E-01 | 0.04 | 0.26 | 0.10 | 0.30 | 0.10 | 1.24E-02 | -0.33 | 0.03 | 0.02 | 0.03 | 0.02 | 1.38E-01 | 0.20 | 8.84E-76 | 2.20 | 0.0099 | -0.36 |
| CEs in medium VLDL | 0.06 | 0.02 | 0.06 | 0.03 | 7.50E-01 | 0.04 | 0.13 | 0.05 | 0.15 | 0.06 | 7.14E-03 | -0.35 | 0.02 | 0.01 | 0.01 | 0.01 | 1.51E-01 | 0.19 | 7.54E-63 | 1.86 | 0.0043 | -0.39 |
| Free cholesterol in medium VLDL | 0.06 | 0.02 | 0.06 | 0.03 | 8.03E-01 | 0.03 | 0.14 | 0.05 | 0.15 | 0.05 | 4.35E-02 | -0.26 | 0.02 | 0.01 | 0.01 | 0.01 | 1.53E-01 | 0.19 | 3.61E-82 | 2.37 | 0.0490 | -0.27 |
| TGs in medium VLDL | 0.24 | 0.11 | 0.24 | 0.12 | 8.93E-01 | 0.02 | 0.51 | 0.25 | 0.52 | 0.21 | 9.06E-01 | -0.02 | 0.08 | 0.04 | 0.08 | 0.04 | 3.37E-01 | 0.13 | 1.80E-54 | 1.59 | 0.8230 | 0.03 |
| Conc. of small VLDL particles | 0.00 | 0.00 | 0.00 | 0.00 | 9.72E-01 | 0.00 | 0.00 | 0.00 | 0.00 | 0.00 | 2.24E-02 | -0.30 | 0.00 | 0.00 | 0.00 | 0.00 | 3.16E-01 | 0.14 | 7.30E-88 | 2.69 | 0.0322 | -0.29 |
| Total lipids in small VLDL | 0.36 | 0.11 | 0.36 | 0.12 | 9.72E-01 | 0.00 | 0.81 | 0.25 | 0.89 | 0.25 | 1.93E-02 | -0.31 | 0.16 | 0.07 | 0.15 | 0.07 | 3.02E-01 | 0.14 | 1.33E-88 | 2.69 | 0.0272 | -0.30 |
| Phospholipids in small VLDL | 0.08 | 0.02 | 0.08 | 0.03 | 9.13E-01 | 0.01 | 0.18 | 0.05 | 0.20 | 0.06 | 9.55E-03 | -0.34 | 0.03 | 0.01 | 0.03 | 0.01 | 2.16E-01 | 0.17 | 1.00E-87 | 2.64 | 0.0117 | -0.35 |
| Cholesterol in small VLDL | 0.117 | 0.039 | 0.116 | 0.044 | 9.72E-01 | 0.00 | 0.282 | 0.090 | 0.318 | 0.093 | 2.54E-03 | -0.40 | 0.06 | 0.02 | 0.05 | 0.02 | 2.03E-01 | 0.17 | 1.83E-86 | 2.72 | 0.0031 | -0.41 |
| CEs in small VLDL | 0.071 | 0.025 | 0.071 | 0.027 | 9.42E-01 | -0.01 | 0.181 | 0.059 | 0.206 | 0.060 | 1.59E-03 | -0.42 | 0.04 | 0.02 | 0.04 | 0.02 | 2.19E-01 | 0.17 | 3.12E-87 | 2.80 | 0.0022 | -0.42 |
| Free cholesterol in small VLDL | 0.05 | 0.01 | 0.05 | 0.02 | 8.25E-01 | 0.03 | 0.10 | 0.03 | 0.11 | 0.03 | 6.62E-03 | -0.36 | 0.02 | 0.01 | 0.02 | 0.01 | 1.82E-01 | 0.18 | 7.74E-84 | 2.52 | 0.0065 | -0.37 |
| TGs in small VLDL | 0.16 | 0.05 | 0.16 | 0.06 | 9.97E-01 | 0.00 | 0.36 | 0.13 | 0.38 | 0.12 | 1.73E-01 | -0.18 | 0.07 | 0.03 | 0.07 | 0.03 | 4.86E-01 | 0.09 | 1.87E-77 | 2.31 | 0.2623 | -0.15 |
| Conc. of very small VLDL particles | 0.00 | 0.00 | 0.00 | 0.00 | 8.01E-01 | 0.03 | 0.00 | 0.00 | 0.00 | 0.00 | 3.52E-03 | -0.38 | 0.00 | 0.00 | 0.00 | 0.00 | 2.46E-01 | 0.16 | 1.21E-95 | 3.12 | 0.0041 | -0.40 |
| Total lipids in very small VLDL | 0.35 | 0.08 | 0.34 | 0.09 | 9.05E-01 | 0.02 | 0.72 | 0.18 | 0.79 | 0.17 | 3.13E-03 | -0.39 | 0.20 | 0.04 | 0.20 | 0.04 | 2.63E-01 | 0.15 | 4.09E-96 | 3.12 | 0.0048 | -0.39 |
| Phospholipids in very small VLDL | 0.099 | 0.026 | 0.099 | 0.028 | 9.63E-01 | -0.01 | 0.220 | 0.057 | 0.242 | 0.054 | 2.96E-03 | -0.39 | 0.065 | 0.013 | 0.063 | 0.013 | 2.56E-01 | 0.15 | 4.45E-96 | 3.15 | 0.0051 | -0.39 |
| Cholesterol in very small VLDL | 0.17 | 0.04 | 0.17 | 0.05 | 7.53E-01 | 0.04 | 0.33 | 0.08 | 0.36 | 0.08 | 4.43E-03 | -0.37 | 0.10 | 0.02 | 0.09 | 0.02 | 2.84E-01 | 0.15 | 2.27E-86 | 2.74 | 0.0050 | -0.39 |
| CEs in very small VLDL | 0.12 | 0.03 | 0.12 | 0.03 | 6.93E-01 | 0.05 | 0.21 | 0.06 | 0.23 | 0.05 | 7.54E-03 | -0.35 | 0.06 | 0.01 | 0.06 | 0.02 | 3.39E-01 | 0.13 | 2.08E-79 | 2.48 | 0.0070 | -0.37 |
| Free cholesterol in very small VLDL | 0.053 | 0.014 | 0.053 | 0.015 | 8.93E-01 | 0.02 | 0.115 | 0.030 | 0.126 | 0.028 | 2.75E-03 | -0.39 | 0.03 | 0.01 | 0.03 | 0.01 | 2.48E-01 | 0.16 | 1.39E-94 | 3.09 | 0.0040 | -0.40 |
| TGs in very small VLDL | 0.08 | 0.02 | 0.08 | 0.02 | 9.26E-01 | -0.01 | 0.18 | 0.05 | 0.19 | 0.05 | 1.62E-02 | -0.32 | 0.04 | 0.01 | 0.04 | 0.01 | 4.62E-01 | 0.10 | 8.86E-96 | 3.13 | 0.0237 | -0.31 |
| Conc. of IDL particles | 0.00 | 0.00 | 0.00 | 0.00 | 4.32E-01 | 0.11 | 0.00 | 0.00 | 0.00 | 0.00 | 1.49E-02 | -0.32 | 0.00 | 0.00 | 0.00 | 0.00 | 2.74E-01 | 0.15 | 1.36E-77 | 2.29 | 0.0075 | -0.37 |
| Total lipids in IDL | 1.26 | 0.23 | 1.23 | 0.28 | 3.83E-01 | 0.12 | 1.98 | 0.45 | 2.12 | 0.46 | 2.80E-02 | -0.29 | 0.53 | 0.11 | 0.52 | 0.13 | 4.36E-01 | 0.11 | 9.50E-75 | 2.25 | 0.0139 | -0.34 |
| Phospholipids in IDL | 0.28 | 0.05 | 0.28 | 0.06 | 5.31E-01 | 0.09 | 0.46 | 0.11 | 0.49 | 0.11 | 2.07E-02 | -0.30 | 0.14 | 0.03 | 0.14 | 0.03 | 3.56E-01 | 0.13 | 1.89E-74 | 2.30 | 0.0138 | -0.34 |
| Cholesterol in IDL | 0.86 | 0.16 | 0.83 | 0.20 | 3.04E-01 | 0.14 | 1.29 | 0.31 | 1.37 | 0.33 | 5.52E-02 | -0.25 | 0.32 | 0.09 | 0.32 | 0.10 | 5.28E-01 | 0.09 | 4.15E-64 | 1.90 | 0.0214 | -0.32 |
| CEs in IDL | 0.65 | 0.12 | 0.62 | 0.15 | 2.80E-01 | 0.15 | 0.96 | 0.23 | 1.02 | 0.24 | 6.29E-02 | -0.24 | 0.23 | 0.07 | 0.22 | 0.07 | 5.45E-01 | 0.08 | 7.73E-63 | 1.87 | 0.0238 | -0.31 |
| Free cholesterol in IDL | 0.21 | 0.04 | 0.21 | 0.05 | 4.05E-01 | 0.11 | 0.32 | 0.08 | 0.35 | 0.08 | 3.84E-02 | -0.27 | 0.10 | 0.02 | 0.09 | 0.03 | 4.87E-01 | 0.09 | 6.11E-67 | 1.97 | 0.0167 | -0.33 |

Supplementary table 1: NMR metabolomics results

| | Case-control comparisons | | | | | | | | | | | | | | | | | | Change during pregnancy | | | |
|---|---|---|---|---|---|---|---|---|---|---|---|---|---|---|---|---|---|---|---|---|---|---|
| | 1st trimester | | | | | | Delivery | | | | | | Cord serum | | | | | | Delivery vs. 1st trimester | | | |
| | LT4 | | Control | | | | LT4 | | Control | | | | LT4 | | Control | | | | All subjects | | LT4 vs. controls | |
| Metabolite abbreviation | Mean | SD | Mean | SD | p | d | Mean | SD | Mean | SD | p | d | Mean | SD | Mean | SD | p | d | p | d | p | d |
| TGs in IDL | 0.11 | 0.03 | 0.11 | 0.03 | 9.98E-01 | 0.00 | 0.24 | 0.06 | 0.26 | 0.06 | 1.33E-02 | -0.32 | 0.06 | 0.02 | 0.06 | 0.02 | 4.77E-01 | 0.10 | 3.56E-98 | 3.27 | 0.0194 | -0.32 |
| Conc. of large LDL particles | 0.00 | 0.00 | 0.00 | 0.00 | 8.32E-01 | 0.03 | 0.00 | 0.00 | 0.00 | 0.00 | 1.58E-02 | -0.32 | 0.00 | 0.00 | 0.00 | 0.00 | 1.59E-01 | 0.19 | 3.43E-77 | 2.26 | 0.0156 | -0.33 |
| Total lipids in large LDL | 1.57 | 0.29 | 1.54 | 0.36 | 4.04E-01 | 0.11 | 2.43 | 0.53 | 2.56 | 0.57 | 6.31E-02 | -0.24 | 0.56 | 0.15 | 0.54 | 0.17 | 3.14E-01 | 0.14 | 1.08E-71 | 2.13 | 0.0330 | -0.29 |
| Phospholipids in large LDL | 0.34 | 0.06 | 0.33 | 0.07 | 3.48E-01 | 0.13 | 0.50 | 0.11 | 0.53 | 0.12 | 4.68E-02 | -0.26 | 0.14 | 0.03 | 0.14 | 0.03 | 3.45E-01 | 0.13 | 8.54E-67 | 1.99 | 0.0218 | -0.32 |
| Cholesterol in large LDL | 1.13 | 0.21 | 1.10 | 0.27 | 4.00E-01 | 0.12 | 1.68 | 0.39 | 1.77 | 0.42 | 1.05E-01 | -0.21 | 0.37 | 0.11 | 0.35 | 0.13 | 3.17E-01 | 0.14 | 7.90E-64 | 1.89 | 0.0550 | -0.26 |
| CEs in large LDL | 0.83 | 0.16 | 0.81 | 0.20 | 4.45E-01 | 0.10 | 1.26 | 0.29 | 1.33 | 0.31 | 1.04E-01 | -0.21 | 0.26 | 0.08 | 0.25 | 0.09 | 2.94E-01 | 0.14 | 1.90E-67 | 1.98 | 0.0604 | -0.26 |
| Free cholesterol in large LDL | 0.30 | 0.05 | 0.29 | 0.07 | 2.96E-01 | 0.14 | 0.42 | 0.11 | 0.44 | 0.12 | 1.15E-01 | -0.21 | 0.10 | 0.03 | 0.10 | 0.04 | 4.00E-01 | 0.11 | 5.54E-52 | 1.61 | 0.0464 | -0.27 |
| TGs in large LDL | 0.11 | 0.02 | 0.11 | 0.03 | 8.42E-01 | 0.03 | 0.24 | 0.06 | 0.26 | 0.05 | 1.85E-02 | -0.31 | 0.06 | 0.02 | 0.06 | 0.02 | 5.29E-01 | 0.09 | 4.99E-98 | 3.32 | 0.0195 | -0.32 |
| Conc. of medium LDL particles | 0.00 | 0.00 | 0.00 | 0.00 | 4.65E-01 | 0.10 | 0.00 | 0.00 | 0.00 | 0.00 | 8.59E-02 | -0.22 | 0.00 | 0.00 | 0.00 | 0.00 | 1.33E-01 | 0.20 | 6.55E-80 | 2.31 | 0.0507 | -0.27 |
| Total lipids in medium LDL | 0.55 | 0.13 | 0.54 | 0.15 | 4.57E-01 | 0.10 | 0.93 | 0.23 | 0.99 | 0.25 | 7.80E-02 | -0.23 | 0.19 | 0.06 | 0.18 | 0.06 | 1.79E-01 | 0.18 | 6.40E-79 | 2.23 | 0.0429 | -0.28 |
| Phospholipids in medium LDL | 0.15 | 0.03 | 0.14 | 0.04 | 4.48E-01 | 0.10 | 0.24 | 0.06 | 0.25 | 0.06 | 5.38E-02 | -0.25 | 0.05 | 0.01 | 0.05 | 0.02 | 2.13E-01 | 0.17 | 1.28E-74 | 2.11 | 0.0272 | -0.30 |
| Cholesterol in medium LDL | 0.37 | 0.09 | 0.36 | 0.10 | 4.48E-01 | 0.10 | 0.61 | 0.16 | 0.65 | 0.17 | 1.14E-01 | -0.21 | 0.12 | 0.04 | 0.11 | 0.04 | 1.57E-01 | 0.19 | 3.42E-73 | 2.04 | 0.0648 | -0.25 |
| CEs in medium LDL | 0.26 | 0.07 | 0.25 | 0.08 | 4.89E-01 | 0.09 | 0.46 | 0.12 | 0.48 | 0.13 | 1.41E-01 | -0.19 | 0.08 | 0.03 | 0.07 | 0.03 | 1.37E-01 | 0.20 | 5.01E-78 | 2.16 | 0.0923 | -0.23 |
| Free cholesterol in medium LDL | 0.11 | 0.02 | 0.11 | 0.03 | 3.70E-01 | 0.12 | 0.16 | 0.04 | 0.17 | 0.05 | 8.43E-02 | -0.23 | 0.04 | 0.01 | 0.04 | 0.01 | 2.69E-01 | 0.15 | 1.54E-51 | 1.53 | 0.0350 | -0.29 |
| TGs in medium LDL | 0.04 | 0.01 | 0.04 | 0.01 | 8.05E-01 | 0.03 | 0.08 | 0.02 | 0.09 | 0.02 | 4.66E-02 | -0.26 | 0.02 | 0.01 | 0.02 | 0.01 | 4.93E-01 | 0.09 | 9.23E-99 | 3.30 | 0.0438 | -0.28 |
| Conc. of small LDL particles | 0.00 | 0.00 | 0.00 | 0.00 | 6.37E-01 | 0.06 | 0.00 | 0.00 | 0.00 | 0.00 | 8.51E-02 | -0.23 | 0.00 | 0.00 | 0.00 | 0.00 | 1.39E-01 | 0.20 | 2.30E-84 | 2.44 | 0.0696 | -0.25 |
| Total lipids in small LDL | 0.25 | 0.05 | 0.24 | 0.05 | 5.73E-01 | 0.08 | 0.41 | 0.10 | 0.43 | 0.10 | 7.06E-02 | -0.24 | 0.13 | 0.02 | 0.13 | 0.02 | 1.47E-01 | 0.20 | 1.82E-80 | 2.26 | 0.0437 | -0.28 |
| Phospholipids in small LDL | 0.08 | 0.01 | 0.08 | 0.02 | 6.81E-01 | 0.06 | 0.12 | 0.03 | 0.13 | 0.03 | 4.07E-02 | -0.27 | 0.05 | 0.01 | 0.05 | 0.01 | 2.18E-01 | 0.17 | 1.14E-78 | 2.20 | 0.0227 | -0.31 |
| Cholesterol in small LDL | 0.15 | 0.03 | 0.15 | 0.04 | 5.03E-01 | 0.09 | 0.25 | 0.06 | 0.26 | 0.07 | 9.61E-02 | -0.22 | 0.07 | 0.01 | 0.07 | 0.02 | 1.36E-01 | 0.20 | 6.38E-73 | 2.04 | 0.0577 | -0.26 |
| CEs in small LDL | 0.11 | 0.02 | 0.11 | 0.03 | 5.05E-01 | 0.09 | 0.19 | 0.05 | 0.20 | 0.05 | 1.11E-01 | -0.21 | 0.05 | 0.01 | 0.05 | 0.01 | 1.34E-01 | 0.20 | 2.32E-78 | 2.23 | 0.0768 | -0.24 |
| Free cholesterol in small LDL | 0.04 | 0.01 | 0.04 | 0.01 | 5.32E-01 | 0.09 | 0.06 | 0.02 | 0.06 | 0.02 | 9.26E-02 | -0.22 | 0.02 | 0.00 | 0.02 | 0.00 | 1.81E-01 | 0.18 | 8.60E-47 | 1.34 | 0.0420 | -0.28 |
| TGs in small LDL | 0.01 | 0.00 | 0.01 | 0.00 | 9.63E-01 | -0.01 | 0.04 | 0.01 | 0.04 | 0.01 | 2.36E-01 | -0.16 | 0.01 | 0.00 | 0.01 | 0.00 | 4.39E-01 | 0.10 | 1.24E-90 | 2.85 | 0.3084 | -0.14 |
| Conc. of very large HDL particles | 0.00 | 0.00 | 0.00 | 0.00 | 6.29E-01 | 0.07 | 0.00 | 0.00 | 0.00 | 0.00 | 2.51E-01 | -0.15 | 0.00 | 0.00 | 0.00 | 0.00 | 7.05E-01 | -0.05 | 3.28E-52 | 1.35 | 0.1140 | -0.22 |
| Total lipids in very large HDL | 0.26 | 0.10 | 0.25 | 0.09 | 6.82E-01 | 0.06 | 0.35 | 0.12 | 0.36 | 0.13 | 3.38E-01 | -0.13 | 0.16 | 0.05 | 0.16 | 0.05 | 6.54E-01 | -0.06 | 6.50E-36 | 0.95 | 0.1578 | -0.19 |
| Phospholipids in very large HDL | 0.13 | 0.05 | 0.13 | 0.05 | 7.05E-01 | 0.05 | 0.17 | 0.06 | 0.18 | 0.07 | 3.95E-01 | -0.11 | 0.08 | 0.03 | 0.08 | 0.03 | 6.41E-01 | -0.06 | 8.93E-31 | 0.84 | 0.2008 | -0.17 |
| Cholesterol in very large HDL | 0.12 | 0.04 | 0.12 | 0.04 | 6.49E-01 | 0.06 | 0.15 | 0.05 | 0.16 | 0.06 | 3.60E-01 | -0.12 | 0.08 | 0.02 | 0.08 | 0.02 | 6.48E-01 | -0.06 | 2.07E-30 | 0.83 | 0.1509 | -0.20 |
| CEs in very large HDL | 0.09 | 0.03 | 0.09 | 0.03 | 6.17E-01 | 0.07 | 0.11 | 0.04 | 0.12 | 0.04 | 3.74E-01 | -0.12 | 0.05 | 0.02 | 0.05 | 0.02 | 6.65E-01 | -0.06 | 1.23E-28 | 0.78 | 0.1492 | -0.20 |
| Free cholesterol in very large HDL | 0.03 | 0.01 | 0.03 | 0.01 | 7.80E-01 | 0.04 | 0.04 | 0.01 | 0.04 | 0.01 | 3.33E-01 | -0.13 | 0.03 | 0.01 | 0.03 | 0.01 | 6.03E-01 | -0.07 | 8.08E-34 | 0.97 | 0.1764 | -0.18 |
| TGs in very large HDL | 0.01 | 0.00 | 0.01 | 0.00 | 8.73E-01 | 0.02 | 0.02 | 0.01 | 0.02 | 0.01 | 5.81E-02 | -0.25 | 0.00 | 0.00 | 0.00 | 0.00 | 6.81E-01 | 0.06 | 4.80E-91 | 2.87 | 0.0710 | -0.25 |
| Conc. of large HDL particles | 0.00 | 0.00 | 0.00 | 0.00 | 4.58E-01 | 0.10 | 0.00 | 0.00 | 0.00 | 0.00 | 7.86E-01 | -0.04 | 0.00 | 0.00 | 0.00 | 0.00 | 6.92E-01 | -0.05 | 8.96E-23 | 0.71 | 0.4056 | -0.11 |
| Total lipids in large HDL | 1.13 | 0.34 | 1.09 | 0.33 | 4.62E-01 | 0.10 | 1.33 | 0.43 | 1.33 | 0.46 | 9.42E-01 | -0.01 | 0.40 | 0.18 | 0.41 | 0.18 | 6.64E-01 | -0.06 | 8.16E-16 | 0.57 | 0.5323 | -0.08 |
| Phospholipids in large HDL | 0.55 | 0.16 | 0.54 | 0.15 | 4.30E-01 | 0.11 | 0.63 | 0.20 | 0.63 | 0.21 | 8.61E-01 | 0.02 | 0.20 | 0.08 | 0.20 | 0.08 | 6.37E-01 | -0.06 | 1.18E-11 | 0.48 | 0.6895 | -0.05 |
| Cholesterol in large HDL | 0.52 | 0.18 | 0.51 | 0.17 | 4.92E-01 | 0.09 | 0.60 | 0.22 | 0.60 | 0.23 | 9.04E-01 | -0.02 | 0.19 | 0.09 | 0.19 | 0.10 | 6.67E-01 | -0.06 | 1.59E-10 | 0.42 | 0.5047 | -0.09 |
| CEs in large HDL | 0.40 | 0.14 | 0.39 | 0.13 | 4.93E-01 | 0.09 | 0.45 | 0.17 | 0.45 | 0.18 | 9.23E-01 | -0.01 | 0.15 | 0.07 | 0.15 | 0.07 | 6.56E-01 | -0.06 | 1.03E-07 | 0.34 | 0.5225 | -0.09 |
| Free cholesterol in large HDL | 0.12 | 0.04 | 0.12 | 0.04 | 4.95E-01 | 0.09 | 0.15 | 0.05 | 0.15 | 0.05 | 8.41E-01 | -0.03 | 0.04 | 0.02 | 0.04 | 0.02 | 7.03E-01 | -0.05 | 2.60E-21 | 0.68 | 0.4537 | -0.10 |
| TGs in large HDL | 0.05 | 0.02 | 0.05 | 0.02 | 6.88E-01 | 0.05 | 0.10 | 0.03 | 0.10 | 0.03 | 1.19E-01 | -0.20 | 0.01 | 0.01 | 0.01 | 0.01 | 7.94E-01 | 0.04 | 4.13E-77 | 2.37 | 0.1067 | -0.22 |
| Conc. of medium HDL particles | 0.01 | 0.00 | 0.01 | 0.00 | 2.32E-01 | 0.16 | 0.01 | 0.00 | 0.01 | 0.00 | 3.31E-01 | 0.13 | 0.00 | 0.00 | 0.00 | 0.00 | 8.79E-01 | -0.02 | 4.14E-07 | 0.39 | 0.8425 | 0.03 |
| Total lipids in medium HDL | 1.35 | 0.20 | 1.32 | 0.23 | 2.59E-01 | 0.15 | 1.46 | 0.29 | 1.42 | 0.30 | 3.06E-01 | 0.13 | 0.54 | 0.10 | 0.54 | 0.10 | 8.34E-01 | -0.03 | 5.84E-07 | 0.39 | 0.7911 | 0.04 |
| Phospholipids in medium HDL | 0.62 | 0.09 | 0.61 | 0.10 | 3.04E-01 | 0.14 | 0.67 | 0.13 | 0.65 | 0.13 | 2.18E-01 | 0.16 | 0.26 | 0.04 | 0.26 | 0.04 | 8.88E-01 | -0.02 | 4.92E-08 | 0.44 | 0.6249 | 0.07 |
| Cholesterol in medium HDL | 0.67 | 0.10 | 0.65 | 0.12 | 2.03E-01 | 0.17 | 0.66 | 0.16 | 0.63 | 0.17 | 2.85E-01 | 0.14 | 0.25 | 0.06 | 0.25 | 0.06 | 7.18E-01 | -0.05 | 2.35E-01 | -0.09 | 0.7850 | 0.04 |
| CEs in medium HDL | 0.54 | 0.08 | 0.53 | 0.09 | 2.01E-01 | 0.18 | 0.51 | 0.13 | 0.49 | 0.13 | 2.53E-01 | 0.15 | 0.22 | 0.05 | 0.22 | 0.05 | 6.86E-01 | -0.05 | 3.45E-04 | -0.27 | 0.7245 | 0.05 |
| Free cholesterol in medium HDL | 0.12 | 0.02 | 0.12 | 0.03 | 2.27E-01 | 0.17 | 0.14 | 0.03 | 0.14 | 0.03 | 4.79E-01 | 0.09 | 0.03 | 0.01 | 0.03 | 0.01 | 8.45E-01 | -0.03 | 3.13E-15 | 0.64 | 0.9499 | -0.01 |
| TGs in medium HDL | 0.07 | 0.02 | 0.07 | 0.02 | 8.27E-01 | 0.03 | 0.13 | 0.03 | 0.13 | 0.03 | 2.49E-01 | -0.15 | 0.02 | 0.01 | 0.02 | 0.01 | 4.53E-01 | 0.10 | 1.26E-85 | 2.53 | 0.2863 | -0.15 |
| Conc. of small HDL particles | 0.01 | 0.00 | 0.01 | 0.00 | 2.49E-01 | 0.16 | 0.01 | 0.00 | 0.01 | 0.00 | 4.95E-01 | 0.09 | 0.01 | 0.00 | 0.01 | 0.00 | 5.59E-01 | 0.08 | 1.46E-08 | 0.41 | 0.9815 | 0.00 |
| Total lipids in small HDL | 1.25 | 0.15 | 1.23 | 0.17 | 3.28E-01 | 0.13 | 1.39 | 0.20 | 1.38 | 0.19 | 4.58E-01 | 0.10 | 0.68 | 0.08 | 0.67 | 0.08 | 5.65E-01 | 0.08 | 3.70E-26 | 0.85 | 0.9003 | 0.02 |
| Phospholipids in small HDL | 0.72 | 0.09 | 0.71 | 0.11 | 3.53E-01 | 0.13 | 0.81 | 0.12 | 0.79 | 0.11 | 3.07E-01 | 0.13 | 0.39 | 0.04 | 0.39 | 0.04 | 5.77E-01 | 0.08 | 3.65E-23 | 0.80 | 0.7059 | 0.05 |
| Cholesterol in small HDL | 0.47 | 0.05 | 0.46 | 0.06 | 2.37E-01 | 0.16 | 0.48 | 0.08 | 0.47 | 0.08 | 4.36E-01 | 0.10 | 0.25 | 0.03 | 0.25 | 0.03 | 6.53E-01 | 0.06 | 2.22E-01 | 0.08 | 0.9398 | 0.01 |
| CEs in small HDL | 0.34 | 0.04 | 0.33 | 0.05 | 2.89E-01 | 0.15 | 0.31 | 0.07 | 0.30 | 0.07 | 2.49E-01 | 0.15 | 0.19 | 0.02 | 0.19 | 0.02 | 6.92E-01 | 0.05 | 7.67E-14 | -0.58 | 0.6168 | 0.07 |
| Free cholesterol in small HDL | 0.13 | 0.01 | 0.13 | 0.02 | 2.35E-01 | 0.16 | 0.17 | 0.02 | 0.17 | 0.02 | 4.09E-01 | -0.11 | 0.06 | 0.01 | 0.06 | 0.01 | 5.96E-01 | 0.07 | 1.40E-71 | 2.08 | 0.2348 | -0.16 |
| TGs in small HDL | 0.05 | 0.02 | 0.05 | 0.02 | 9.50E-01 | -0.01 | 0.11 | 0.03 | 0.11 | 0.03 | 1.78E-01 | -0.18 | 0.03 | 0.01 | 0.03 | 0.01 | 4.32E-01 | 0.11 | 1.61E-92 | 2.66 | 0.2330 | -0.16 |
| Phospholipids to total lipids ratio in CMs and extremely large VLDL | 14.01 | 6.57 | 11.83 | 7.45 | 8.77E-02 | 0.31 | 16.73 | 4.20 | 17.44 | 4.56 | 2.19E-01 | -0.16 | 13.95 | 9.41 | 13.70 | 10.07 | 8.64E-01 | 0.03 | 2.16E-07 | 0.76 | 0.0086 | -0.48 |
| Cholesterol to total lipids ratio in CMs and extremely large VLDL | 40.35 | 24.14 | 36.53 | 24.32 | 3.88E-01 | 0.16 | 32.80 | 18.01 | 34.19 | 17.37 | 5.51E-01 | -0.08 | 27.49 | 15.04 | 26.01 | 17.85 | 5.56E-01 | 0.09 | 2.58E-03 | -0.22 | 0.1035 | -0.30 |
| CEs to total lipids ratio in CMs and extremely large VLDL | 26.36 | 18.04 | 24.02 | 19.40 | 4.91E-01 | 0.12 | 20.48 | 14.15 | 21.28 | 12.97 | 6.57E-01 | -0.06 | 14.84 | 9.05 | 13.20 | 11.20 | 2.91E-01 | 0.16 | 9.37E-04 | -0.25 | 0.1781 | -0.24 |
| Free cholesterol to total lipids ratio in CMs and extremely large VLDL | 13.99 | 7.35 | 12.51 | 6.82 | 2.58E-01 | 0.21 | 12.32 | 4.53 | 12.91 | 4.74 | 3.30E-01 | -0.13 | 12.66 | 6.60 | 12.81 | 7.46 | 8.85E-01 | -0.02 | 1.09E-01 | -0.08 | 0.0383 | -0.38 |
| TGs to total lipids ratio in CMs and extremely large VLDL | 45.65 | 27.16 | 51.64 | 26.44 | 2.23E-01 | -0.22 | 50.47 | 20.61 | 48.37 | 20.36 | 4.37E-01 | 0.10 | 58.56 | 23.68 | 60.29 | 26.97 | 6.54E-01 | -0.07 | 2.28E-01 | 0.01 | 0.0175 | 0.43 |
| Phospholipids to total lipids ratio in very large VLDL | 14.57 | 4.58 | 14.78 | 4.41 | 7.38E-01 | -0.05 | 19.86 | 2.03 | 20.18 | 1.62 | 1.81E-01 | -0.18 | 14.36 | 5.06 | 13.29 | 6.15 | 1.77E-01 | 0.19 | 7.72E-40 | 1.69 | 0.9548 | -0.01 |
| Cholesterol to total lipids ratio in very large VLDL | 25.97 | 9.00 | 27.80 | 13.80 | 2.60E-01 | -0.16 | 26.87 | 7.52 | 27.32 | 6.01 | 6.12E-01 | -0.07 | 29.66 | 7.67 | 28.08 | 9.73 | 2.00E-01 | 0.18 | 6.40E-01 | 0.01 | 0.3228 | 0.14 |
| CEs to total lipids ratio in very large VLDL | 16.06 | 7.64 | 17.34 | 11.31 | 3.42E-01 | -0.14 | 15.07 | 5.77 | 15.36 | 4.55 | 6.69E-01 | -0.06 | 17.60 | 5.86 | 16.55 | 6.77 | 2.36E-01 | 0.17 | 4.91E-03 | -0.21 | 0.3800 | 0.12 |
| Free cholesterol to total lipids ratio in very large VLDL | 9.90 | 2.30 | 10.45 | 3.03 | 1.47E-01 | -0.21 | 11.81 | 1.88 | 11.97 | 1.55 | 4.72E-01 | -0.09 | 12.06 | 2.39 | 11.53 | 3.44 | 2.09E-01 | 0.18 | 5.57E-14 | 0.76 | 0.2590 | 0.16 |
| TGs to total lipids ratio in very large VLDL | 59.46 | 10.71 | 57.42 | 14.70 | 2.57E-01 | 0.16 | 53.27 | 9.10 | 52.50 | 7.34 | 4.74E-01 | 0.09 | 55.98 | 10.89 | 58.63 | 14.45 | 1.41E-01 | -0.21 | 3.57E-07 | -0.51 | 0.4484 | -0.11 |
| Phospholipids to total lipids ratio in large VLDL | 17.52 | 3.85 | 17.81 | 4.11 | 5.96E-01 | -0.07 | 23.21 | 3.15 | 23.75 | 2.86 | 1.63E-01 | -0.18 | 16.01 | 5.92 | 14.98 | 6.67 | 2.32E-01 | 0.16 | 1.12E-45 | 1.66 | 0.6031 | -0.07 |
| Cholesterol to total lipids ratio in large VLDL | 25.83 | 5.26 | 26.54 | 6.63 | 3.84E-01 | -0.12 | 31.31 | 6.81 | 32.57 | 6.16 | 1.35E-01 | -0.20 | 27.73 | 6.20 | 26.56 | 7.56 | 2.10E-01 | 0.17 | 1.01E-21 | 0.91 | 0.5543 | -0.08 |
| CEs to total lipids ratio in large VLDL | 13.56 | 3.56 | 14.02 | 4.66 | 4.22E-01 | -0.11 | 16.42 | 4.72 | 17.35 | 4.19 | 1.10E-01 | -0.21 | 14.71 | 4.08 | 13.95 | 4.94 | 2.12E-01 | 0.17 | 1.03E-15 | 0.71 | 0.4497 | -0.10 |
| Free cholesterol to total lipids ratio in large VLDL | 12.26 | 1.98 | 12.52 | 2.18 | 3.66E-01 | -0.12 | 14.89 | 2.15 | 15.22 | 2.05 | 2.18E-01 | -0.16 | 13.02 | 2.60 | 12.61 | 2.98 | 2.81E-01 | 0.15 | 8.70E-32 | 1.27 | 0.7825 | -0.04 |
| TGs to total lipids ratio in large VLDL | 56.65 | 8.42 | 55.65 | 9.87 | 4.25E-01 | 0.11 | 45.49 | 9.66 | 43.67 | 8.75 | 1.31E-01 | 0.20 | 56.26 | 11.20 | 58.46 | 13.45 | 1.90E-01 | -0.18 | 1.65E-32 | -1.25 | 0.4908 | 0.09 |

Supplementary table 1: NMR metabolomics results

| | Case-control comparisons | | | | | | | | | | | | | | | | | | Change during pregnancy | | | |
|---|---|---|---|---|---|---|---|---|---|---|---|---|---|---|---|---|---|---|---|---|---|---|
| | 1st trimester | | | | | | Delivery | | | | | | Cord serum | | | | | | Delivery vs. 1st trimester | | | |
| | LT4 | | Control | | | | LT4 | | Control | | | | LT4 | | Control | | | | All subjects | | LT4 vs. controls | |
| Metabolite abbreviation | Mean | SD | Mean | SD | p | d | Mean | SD | Mean | SD | p | d | Mean | SD | Mean | SD | p | d | p | d | p | d |
| Phospholipids to total lipids ratio in medium VLDL | 21.24 | 1.97 | 21.09 | 2.97 | 6.48E-01 | 0.06 | 23.25 | 2.20 | 23.61 | 1.83 | 1.73E-01 | -0.18 | 16.20 | 5.38 | 14.89 | 6.07 | 9.24E-02 | 0.23 | 7.29E-25 | 0.99 | 0.2447 | -0.16 |
| Cholesterol to total lipids ratio in medium VLDL | 26.28 | 5.64 | 26.08 | 6.76 | 8.11E-01 | 0.03 | 27.38 | 6.76 | 28.37 | 5.86 | 2.32E-01 | -0.16 | 20.88 | 8.82 | 18.96 | 9.85 | 1.30E-01 | 0.21 | 4.19E-04 | 0.27 | 0.1877 | -0.18 |
| CEs to total lipids ratio in medium VLDL | 13.59 | 4.18 | 13.49 | 4.88 | 8.64E-01 | 0.02 | 13.56 | 4.94 | 14.23 | 4.33 | 2.71E-01 | -0.14 | 11.02 | 5.80 | 9.91 | 6.37 | 1.79E-01 | 0.18 | 3.52E-01 | 0.08 | 0.2081 | -0.17 |
| Free cholesterol to total lipids ratio in medium VLDL | 12.69 | 1.53 | 12.59 | 2.00 | 6.92E-01 | 0.05 | 13.82 | 1.84 | 14.14 | 1.56 | 1.52E-01 | -0.19 | 9.86 | 3.40 | 9.05 | 3.84 | 1.00E-01 | 0.22 | 9.33E-19 | 0.77 | 0.1739 | -0.19 |
| TGs to total lipids ratio in medium VLDL | 52.47 | 7.44 | 52.83 | 9.43 | 7.57E-01 | -0.04 | 49.37 | 8.90 | 48.02 | 7.63 | 2.13E-01 | 0.16 | 62.93 | 13.64 | 66.15 | 15.38 | 1.02E-01 | -0.22 | 3.74E-09 | -0.47 | 0.1446 | 0.20 |
| Phospholipids to total lipids ratio in small VLDL | 22.25 | 1.53 | 22.14 | 1.85 | 6.41E-01 | 0.06 | 21.83 | 1.53 | 21.96 | 1.45 | 5.27E-01 | -0.08 | 20.47 | 1.90 | 20.10 | 1.86 | 1.46E-01 | 0.20 | 8.83E-03 | -0.18 | 0.6176 | -0.07 |
| Cholesterol to total lipids ratio in small VLDL | 32.50 | 4.18 | 32.62 | 4.19 | 8.35E-01 | -0.03 | 34.79 | 4.22 | 35.73 | 3.70 | 7.06E-02 | -0.24 | 36.42 | 3.98 | 35.88 | 4.03 | 3.22E-01 | 0.13 | 5.03E-16 | 0.66 | 0.2586 | -0.16 |
| CEs to total lipids ratio in small VLDL | 19.60 | 2.90 | 19.87 | 2.73 | 4.81E-01 | -0.10 | 22.30 | 2.78 | 23.09 | 2.33 | 1.77E-02 | -0.31 | 25.24 | 2.27 | 25.04 | 2.43 | 5.11E-01 | 0.09 | 4.12E-32 | 1.09 | 0.2476 | -0.16 |
| Free cholesterol to total lipids ratio in small VLDL | 12.90 | 1.56 | 12.75 | 1.84 | 5.09E-01 | 0.09 | 12.50 | 1.62 | 12.63 | 1.52 | 4.97E-01 | -0.09 | 11.17 | 2.10 | 10.84 | 2.04 | 2.42E-01 | 0.16 | 2.38E-02 | -0.15 | 0.3310 | -0.13 |
| TGs to total lipids ratio in small VLDL | 45.25 | 5.58 | 45.24 | 5.87 | 9.88E-01 | 0.00 | 43.38 | 5.65 | 42.32 | 5.07 | 1.30E-01 | 0.20 | 43.12 | 5.72 | 44.03 | 5.71 | 2.41E-01 | -0.16 | 1.57E-08 | -0.43 | 0.2123 | 0.17 |
| Phospholipids to total lipids ratio in very small VLDL | 28.44 | 1.11 | 28.68 | 1.04 | 1.02E-01 | -0.23 | 30.31 | 0.88 | 30.51 | 0.83 | 6.74E-02 | -0.24 | 32.09 | 1.02 | 32.13 | 1.00 | 7.59E-01 | -0.04 | 7.38E-65 | 1.90 | 0.8216 | 0.03 |
| Cholesterol to total lipids ratio in very small VLDL | 49.32 | 3.06 | 48.99 | 3.02 | 4.21E-01 | 0.11 | 45.33 | 3.84 | 45.40 | 3.47 | 8.95E-01 | -0.02 | 47.48 | 4.65 | 47.34 | 4.62 | 8.21E-01 | 0.03 | 5.36E-42 | -1.13 | 0.6415 | -0.06 |
| CEs to total lipids ratio in very small VLDL | 33.92 | 2.86 | 33.59 | 2.85 | 3.93E-01 | 0.12 | 29.52 | 3.64 | 29.48 | 3.28 | 9.19E-01 | 0.01 | 31.28 | 4.40 | 31.17 | 4.40 | 8.44E-01 | 0.03 | 2.46E-52 | -1.34 | 0.6403 | -0.06 |
| Free cholesterol to total lipids ratio in very small VLDL | 15.40 | 0.37 | 15.40 | 0.34 | 9.83E-01 | 0.00 | 15.81 | 0.37 | 15.92 | 0.31 | 1.48E-02 | -0.32 | 16.20 | 0.32 | 16.17 | 0.30 | 5.57E-01 | 0.08 | 1.65E-43 | 1.34 | 0.6990 | -0.05 |
| TGs to total lipids ratio in very small VLDL | 22.24 | 2.75 | 22.33 | 2.65 | 8.03E-01 | -0.03 | 24.36 | 3.32 | 24.09 | 2.86 | 5.08E-01 | 0.09 | 20.43 | 4.01 | 20.53 | 3.98 | 8.53E-01 | -0.02 | 1.02E-20 | 0.67 | 0.3478 | 0.13 |
| Phospholipids to total lipids ratio in IDL | 22.60 | 0.93 | 22.85 | 1.08 | 7.74E-02 | -0.24 | 22.93 | 0.76 | 23.08 | 0.80 | 1.54E-01 | -0.19 | 27.18 | 1.24 | 27.20 | 1.29 | 8.91E-01 | -0.02 | 6.28E-04 | 0.30 | 0.7884 | 0.04 |
| Cholesterol to total lipids ratio in IDL | 68.27 | 2.03 | 67.73 | 2.24 | 6.94E-02 | 0.25 | 64.52 | 3.19 | 64.30 | 2.58 | 5.48E-01 | 0.08 | 60.41 | 4.69 | 60.30 | 4.51 | 8.54E-01 | 0.02 | 1.60E-51 | -1.41 | 0.8737 | -0.02 |
| CEs to total lipids ratio in IDL | 51.35 | 1.91 | 50.78 | 2.18 | 4.33E-02 | 0.28 | 48.25 | 2.52 | 47.97 | 2.14 | 3.54E-01 | 0.12 | 42.39 | 4.23 | 42.27 | 4.08 | 8.21E-01 | 0.03 | 4.47E-45 | -1.33 | 0.8288 | -0.03 |
| Free cholesterol to total lipids ratio in IDL | 16.92 | 0.69 | 16.95 | 0.69 | 7.04E-01 | -0.05 | 16.27 | 0.84 | 16.33 | 0.61 | 5.64E-01 | -0.08 | 18.02 | 0.70 | 18.03 | 0.72 | 8.96E-01 | -0.02 | 3.75E-27 | -0.89 | 0.9764 | 0.00 |
| TGs to total lipids ratio in IDL | 9.13 | 1.52 | 9.42 | 1.65 | 1.84E-01 | -0.18 | 12.54 | 3.16 | 12.63 | 2.52 | 8.25E-01 | -0.03 | 12.41 | 3.74 | 12.50 | 3.63 | 8.54E-01 | -0.02 | 3.16E-50 | 1.48 | 0.5325 | 0.09 |
| Phospholipids to total lipids ratio in large LDL | 21.45 | 0.84 | 21.46 | 0.98 | 9.50E-01 | -0.01 | 20.75 | 0.49 | 20.86 | 0.49 | 7.99E-02 | -0.23 | 25.40 | 3.25 | 26.15 | 5.75 | 2.38E-01 | -0.17 | 2.02E-19 | -0.93 | 0.8653 | -0.02 |
| Cholesterol to total lipids ratio in large LDL | 71.41 | 0.99 | 71.19 | 1.60 | 2.32E-01 | 0.17 | 69.20 | 2.43 | 68.92 | 2.00 | 3.25E-01 | 0.13 | 63.96 | 5.32 | 62.64 | 9.72 | 2.14E-01 | 0.18 | 3.47E-35 | -1.24 | 0.8630 | 0.02 |
| CEs to total lipids ratio in large LDL | 52.59 | 1.15 | 52.51 | 1.74 | 6.80E-01 | 0.06 | 52.04 | 1.47 | 51.77 | 1.30 | 1.33E-01 | 0.20 | 45.44 | 5.25 | 44.39 | 7.75 | 2.41E-01 | 0.16 | 1.48E-07 | -0.44 | 0.7015 | 0.05 |
| Free cholesterol to total lipids ratio in large LDL | 18.82 | 0.83 | 18.69 | 0.88 | 2.60E-01 | 0.15 | 17.16 | 1.61 | 17.15 | 1.20 | 9.45E-01 | 0.01 | 18.52 | 1.82 | 18.26 | 2.66 | 3.87E-01 | 0.12 | 2.23E-49 | -1.40 | 0.7183 | -0.05 |
| TGs to total lipids ratio in large LDL | 7.14 | 1.12 | 7.34 | 1.37 | 2.25E-01 | -0.17 | 10.04 | 2.52 | 10.22 | 2.17 | 5.70E-01 | -0.07 | 10.64 | 3.16 | 11.21 | 4.53 | 2.80E-01 | -0.15 | 6.38E-51 | 1.59 | 0.8858 | 0.02 |
| Phospholipids to total lipids ratio in medium LDL | 26.74 | 0.78 | 26.84 | 1.17 | 4.61E-01 | -0.10 | 25.45 | 0.73 | 25.53 | 0.65 | 4.03E-01 | -0.11 | 29.05 | 2.79 | 29.58 | 3.56 | 2.28E-01 | -0.16 | 3.10E-47 | -1.53 | 0.9036 | 0.02 |
| Cholesterol to total lipids ratio in medium LDL | 66.57 | 1.49 | 66.27 | 2.16 | 2.30E-01 | 0.16 | 65.52 | 1.85 | 65.30 | 1.91 | 3.83E-01 | 0.11 | 61.89 | 4.35 | 60.78 | 7.17 | 1.69E-01 | 0.19 | 4.86E-12 | -0.53 | 0.9404 | 0.01 |
| CEs to total lipids ratio in medium LDL | 46.38 | 2.17 | 46.13 | 2.98 | 4.68E-01 | 0.10 | 48.57 | 2.09 | 48.38 | 1.75 | 4.53E-01 | 0.10 | 40.58 | 5.85 | 39.52 | 5.89 | 1.86E-01 | 0.18 | 1.95E-32 | 0.98 | 0.8524 | 0.03 |
| Free cholesterol to total lipids ratio in medium LDL | 20.19 | 1.42 | 20.15 | 1.65 | 8.29E-01 | 0.03 | 16.95 | 2.23 | 16.92 | 1.83 | 9.24E-01 | 0.01 | 21.31 | 2.92 | 21.26 | 3.41 | 9.04E-01 | 0.02 | 2.12E-65 | -1.80 | 0.8814 | -0.02 |
| TGs to total lipids ratio in medium LDL | 6.69 | 1.34 | 6.89 | 1.68 | 3.26E-01 | -0.13 | 9.03 | 1.96 | 9.17 | 1.96 | 5.88E-01 | -0.07 | 9.06 | 2.40 | 9.64 | 3.96 | 1.89E-01 | -0.18 | 3.32E-46 | 1.32 | 0.8568 | 0.02 |
| Phospholipids to total lipids ratio in small LDL | 31.56 | 1.44 | 31.87 | 2.05 | 1.98E-01 | -0.18 | 30.39 | 1.33 | 30.57 | 1.17 | 2.69E-01 | -0.14 | 38.95 | 2.77 | 39.76 | 5.75 | 1.83E-01 | -0.19 | 1.18E-22 | -0.82 | 0.8296 | 0.03 |
| Cholesterol to total lipids ratio in small LDL | 62.39 | 1.47 | 61.98 | 2.16 | 9.97E-02 | 0.23 | 60.83 | 2.09 | 60.62 | 2.22 | 4.68E-01 | 0.09 | 55.61 | 2.59 | 54.78 | 5.87 | 1.74E-01 | 0.20 | 2.10E-17 | -0.71 | 0.9520 | -0.01 |
| CEs to total lipids ratio in small LDL | 44.52 | 1.71 | 44.15 | 2.29 | 1.81E-01 | 0.18 | 45.93 | 1.57 | 45.75 | 1.56 | 3.75E-01 | 0.12 | 36.90 | 2.99 | 36.12 | 4.48 | 1.35E-01 | 0.21 | 4.13E-22 | 0.84 | 0.9998 | 0.00 |
| Free cholesterol to total lipids ratio in small LDL | 17.87 | 1.41 | 17.82 | 1.52 | 8.19E-01 | 0.03 | 14.89 | 2.15 | 14.87 | 1.83 | 9.30E-01 | 0.01 | 18.72 | 1.30 | 18.65 | 2.17 | 7.99E-01 | 0.04 | 3.02E-59 | -1.71 | 0.8807 | -0.02 |
| TGs to total lipids ratio in small LDL | 6.05 | 1.36 | 6.15 | 1.55 | 6.06E-01 | -0.07 | 8.79 | 2.17 | 8.81 | 1.93 | 9.28E-01 | -0.01 | 5.44 | 1.34 | 5.46 | 1.30 | 9.08E-01 | -0.02 | 5.07E-53 | 1.53 | 0.6725 | 0.06 |
| Phospholipids to total lipids ratio in very large HDL | 49.55 | 3.11 | 49.38 | 5.24 | 7.79E-01 | 0.04 | 49.36 | 2.66 | 48.99 | 5.12 | 4.80E-01 | 0.10 | 49.62 | 2.72 | 49.89 | 1.84 | 3.87E-01 | -0.12 | 1.23E-01 | -0.07 | 0.6525 | 0.06 |
| Cholesterol to total lipids ratio in very large HDL | 46.40 | 2.44 | 46.48 | 4.10 | 8.56E-01 | -0.03 | 43.49 | 1.72 | 43.08 | 4.11 | 3.20E-01 | 0.14 | 47.88 | 1.22 | 47.86 | 1.25 | 9.21E-01 | 0.01 | 2.01E-13 | -0.96 | 0.5838 | 0.08 |
| CEs to total lipids ratio in very large HDL | 34.84 | 1.71 | 34.66 | 2.20 | 4.89E-01 | 0.09 | 32.43 | 1.85 | 32.07 | 3.21 | 2.95E-01 | 0.14 | 31.05 | 1.81 | 31.22 | 1.23 | 4.23E-01 | -0.11 | 9.14E-19 | -1.08 | 0.7166 | 0.05 |
| Free cholesterol to total lipids ratio in very large HDL | 11.55 | 1.32 | 11.82 | 2.44 | 3.10E-01 | -0.14 | 11.06 | 0.96 | 11.01 | 1.66 | 7.73E-01 | 0.04 | 16.83 | 2.50 | 16.65 | 1.82 | 5.32E-01 | 0.09 | 1.77E-04 | -0.39 | 0.4279 | 0.11 |
| TGs to total lipids ratio in very large HDL | 4.06 | 1.61 | 4.14 | 2.25 | 7.62E-01 | -0.04 | 7.15 | 2.77 | 7.93 | 8.57 | 3.45E-01 | -0.14 | 2.50 | 1.99 | 2.25 | 1.09 | 2.41E-01 | 0.17 | 8.78E-16 | 0.82 | 0.3327 | -0.15 |
| Phospholipids to total lipids ratio in large HDL | 49.38 | 1.77 | 49.43 | 2.05 | 8.61E-01 | -0.02 | 47.76 | 1.80 | 47.39 | 2.54 | 1.93E-01 | 0.17 | 50.40 | 3.13 | 50.67 | 2.86 | 5.11E-01 | -0.09 | 6.32E-31 | -0.89 | 0.5267 | 0.09 |
| Cholesterol to total lipids ratio in large HDL | 46.02 | 2.78 | 45.96 | 3.14 | 8.88E-01 | 0.02 | 44.15 | 3.45 | 44.17 | 3.91 | 9.57E-01 | -0.01 | 44.71 | 7.68 | 45.55 | 4.59 | 3.32E-01 | -0.14 | 1.90E-16 | -0.55 | 0.9312 | 0.01 |
| CEs to total lipids ratio in large HDL | 35.50 | 2.48 | 35.46 | 2.76 | 9.04E-01 | 0.02 | 33.03 | 3.37 | 33.04 | 3.45 | 9.94E-01 | 0.00 | 35.02 | 5.87 | 35.75 | 3.17 | 2.51E-01 | -0.16 | 1.70E-27 | -0.81 | 0.8922 | 0.02 |
| Free cholesterol to total lipids ratio in large HDL | 10.51 | 0.49 | 10.50 | 0.51 | 8.38E-01 | 0.03 | 11.11 | 0.53 | 11.13 | 0.62 | 7.62E-01 | -0.04 | 9.69 | 1.89 | 9.79 | 1.53 | 6.73E-01 | -0.06 | 1.11E-48 | 1.15 | 0.8952 | -0.02 |
| TGs to total lipids ratio in large HDL | 4.60 | 1.43 | 4.62 | 1.50 | 9.55E-01 | -0.01 | 8.09 | 3.00 | 8.44 | 2.45 | 3.30E-01 | -0.13 | 4.89 | 7.70 | 3.79 | 2.17 | 1.53E-01 | 0.22 | 1.78E-60 | 1.74 | 0.2859 | -0.15 |
| Phospholipids to total lipids ratio in medium HDL | 45.89 | 0.82 | 46.03 | 0.89 | 2.59E-01 | -0.15 | 46.39 | 1.40 | 46.18 | 1.00 | 1.83E-01 | 0.18 | 48.98 | 1.46 | 48.88 | 1.35 | 6.05E-01 | 0.07 | 5.77E-05 | 0.30 | 0.5103 | 0.09 |
| Cholesterol to total lipids ratio in medium HDL | 49.22 | 1.95 | 49.05 | 1.90 | 5.22E-01 | 0.09 | 44.51 | 4.23 | 44.19 | 3.24 | 5.21E-01 | 0.08 | 46.31 | 2.93 | 46.60 | 2.45 | 4.36E-01 | -0.11 | 3.98E-56 | -1.68 | 0.8278 | 0.03 |
| CEs to total lipids ratio in medium HDL | 40.04 | 1.84 | 39.96 | 1.82 | 7.68E-01 | 0.04 | 34.79 | 4.09 | 34.42 | 3.10 | 4.42E-01 | 0.10 | 40.27 | 2.19 | 40.55 | 1.62 | 2.70E-01 | -0.15 | 2.43E-64 | -1.98 | 0.6842 | 0.06 |
| Free cholesterol to total lipids ratio in medium HDL | 9.18 | 0.40 | 9.09 | 0.54 | 1.39E-01 | 0.20 | 9.72 | 0.51 | 9.77 | 0.60 | 5.03E-01 | -0.09 | 6.05 | 1.18 | 6.04 | 1.34 | 9.88E-01 | 0.00 | 8.97E-41 | 1.19 | 0.4159 | -0.11 |
| TGs to total lipids ratio in medium HDL | 4.89 | 1.29 | 4.92 | 1.30 | 8.34E-01 | -0.03 | 9.10 | 3.07 | 9.63 | 2.60 | 1.56E-01 | -0.19 | 4.71 | 1.72 | 4.52 | 1.46 | 3.86E-01 | 0.12 | 3.24E-67 | 2.16 | 0.2438 | -0.16 |
| Phospholipids to total lipids ratio in small HDL | 57.87 | 1.18 | 57.88 | 1.08 | 9.69E-01 | -0.01 | 57.97 | 1.47 | 57.63 | 1.60 | 9.28E-02 | 0.22 | 57.73 | 0.78 | 57.78 | 0.87 | 6.56E-01 | -0.06 | 3.15E-01 | -0.06 | 0.6079 | 0.07 |
| Cholesterol to total lipids ratio in small HDL | 37.91 | 1.66 | 37.83 | 1.69 | 7.07E-01 | 0.05 | 34.13 | 2.41 | 33.98 | 2.25 | 6.37E-01 | 0.06 | 37.71 | 1.00 | 37.75 | 0.97 | 7.71E-01 | -0.04 | 2.26E-63 | -1.91 | 0.8604 | 0.02 |
| CEs to total lipids ratio in small HDL | 27.20 | 1.91 | 27.17 | 1.96 | 9.04E-01 | 0.02 | 21.91 | 2.98 | 21.45 | 2.96 | 2.36E-01 | 0.15 | 28.53 | 1.11 | 28.59 | 1.13 | 7.11E-01 | -0.05 | 5.21E-72 | -2.24 | 0.4288 | 0.11 |
| Free cholesterol to total lipids ratio in small HDL | 10.71 | 0.62 | 10.66 | 0.65 | 5.36E-01 | 0.08 | 12.22 | 1.04 | 12.54 | 1.13 | 2.65E-02 | -0.29 | 9.18 | 0.50 | 9.16 | 0.57 | 8.08E-01 | 0.03 | 6.80E-65 | 1.96 | 0.0850 | -0.23 |
| TGs to total lipids ratio in small HDL | 4.22 | 0.98 | 4.30 | 1.03 | 5.62E-01 | -0.08 | 7.90 | 2.28 | 8.39 | 2.06 | 9.04E-02 | -0.22 | 4.55 | 1.12 | 4.46 | 1.02 | 5.41E-01 | 0.08 | 1.90E-77 | 2.44 | 0.2021 | -0.17 |
