## Supplementary material for "Levothyroxine treatment during pregnancy: a metabolomics study": Table S2

Supplementary table 2: Correlations between metabolomics variables measured from CORD serum samples and characteristics of the born child

| Variable | | | TSH | | | | Apgar score, 1 min | | | | Apgar score, 5 min | | | | Child weight | | | | Head circumstance | | | | Placenta weight | | | |
|---|---|---|---|---|---|---|---|---|---|---|---|---|---|---|---|---|---|---|---|---|---|---|---|---|---|---|
| | | | Controls | | Levothyroxine | | Controls | | Levothyroxine | | Controls | | Levothyroxine | | Controls | | Levothyroxine | | Controls | | Levothyroxine | | Controls | | Levothyroxine | |
| Group | Subgroup | Variable | β | p | β | p | β | p | β | p | β | p | β | p | β | p | β | p | β | p | β | p | β | p | β | p |
| Cholesterol | | Total cholesterol | 0.02 | 8.34E-01 | -0.03 | 7.52E-01 | -0.24 | 1.16E-02 | -0.31 | 1.14E-03 | -0.11 | 2.61E-01 | -0.07 | 5.08E-01 | -0.17 | 8.48E-02 | -0.12 | 2.13E-01 | -0.13 | 1.77E-01 | -0.01 | 9.18E-01 | -0.04 | 7.00E-01 | 0.16 | 9.41E-02 |
| | | Total cholesterol minus HDL-C | 0.01 | 9.38E-01 | 0.03 | 7.46E-01 | -0.24 | 1.08E-02 | -0.34 | 4.45E-04 | -0.17 | 8.15E-02 | -0.15 | 1.20E-01 | -0.19 | 4.28E-02 | -0.16 | 9.84E-02 | -0.18 | 6.71E-02 | 0.00 | 9.67E-01 | -0.09 | 3.40E-01 | 0.11 | 2.50E-01 |
| | | Remnant cholesterol | 0.02 | 8.57E-01 | 0.08 | 4.47E-01 | -0.21 | 3.23E-02 | -0.39 | 4.27E-05 | -0.16 | 1.01E-01 | -0.23 | 1.82E-02 | -0.26 | 5.74E-03 | -0.22 | 2.32E-02 | -0.24 | 1.65E-02 | 0.03 | 7.82E-01 | -0.13 | 1.92E-01 | 0.05 | 5.82E-01 |
| | | VLDL cholesterol | 0.02 | 8.73E-01 | 0.18 | 7.30E-02 | -0.19 | 5.18E-02 | -0.30 | 2.09E-03 | -0.18 | 6.09E-02 | -0.33 | 6.36E-04 | -0.29 | 2.47E-03 | -0.19 | 4.58E-02 | -0.25 | 1.12E-02 | 0.11 | 2.67E-01 | -0.22 | 2.10E-02 | -0.03 | 7.65E-01 |
| | | Clinical LDL cholesterol | -0.01 | 9.49E-01 | -0.02 | 8.26E-01 | -0.24 | 1.26E-02 | -0.30 | 1.75E-03 | -0.16 | 1.00E-01 | -0.07 | 4.94E-01 | -0.13 | 1.68E-01 | -0.13 | 1.92E-01 | -0.14 | 1.66E-01 | -0.04 | 6.73E-01 | -0.04 | 6.85E-01 | 0.14 | 1.47E-01 |
| | | LDL cholesterol | 0.00 | 9.71E-01 | -0.01 | 9.47E-01 | -0.26 | 5.36E-03 | -0.28 | 3.78E-03 | -0.17 | 8.11E-02 | -0.08 | 4.10E-01 | -0.12 | 1.96E-01 | -0.11 | 2.82E-01 | -0.12 | 2.13E-01 | -0.01 | 8.87E-01 | -0.06 | 5.50E-01 | 0.16 | 1.09E-01 |
| | | HDL cholesterol | 0.04 | 6.88E-01 | -0.13 | 1.86E-01 | -0.17 | 7.56E-02 | -0.18 | 7.22E-02 | 0.03 | 7.34E-01 | 0.11 | 2.81E-01 | -0.06 | 5.10E-01 | -0.02 | 8.76E-01 | -0.01 | 9.51E-01 | -0.03 | 7.36E-01 | 0.08 | 4.31E-01 | 0.20 | 4.04E-02 |
| Triglycerides | | Total TGs | 0.04 | 7.14E-01 | 0.19 | 5.61E-02 | -0.15 | 1.08E-01 | -0.09 | 3.46E-01 | -0.13 | 1.75E-01 | -0.24 | 1.27E-02 | -0.22 | 2.22E-02 | -0.04 | 6.77E-01 | -0.16 | 1.16E-01 | 0.18 | 7.03E-02 | -0.26 | 5.66E-03 | 0.00 | 9.61E-01 |
| | | TGs in VLDL | 0.06 | 5.13E-01 | 0.22 | 2.89E-02 | -0.12 | 2.31E-01 | -0.08 | 3.88E-01 | -0.10 | 2.88E-01 | -0.23 | 2.03E-02 | -0.21 | 2.63E-02 | -0.07 | 4.92E-01 | -0.12 | 2.39E-01 | 0.15 | 1.28E-01 | -0.27 | 4.74E-03 | -0.05 | 6.33E-01 |
| | | TGs in LDL | -0.06 | 5.43E-01 | 0.07 | 4.95E-01 | -0.20 | 3.54E-02 | -0.09 | 3.36E-01 | -0.15 | 1.26E-01 | -0.22 | 2.21E-02 | -0.15 | 1.14E-01 | 0.04 | 6.88E-01 | -0.18 | 6.73E-02 | 0.16 | 1.07E-01 | -0.16 | 9.43E-02 | 0.10 | 2.91E-01 |
| | | TGs in HDL | 0.02 | 8.51E-01 | 0.12 | 2.45E-01 | -0.24 | 1.35E-02 | -0.09 | 3.84E-01 | -0.19 | 5.40E-02 | -0.20 | 3.95E-02 | -0.22 | 2.08E-02 | 0.00 | 9.95E-01 | -0.22 | 2.48E-02 | 0.24 | 1.62E-02 | -0.23 | 1.60E-02 | 0.07 | 4.52E-01 |
| Phospholipids | | Phospholipids in lipoprotein particles | 0.03 | 7.57E-01 | -0.03 | 7.94E-01 | -0.26 | 6.94E-03 | -0.28 | 3.80E-03 | -0.11 | 2.57E-01 | -0.06 | 5.23E-01 | -0.18 | 6.42E-02 | -0.08 | 4.14E-01 | -0.13 | 1.90E-01 | 0.05 | 6.49E-01 | -0.05 | 5.88E-01 | 0.18 | 6.35E-02 |
| | | Phospholipids in VLDL | 0.02 | 8.67E-01 | 0.19 | 5.29E-02 | -0.17 | 7.71E-02 | -0.21 | 3.13E-02 | -0.17 | 7.34E-02 | -0.31 | 1.22E-03 | -0.26 | 6.56E-03 | -0.14 | 1.62E-01 | -0.22 | 2.87E-02 | 0.14 | 1.54E-01 | -0.24 | 1.14E-02 | -0.03 | 7.24E-01 |
| | | Phospholipids in LDL | -0.01 | 8.87E-01 | -0.01 | 9.36E-01 | -0.26 | 6.45E-03 | -0.29 | 2.59E-03 | -0.18 | 6.87E-02 | -0.11 | 2.80E-01 | -0.15 | 1.25E-01 | -0.11 | 2.68E-01 | -0.15 | 1.36E-01 | -0.01 | 8.85E-01 | -0.07 | 4.50E-01 | 0.15 | 1.34E-01 |
| | | Phospholipids in HDL | 0.05 | 6.19E-01 | -0.12 | 2.44E-01 | -0.20 | 3.70E-02 | -0.16 | 9.29E-02 | -0.01 | 8.91E-01 | 0.10 | 3.26E-01 | -0.09 | 3.63E-01 | 0.01 | 9.44E-01 | -0.03 | 7.66E-01 | 0.02 | 8.33E-01 | 0.04 | 6.73E-01 | 0.21 | 2.96E-02 |
| Cholesteryl esters | | Total esterified cholesterol | 0.02 | 8.23E-01 | -0.04 | 6.61E-01 | -0.24 | 1.12E-02 | -0.30 | 1.62E-03 | -0.10 | 2.89E-01 | -0.05 | 6.39E-01 | -0.15 | 1.18E-01 | -0.11 | 2.58E-01 | -0.12 | 2.34E-01 | -0.01 | 8.87E-01 | -0.03 | 7.89E-01 | 0.17 | 8.02E-02 |
| | | CEs in VLDL | 0.01 | 9.22E-01 | 0.17 | 9.96E-02 | -0.19 | 4.85E-02 | -0.33 | 5.77E-04 | -0.18 | 5.60E-02 | -0.33 | 4.55E-04 | -0.29 | 2.52E-03 | -0.22 | 2.64E-02 | -0.26 | 8.11E-03 | 0.10 | 3.38E-01 | -0.20 | 3.42E-02 | -0.03 | 7.66E-01 |
| | | CEs in LDL | 0.00 | 9.78E-01 | 0.01 | 9.02E-01 | -0.28 | 3.78E-03 | -0.27 | 4.59E-03 | -0.18 | 6.09E-02 | -0.10 | 3.04E-01 | -0.13 | 1.72E-01 | -0.10 | 3.05E-01 | -0.13 | 1.96E-01 | 0.00 | 9.73E-01 | -0.07 | 4.61E-01 | 0.15 | 1.21E-01 |
| | | CEs in HDL | 0.04 | 6.65E-01 | -0.14 | 1.73E-01 | -0.17 | 6.98E-02 | -0.17 | 8.85E-02 | 0.04 | 7.10E-01 | 0.12 | 2.32E-01 | -0.05 | 5.79E-01 | -0.01 | 9.46E-01 | 0.01 | 9.36E-01 | -0.04 | 7.13E-01 | 0.08 | 4.03E-01 | 0.20 | 3.73E-02 |
| Free cholesterol | | Total free cholesterol | 0.02 | 8.75E-01 | 0.00 | 9.65E-01 | -0.24 | 1.38E-02 | -0.34 | 4.00E-04 | -0.12 | 2.11E-01 | -0.12 | 2.13E-01 | -0.20 | 3.95E-02 | -0.15 | 1.25E-01 | -0.17 | 8.71E-02 | 0.00 | 9.89E-01 | -0.06 | 5.10E-01 | 0.14 | 1.52E-01 |
| | | Free cholesterol in VLDL | 0.03 | 7.99E-01 | 0.19 | 5.33E-02 | -0.18 | 6.42E-02 | -0.25 | 1.09E-02 | -0.17 | 7.65E-02 | -0.31 | 1.30E-03 | -0.28 | 3.23E-03 | -0.16 | 9.24E-02 | -0.23 | 2.04E-02 | 0.13 | 2.03E-01 | -0.24 | 1.22E-02 | -0.03 | 7.59E-01 |
| | | Free cholesterol in LDL | -0.01 | 9.49E-01 | -0.05 | 5.88E-01 | -0.23 | 1.56E-02 | -0.28 | 3.42E-03 | -0.13 | 1.66E-01 | -0.03 | 7.98E-01 | -0.10 | 2.87E-01 | -0.12 | 2.38E-01 | -0.11 | 2.64E-01 | -0.06 | 5.38E-01 | -0.02 | 8.39E-01 | 0.16 | 9.88E-02 |
| | | Free cholesterol in HDL | 0.03 | 7.58E-01 | -0.12 | 2.48E-01 | -0.16 | 1.01E-01 | -0.20 | 4.23E-02 | 0.02 | 8.17E-01 | 0.07 | 4.89E-01 | -0.10 | 3.23E-01 | -0.04 | 6.84E-01 | -0.05 | 6.03E-01 | -0.02 | 8.18E-01 | 0.06 | 5.34E-01 | 0.19 | 4.91E-02 |
| Total lipids | | Lipids in lipoprotein particles | 0.03 | 7.47E-01 | 0.03 | 8.03E-01 | -0.27 | 5.19E-03 | -0.30 | 1.69E-03 | -0.13 | 1.74E-01 | -0.12 | 2.14E-01 | -0.21 | 2.82E-02 | -0.11 | 2.61E-01 | -0.16 | 1.11E-01 | 0.05 | 6.11E-01 | -0.10 | 2.90E-01 | 0.15 | 1.15E-01 |
| | | Total lipids in VLDL | 0.04 | 6.77E-01 | 0.21 | 3.63E-02 | -0.16 | 1.01E-01 | -0.19 | 5.70E-02 | -0.15 | 1.21E-01 | -0.29 | 2.84E-03 | -0.26 | 6.14E-03 | -0.13 | 1.93E-01 | -0.19 | 5.17E-02 | 0.15 | 1.52E-01 | -0.26 | 5.63E-03 | -0.04 | 6.82E-01 |
| | | Total lipids in LDL | -0.01 | 9.07E-01 | 0.00 | 9.88E-01 | -0.27 | 5.04E-03 | -0.28 | 4.12E-03 | -0.17 | 7.08E-02 | -0.11 | 2.84E-01 | -0.14 | 1.53E-01 | -0.10 | 3.32E-01 | -0.14 | 1.59E-01 | 0.00 | 9.74E-01 | -0.07 | 4.49E-01 | 0.16 | 1.07E-01 |
| | | Total lipids in HDL | 0.05 | 6.45E-01 | -0.12 | 2.44E-01 | -0.20 | 4.11E-02 | -0.18 | 7.05E-02 | 0.00 | 9.98E-01 | 0.09 | 3.72E-01 | -0.09 | 3.58E-01 | 0.00 | 9.64E-01 | -0.03 | 7.62E-01 | 0.01 | 9.29E-01 | 0.05 | 6.40E-01 | 0.21 | 2.96E-02 |
| Lipoprotein particle concentrations | | Conc. of lipoprotein particles | 0.04 | 6.63E-01 | -0.10 | 3.44E-01 | -0.31 | 9.43E-04 | -0.15 | 1.30E-01 | -0.10 | 2.82E-01 | 0.08 | 4.40E-01 | -0.08 | 4.08E-01 | 0.05 | 6.42E-01 | -0.02 | 8.37E-01 | 0.07 | 5.14E-01 | -0.02 | 8.14E-01 | 0.25 | 1.14E-02 |
| | | Conc. of VLDL particles | -0.01 | 9.49E-01 | 0.17 | 9.89E-02 | -0.19 | 4.64E-02 | -0.21 | 3.16E-02 | -0.19 | 4.98E-02 | -0.30 | 1.65E-03 | -0.25 | 8.77E-03 | -0.12 | 2.33E-01 | -0.23 | 1.82E-02 | 0.15 | 1.30E-01 | -0.22 | 1.95E-02 | 0.00 | 9.90E-01 |
| | | Conc. of LDL particles | -0.01 | 9.05E-01 | 0.08 | 4.25E-01 | -0.23 | 1.64E-02 | -0.34 | 3.25E-04 | -0.18 | 5.94E-02 | -0.26 | 7.56E-03 | -0.21 | 2.63E-02 | -0.19 | 5.41E-02 | -0.21 | 3.61E-02 | 0.02 | 8.66E-01 | -0.13 | 1.83E-01 | 0.07 | 5.08E-01 |
| | | Conc. of HDL particles | 0.05 | 6.34E-01 | -0.11 | 2.72E-01 | -0.31 | 1.19E-03 | -0.12 | 2.21E-01 | -0.09 | 3.65E-01 | 0.11 | 2.66E-01 | -0.06 | 5.34E-01 | 0.07 | 4.74E-01 | 0.00 | 9.72E-01 | 0.07 | 5.10E-01 | -0.01 | 9.26E-01 | 0.25 | 8.81E-03 |
| Lipoprotein particle sizes | | Average diameter for VLDL particles | 0.13 | 1.88E-01 | 0.23 | 1.90E-02 | -0.14 | 1.46E-01 | -0.12 | 2.13E-01 | -0.13 | 1.94E-01 | -0.18 | 6.14E-02 | -0.22 | 1.98E-02 | -0.07 | 4.54E-01 | -0.14 | 1.71E-01 | 0.10 | 3.41E-01 | -0.30 | 1.51E-03 | -0.07 | 5.07E-01 |
| | | Average diameter for LDL particles | -0.08 | 4.47E-01 | -0.16 | 1.09E-01 | -0.04 | 6.61E-01 | -0.18 | 7.16E-02 | -0.08 | 4.22E-01 | -0.23 | 1.76E-02 | 0.06 | 5.06E-01 | -0.11 | 2.49E-01 | 0.06 | 5.59E-01 | -0.24 | 1.69E-02 | 0.08 | 4.07E-01 | 0.05 | 6.40E-01 |
| | | Average diameter for HDL particles | 0.04 | 6.51E-01 | -0.10 | 3.09E-01 | -0.04 | 7.07E-01 | -0.19 | 5.16E-02 | 0.09 | 3.49E-01 | 0.02 | 8.18E-01 | -0.08 | 4.19E-01 | -0.13 | 1.99E-01 | -0.07 | 5.00E-01 | -0.09 | 3.68E-01 | 0.13 | 1.92E-01 | 0.07 | 4.90E-01 |
| Other lipids | | Phosphoglycerides | 0.00 | 9.73E-01 | -0.04 | 6.91E-01 | -0.29 | 2.55E-03 | -0.23 | 1.99E-02 | -0.11 | 2.48E-01 | -0.05 | 5.90E-01 | -0.16 | 1.06E-01 | 0.00 | 1.00E+00 | -0.11 | 2.73E-01 | 0.09 | 3.98E-01 | -0.08 | 3.98E-01 | 0.22 | 2.30E-02 |
| | | Ratio of TGs to phosphoglycerides | 0.04 | 7.12E-01 | 0.23 | 1.96E-02 | 0.01 | 8.86E-01 | 0.00 | 9.91E-01 | -0.05 | 5.96E-01 | -0.21 | 2.91E-02 | -0.15 | 1.08E-01 | -0.02 | 8.07E-01 | -0.10 | 3.25E-01 | 0.15 | 1.35E-01 | -0.23 | 1.43E-02 | -0.10 | 3.30E-01 |
| | | Total cholines | 0.00 | 9.97E-01 | -0.05 | 6.15E-01 | -0.29 | 2.58E-03 | -0.24 | 1.20E-02 | -0.10 | 2.92E-01 | -0.05 | 6.37E-01 | -0.15 | 1.25E-01 | 0.00 | 9.93E-01 | -0.10 | 2.96E-01 | 0.07 | 4.96E-01 | -0.06 | 5.12E-01 | 0.23 | 1.72E-02 |
| | | Phosphatidylcholines | 0.00 | 9.89E-01 | -0.06 | 5.39E-01 | -0.27 | 4.63E-03 | -0.20 | 3.87E-02 | -0.12 | 2.31E-01 | -0.05 | 6.45E-01 | -0.17 | 7.83E-02 | -0.03 | 7.93E-01 | -0.13 | 1.87E-01 | 0.08 | 4.09E-01 | -0.07 | 4.48E-01 | 0.20 | 4.15E-02 |
| | | Sphingomyelins | 0.02 | 8.56E-01 | -0.02 | 8.33E-01 | -0.27 | 5.31E-03 | -0.30 | 1.49E-03 | -0.08 | 3.93E-01 | -0.04 | 6.76E-01 | -0.11 | 2.47E-01 | -0.02 | 8.17E-01 | -0.08 | 4.30E-01 | 0.03 | 7.49E-01 | 0.01 | 9.55E-01 | 0.23 | 2.00E-02 |
| Apolipoproteins | | Apolipoprotein B | -0.01 | 8.95E-01 | 0.09 | 3.64E-01 | -0.22 | 2.09E-02 | -0.35 | 2.18E-04 | -0.19 | 5.25E-02 | -0.26 | 6.31E-03 | -0.23 | 1.75E-02 | -0.19 | 5.40E-02 | -0.22 | 2.41E-02 | 0.03 | 7.85E-01 | -0.13 | 1.68E-01 | 0.06 | 5.62E-01 |
| | | Apolipoprotein A1 | 0.05 | 6.20E-01 | -0.12 | 2.47E-01 | -0.25 | 9.42E-03 | -0.15 | 1.26E-01 | -0.04 | 6.97E-01 | 0.10 | 3.12E-01 | -0.09 | 3.74E-01 | 0.03 | 7.73E-01 | -0.02 | 8.10E-01 | 0.04 | 6.79E-01 | 0.02 | 8.59E-01 | 0.24 | 1.55E-02 |
| | | Ratio of apolipoprotein B to apolipoprotein A1 | -0.06 | 5.73E-01 | 0.19 | 6.23E-02 | -0.03 | 7.61E-01 | -0.23 | 2.00E-02 | -0.16 | 1.07E-01 | -0.34 | 3.31E-04 | -0.17 | 6.95E-02 | -0.19 | 5.47E-02 | -0.20 | 4.22E-02 | 0.01 | 9.60E-01 | -0.16 | 9.57E-02 | -0.11 | 2.48E-01 |
| Fatty acids | | Total FAs | 0.01 | 9.37E-01 | 0.05 | 6.23E-01 | -0.28 | 3.41E-03 | -0.28 | 4.23E-03 | -0.13 | 1.86E-01 | -0.21 | 3.19E-02 | -0.23 | 1.70E-02 | -0.07 | 4.82E-01 | -0.15 | 1.28E-01 | 0.09 | 3.80E-01 | -0.17 | 7.50E-02 | 0.14 | 1.67E-01 |
| | | Degree of unsaturation | -0.17 | 8.44E-02 | -0.19 | 6.53E-02 | -0.08 | 3.82E-01 | 0.12 | 2.29E-01 | -0.04 | 7.06E-01 | 0.24 | 1.46E-02 | 0.16 | 8.99E-02 | 0.13 | 1.71E-01 | 0.11 | 2.80E-01 | 0.09 | 3.71E-01 | 0.07 | 4.81E-01 | 0.21 | 3.49E-02 |
| | | Omega-3 FAs | -0.07 | 5.04E-01 | 0.02 | 8.57E-01 | -0.16 | 9.92E-02 | 0.00 | 9.64E-01 | -0.07 | 4.41E-01 | -0.02 | 8.66E-01 | -0.10 | 3.22E-01 | 0.05 | 6.29E-01 | -0.07 | 4.99E-01 | 0.19 | 5.86E-02 | -0.14 | 1.49E-01 | 0.12 | 2.22E-01 |
| | | Omega-6 FAs | -0.03 | 7.57E-01 | -0.10 | 3.09E-01 | -0.30 | 1.46E-03 | -0.27 | 4.63E-03 | -0.10 | 2.97E-01 | -0.09 | 3.38E-01 | -0.15 | 1.28E-01 | -0.07 | 4.89E-01 | -0.08 | 4.45E-01 | -0.03 | 7.42E-01 | -0.05 | 6.12E-01 | 0.17 | 7.55E-02 |
| | | PUFAs | -0.05 | 6.05E-01 | -0.07 | 4.92E-01 | -0.30 | 1.84E-03 | -0.21 | 3.32E-02 | -0.11 | 2.67E-01 | -0.08 | 4.19E-01 | -0.15 | 1.16E-01 | -0.03 | 7.27E-01 | -0.09 | 3.89E-01 | 0.05 | 6.22E-01 | -0.09 | 3.29E-01 | 0.18 | 6.59E-02 |
| | | MUFAs | 0.05 | 5.89E-01 | 0.13 | 1.83E-01 | -0.21 | 2.70E-02 | -0.27 | 4.47E-03 | -0.15 | 1.30E-01 | -0.36 | 1.21E-04 | -0.25 | 1.02E-02 | -0.09 | 3.44E-01 | -0.14 | 1.45E-01 | 0.10 | 3.28E-01 | -0.25 | 1.00E-02 | 0.01 | 8.92E-01 |
| | | Saturated FAs | 0.03 | 7.68E-01 | 0.07 | 4.70E-01 | -0.26 | 6.47E-03 | -0.27 | 4.41E-03 | -0.11 | 2.54E-01 | -0.15 | 1.21E-01 | -0.25 | 9.94E-03 | -0.07 | 4.95E-01 | -0.19 | 5.52E-02 | 0.10 | 3.19E-01 | -0.16 | 1.00E-01 | 0.16 | 9.95E-02 |
| | | LA | -0.04 | 7.23E-01 | -0.10 | 3.05E-01 | -0.28 | 3.50E-03 | -0.31 | 1.24E-03 | -0.08 | 4.24E-01 | -0.21 | 2.71E-02 | -0.17 | 7.13E-02 | -0.13 | 2.01E-01 | -0.10 | 3.40E-01 | -0.08 | 4.14E-01 | -0.06 | 5.05E-01 | 0.10 | 3.34E-01 |
| | | DHA | -0.06 | 5.71E-01 | -0.04 | 7.13E-01 | -0.19 | 4.62E-02 | -0.04 | 6.88E-01 | -0.08 | 3.85E-01 | -0.04 | 7.06E-01 | -0.08 | 4.09E-01 | 0.04 | 6.70E-01 | -0.04 | 6.70E-01 | 0.21 | 4.23E-02 | -0.10 | 3.03E-01 | 0.13 | 1.83E-01 |
| | Fatty acid ratios | Ratio of omega-3 FAs to total FAs | -0.11 | 2.72E-01 | 0.01 | 9.41E-01 | -0.05 | 5.77E-01 | 0.12 | 2.12E-01 | -0.04 | 6.93E-01 | 0.08 | 4.35E-01 | 0.00 | 9.89E-01 | 0.10 | 3.07E-01 | -0.02 | 8.54E-01 | 0.22 | 3.32E-02 | -0.12 | 2.23E-01 | 0.09 | 3.39E-01 |
| | | Ratio of omega-6 FAs to total FAs | -0.08 | 4.40E-01 | -0.20 | 5.11E-02 | 0.14 | 1.55E-01 | 0.14 | 1.51E-01 | 0.12 | 2.24E-01 | 0.18 | 6.72E-02 | 0.19 | 5.11E-02 | 0.00 | 9.88E-01 | 0.15 | 1.22E-01 | -0.17 | 8.59E-02 | 0.24 | 1.13E-02 | -0.05 | 5.98E-01 |
| | | Ratio of PUFA to total FAs | -0.17 | 9.18E-02 | -0.24 | 1.70E-02 | 0.14 | 1.34E-01 | 0.24 | 1.45E-02 | 0.13 | 1.80E-01 | 0.26 | 6.48E-03 | 0.24 | 1.10E-02 | 0.05 | 6.27E-01 | 0.19 | 5.78E-02 | -0.11 | 2.67E-01 | 0.24 | 1.17E-02 | -0.02 | 8.52E-01 |
| | | Ratio of MUFAs to total FAs | 0.11 | 2.48E-01 | 0.18 | 7.93E-02 | -0.13 | 1.86E-01 | -0.18 | 7.00E-02 | -0.15 | 1.22E-01 | -0.31 | 1.41E-03 | -0.18 | 5.51E-02 | -0.03 | 7.30E-01 | -0.09 | 3.54E-01 | 0.07 | 4.94E-01 | -0.25 | 9.84E-03 | -0.05 | 6.47E-01 |
| | | Ratio of saturated FAs to total FAs | 0.11 | 2.63E-01 | 0.12 | 2.30E-01 | -0.05 | 5.95E-01 | -0.11 | 2.54E-01 | 0.02 | 8.32E-01 | 0.12 | 2.29E-01 | -0.13 | 1.88E-01 | -0.03 | 7.62E-01 | -0.19 | 4.89E-02 | 0.08 | 4.11E-01 | -0.01 | 8.98E-01 | 0.14 | 1.59E-01 |
| | | Ratio of LA to total FAs | -0.06 | 5.69E-01 | -0.21 | 3.51E-02 | -0.10 | 3.02E-01 | -0.05 | 5.93E-01 | 0.01 | 9.22E-01 | -0.02 | 8.45E-01 | 0.00 | 9.90E-01 | -0.03 | 3.53E-01 | 0.02 | 8.50E-01 | -0.21 | 3.94E-02 | -0.10 | 2.79E-01 | -0.03 | 7.67E-01 |
| | | Ratio of DHA to total FAs | -0.12 | 2.35E-01 | -0.09 | 3.58E-01 | -0.03 | 7.31E-01 | 0.15 | 1.28E-01 | -0.01 | 9.17E-01 | 0.11 | 2.74E-01 | 0.07 | 4.77E-01 | 0.13 | 2.00E-01 | 0.05 | 6.07E-01 | 0.25 | 1.21E-02 | -0.04 | 6.85E-01 | 0.09 | 3.80E-01 |
| | | Ratio of PUFAs to MUFAs | -0.16 | 1.04E-01 | -0.20 | 4.30E-02 | 0.19 | 5.16E-02 | 0.15 | 1.29E-01 | 0.18 | 5.57E-02 | 0.19 | 4.70E-02 | 0.16 | 1.06E-01 | -0.03 | 7.85E-01 | 0.08 | 4.05E-01 | -0.06 | 5.62E-01 | 0.21 | 3.14E-02 | 0.00 | 9.96E-01 |
| | | Ratio of omega-6 FAs to omega-3 FAs | 0.08 | 4.07E-01 | -0.11 | 2.83E-01 | 0.03 | 7.21E-01 | -0.01 | 9.28E-01 | 0.07 | 4.76E-01 | -0.02 | 8.69E-01 | 0.01 | 9.07E-01 | -0.10 | 3.23E-01 | 0.03 | 7.92E-01 | -0.22 | 2.65E-02 | 0.20 | 3.96E-02 | -0.12 | 2.25E-01 |
| Amino acids | | Alanine | 0.37 | 1.27E-04 | 0.34 | 5.06E-04 | -0.07 | 4.84E-01 | -0.48 | 2.00E-07 | 0.02 | 8.22E-01 | -0.54 | 1.90E-09 | -0.17 | 7.29E-02 | -0.07 | 5.01E-01 | -0.11 | 2.49E-01 | 0.13 | 2.10E-01 | -0.12 | 2.26E-01 | 0.02 | 8.71E-01 |
| | | Glutamine | 0.15 | 1.18E-01 | 0.11 | 2.98E-01 | 0.06 | 5.12E-01 | -0.24 | 1.52E-02 | -0.06 | 5.40E-01 | -0.18 | 5.88E-02 | -0.20 | 4.07E-02 | -0.03 | 7.55E-01 | 0.00 | 9.75E-01 | 0.09 | 3.55E-01 | -0.09 | 3.33E-01 | 0.17 | 7.94E-02 |
| | | Glycine | 0.18 | 7.55E-02 | 0.05 | 6.04E-01 | -0.07 | 4.60E-01 | -0.46 | 8.00E-07 | -0.09 | 3.63E-01 | -0.55 | 1.39E-09 | -0.15 | 1.13E-01 | -0.23 | 1.65E-02 | -0.03 | 7.93E-01 | 0.07 | 5.02E-01 | -0.13 | 1.76E-01 | -0.07 | 4.99E-01 |
| | | Histidine | 0.03 | 7.66E-01 | 0.07 | 4.87E-01 | 0.10 | 3.02E-01 | -0.39 | 4.53E-05 | 0.05 | 6.36E-01 | -0.23 | 1.75E-02 | -0.03 | 7.74E-01 | 0.03 | 7.96E-01 | 0.09 | 3.50E-01 | 0.11 | 2.72E-01 | -0.02 | 8.47E-01 | 0.14 | 1.60E-01 |
| | Branched-chain amino acids | Total Conc. of BCAA | 0.08 | 3.98E-01 | 0.06 | 5.63E-01 | -0.02 | 8.42E-01 | -0.45 | 1.30E-06 | 0.05 | 5.97E-01 | -0.45 | 1.20E-06 | 0.11 | 2.35E-01 | -0.23 | 1.94E-02 | 0.04 | 7.13E-01 | -0.03 | 7.91E-01 | 0.01 | 8.91E-01 | -0.04 | 6.72E-01 |
| | | Isoleucine | 0.04 | 6.60E-01 | 0.03 | 8.03E-01 | -0.01 | 9.19E-01 | -0.45 | 1.20E-06 | -0.02 | 8.59E-01 | -0.38 | 5.37E-05 | 0.15 | 1.15E-01 | -0.22 | 2.11E-02 | 0.07 | 4.98E-01 | -0.03 | 8.00E-01 | 0.04 | 6.90E-01 | 0.01 | 9.13E-01 |
| | | Leucine | 0.19 | 5.73E-02 | 0.07 | 4.83E-01 | -0.03 | 7.79E-01 | -0.50 | 3.38E-08 | 0.05 | 6.10E-01 | -0.50 | 4.04E-08 | 0.05 | 5.79E-01 | -0.29 | 3.04E-03 | 0.02 | 8.10E-01 | -0.05 | 5.93E-01 | -0.06 | 5.24E-01 | -0.07 | 4.56E-01 |
| | | Valine | 0.02 | 8.58E-01 | 0.05 | 6.07E-01 | -0.01 | 8.82E-01 | -0.35 | 2.69E-04 | 0.06 | 5.04E-01 | -0.38 | 6.62E-05 | 0.12 | 2.03E-01 | -0.15 | 1.16E-01 | 0.03 | 7.55E-01 | -0.01 | 9.42E-01 | 0.05 | 5.92E-01 | -0.03 | 7.27E-01 |
| | Aromatic amino acids | Phenylalanine | 0.26 | 6.56E-03 | 0.13 | 1.87E-01 | 0.01 | 9.05E-01 | -0.51 | 1.96E-08 | 0.11 | 2.39E-01 | -0.58 | 9.91E-11 | -0.19 | 4.91E-02 | -0.30 | 1.94E-03 | -0.11 | 2.81E-01 | -0.13 | 2.04E-01 | -0.18 | 5.65E-02 | -0.10 | 2.89E-01 |
| | | Tyrosine | 0.09 | 3.49E-01 | 0.14 | 1.76E-01 | 0.01 | 8.78E-01 | -0.45 | 1.50E-06 | 0.01 | 9.24E-01 | -0.46 | 7.00E-07 | -0.17 | 7.74E-02 | -0.28 | 3.08E-03 | -0.22 | 2.77E-02 | -0.01 | 9.44E-01 | -0.05 | 5.91E-01 | -0.09 | 3.42E-01 |
| Glycolysis related metabolites | | Glucose | 0.19 | 5.77E-02 | 0.21 | 3.73E-02 | -0.08 | 4.01E-01 | -0.06 | 5.58E-01 | -0.02 | 7.98E-01 | -0.01 | 8.92E-01 | 0.27 | 4.47E-03 | 0.14 | 1.57E-01 | 0.26 | 8.01E-03 | 0.00 | 9.70E-01 | 0.03 | 7.51E-01 | 0.04 | 7.18E-01 |
| | | Lactate | 0.50 | 5.24E-08 | 0.37 | 1.70E-04 | -0.09 | 3.65E-01 | -0.44 | 1.80E-06 | 0.00 | 9.98E-01 | -0.44 | 3.00E-06 | -0.20 | 3.82E-02 | -0.12 | 2.12E-01 | -0.11 | 2.68E-01 | -0.13 | 2.09E-01 | -0.14 | 1.37E-01 | -0.13 | 1.72E-01 |
| | | Pyruvate | 0.35 | 2.33E-04 | 0.34 | 4.66E-04 | -0.02 | 8.26E-01 | -0.13 | 1.68E-01 | 0.06 | 5.20E-01 | -0.13 | 1.83E-01 | -0.07 | 4.42E-01 | 0.07 | 4.95E-01 | -0.01 | 8.92E-01 | -0.02 | 8.28E-01 | -0.06 | 5.68E-01 | 0.01 | 8.85E-01 |
| | | Citrate | 0.18 | 6.83E-02 | 0.31 | 2.11E-03 | 0.05 | 6.37E-01 | -0.28 | 4.79E-03 | 0.02 | 8.59E-01 | -0.23 | 2.07E-02 | -0.20 | 3.63E-02 | -0.08 | 4.38E-01 | -0.10 | 3.04E-01 | 0.08 | 4.41E-01 | -0.08 | 4.16E-01 | 0.02 | 8.64E-01 |
| | | Glycerol | 0.22 | 3.04E-02 | 0.41 | 3.59E-05 | -0.21 | 2.92E-02 | -0.30 | 2.85E-03 | -0.13 | 1.90E-01 | -0.20 | 4.83E-02 | -0.27 | 5.32E-03 | 0.19 | 5.47E-02 | -0.11 | 2.68E-01 | 0.04 | 6.78E-01 | -0.27 | 5.10E-03 | 0.22 | 2.62E-02 |
| Ketone bodies | | 3-Hydroxybutyrate | -0.02 | 8.61E-01 | -0.05 | 6.07E-01 | -0.02 | 8.11E-01 | -0.16 | 1.05E-01 | -0.01 | 9.33E-01 | -0.03 | 7.70E-01 | 0.14 | 1.51E-01 | 0.02 | 8.68E-01 | 0.21 | 3.77E-02 | 0.00 | 9.77E-01 | 0.14 | 1.54E-01 | 0.19 | 5.26E-02 |
| | | Acetate | -0.10 | 3.36E-01 | -0.04 | 6.94E-01 | -0.02 | 8.10E-01 | -0.15 | 1.26E-01 | 0.04 | 6.63E-01 | -0.19 | 4.96E-02 | -0.17 | 8.30E-02 | -0.08 | 4.30E-01 | 0.00 | 9.89E-01 | -0.11 | 2.93E-01 | -0.06 | 5.03E-01 | 0.06 | 5.35E-01 |
| | | Acetoacetate | -0.12 | 2.27E-01 | -0.09 | 3.92E-01 | 0.00 | 9.65E-01 | -0.04 | 6.69E-01 | 0.02 | 8.33E-01 | 0.06 | 5.35E-01 | 0.17 | 7.11E-02 | 0.09 | 3.70E-01 | 0.22 | 2.81E-02 | 0.14 | 1.74E-01 | 0.12 | 2.13E-01 | 0.24 | 1.47E-02 |
| | | Acetone | -0.09 | 3.64E-01 | -0.11 | 2.58E-01 | -0.05 | 6.05E-01 | -0.03 | 7.37E-01 | -0.01 | 9.04E-01 | 0.05 | 5.90E-01 | 0.23 | 1.42E-02 | 0.08 | 4.05E-01 | 0.28 | 3.79E-03 | 0.04 | 7.18E-01 | 0.23 | 1.81E-02 | 0.20 | 4.24E-02 |
| Fluid balance | | Creatinine | 0.10 | 3.20E-01 | 0.04 | 7.24E-01 | 0.02 | 8.38E-01 | -0.04 | 6.74E-01 | -0.05 | 6.22E-01 | 0.02 | 8.71E-01 | 0.13 | 1.94E-01 | 0.14 | 1.67E-01 | 0.04 | 7.00E-01 | 0.09 | 3.78E-01 | -0.04 | 7.12E-01 | -0.01 | 9.10E-01 |
| | | Albumin | -0.06 | 5.27E-01 | -0.04 | 7.26E-01 | -0.30 | 1.46E-03 | 0.09 | 3.44E-01 | -0.08 | 4.00E-01 | 0.27 | 5.55E-03 | 0.13 | 1.86E-01 | 0.36 | 1.53E-04 | 0.19 | 5.61E-02 | 0.26 | 1.05E-02 | -0.05 | 5.85E-01 | 0.38 | 7.08E-05 |
| Inflammation | | Glycoprotein acetyls | 0.04 | 7.21E-01 | 0.09 | 3.58E-01 | -0.29 | 1.95E-03 | -0.34 | 4.30E-04 | -0.17 | 7.86E-02 | -0.33 | 4.57E-04 | 0.00 | 9.79E-01 | -0.04 | 6.61E-01 | 0.01 | 9.22E-01 | 0.14 | 1.68E-01 | 0.04 | 7.16E-01 | 0.12 | 2.36E-01 |
| Lipoprotein subclasses | CMs and extremely large VLDL (particle diameters > 75 nm) | Conc. of CMs and extremely large VLDL particles | 0.10 | 2.99E-01 | 0.23 | 1.89E-02 | -0.03 | 7.33E-01 | -0.19 | 5.10E-02 | -0.03 | 7.54E-01 | -0.32 | 7.45E-04 | -0.24 | 1.18E-02 | -0.18 | 7.26E-02 | -0.14 | 1.51E-01 | 0.10 | 3.16E-01 | -0.27 | 5.21E-03 | -0.11 | 2.85E-01 |
| | | Total lipids in CMs and extremely large VLDL | 0.13 | 2.04E-01 | 0.25 | 1.14E-02 | 0.01 | 9.06E-01 | -0.24 | 1.36E-02 | 0.03 | 7.39E-01 | -0.32 | 7.02E-04 | -0.26 | 6.77E-03 | -0.11 | 2.85E-02 | -0.13 | 2.00E-01 | 0.06 | 6.52E-01 | -0.24 | 1.14E-02 | -0.11 | 1.94E-01 |
| | | Phospholipids in CMs and extremely large VLDL | 0.10 | 2.97E-01 | 0.25 | 1.36E-02 | -0.06 | 5.30E-01 | -0.20 | 3.99E-02 | -0.03 | 7.54E-01 | -0.34 | 3.56E-04 | -0.23 | 1.68E-02 | -0.16 | 9.15E-02 | -0.13 | 1.97E-01 | 0.12 | 2.33E-01 | -0.25 | 1.01E-02 | -0.11 | 2.83E-01 |
| | | Cholesterol in CMs and extremely large VLDL | 0.08 | 4.26E-01 | 0.24 | 1.72E-02 | -0.08 | 4.13E-01 | -0.24 | 1.50E-02 | -0.03 | 7.60E-01 | -0.37 | 7.66E-05 | -0.27 | 4.53E-03 | -0.17 | 8.18E-02 | -0.16 | 1.03E-01 | 0.12 | 2.53E-01 | -0.28 | 2.87E-03 | -0.12 | 2.66E-01 |
| | | CEs in CMs and extremely large VLDL | 0.06 | 5.41E-01 | 0.23 | 2.21E-02 | -0.08 | 4.33E-01 | -0.25 | 8.70E-03 | -0.02 | 8.48E-01 | -0.40 | 1.78E-05 | -0.28 | 3.16E-03 | -0.17 | 7.33E-02 | -0.17 | 8.14E-02 | 0.10 | 3.49E-01 | -0.30 | 1.42E-03 | -0.12 | 2.20E-01 |
| | | Free cholesterol in CMs and extremely large VLDL | 0.10 | 3.01E-01 | 0.25 | 1.40E-02 | -0.08 | 3.93E-01 | -0.21 | 3.33E-02 | -0.05 | 6.38E-01 | -0.33 | 5.93E-04 | -0.25 | 8.75E-03 | -0.16 | 1.01E-01 | -0.14 | 1.51E-01 | 0.14 | 1.69E-01 | -0.25 | 8.46E-03 | -0.09 | 3.49E-01 |

| | | | TSH | | | | Apgar score, 1 min | | | | Apgar score, 5 min | | | | Child weight | | | | Head circumstance | | | | Placenta weight | | | |
|---|---|---|---|---|---|---|---|---|---|---|---|---|---|---|---|---|---|---|---|---|---|---|---|---|---|---|
| Variable | | | Controls | | Levothyroxine | | Controls | | Levothyroxine | | Controls | | Levothyroxine | | Controls | | Levothyroxine | | Controls | | Levothyroxine | | Controls | | Levothyroxine | |
| Group | Subgroup | Variable | β | p | β | p | β | p | β | p | β | p | β | p | β | p | β | p | β | p | β | p | β | p | β | p |
| | | TGs in CMs and extremely large VLDL | 0.14 | 1.44E-01 | 0.23 | 1.90E-02 | 0.08 | 3.94E-01 | -0.23 | 1.93E-02 | 0.08 | 3.92E-01 | -0.25 | 9.25E-03 | -0.23 | 1.44E-02 | -0.23 | 1.79E-02 | -0.09 | 3.43E-01 | -0.02 | 8.26E-01 | -0.19 | 4.50E-02 | -0.13 | 1.81E-01 |
| | Very large VLDL (average diameter 64 nm) | Conc. of very large VLDL particles | 0.04 | 6.90E-01 | 0.23 | 2.43E-02 | -0.09 | 3.69E-01 | -0.08 | 4.04E-01 | -0.10 | 2.84E-01 | -0.24 | 1.36E-02 | -0.21 | 3.12E-02 | -0.07 | 5.06E-01 | -0.13 | 1.93E-01 | 0.15 | 1.33E-01 | -0.26 | 6.55E-03 | -0.06 | 5.75E-01 |
| | | Total lipids in very large VLDL | 0.07 | 4.95E-01 | 0.24 | 1.69E-02 | -0.08 | 4.27E-01 | -0.10 | 3.20E-01 | -0.09 | 3.55E-01 | -0.24 | 1.48E-02 | -0.22 | 2.32E-02 | -0.10 | 3.09E-01 | -0.12 | 2.42E-01 | 0.13 | 2.08E-01 | -0.26 | 6.11E-03 | -0.09 | 3.88E-01 |
| | | Phospholipids in very large VLDL | 0.05 | 6.04E-01 | 0.24 | 1.52E-02 | -0.08 | 4.34E-01 | -0.13 | 1.80E-01 | -0.10 | 2.78E-01 | -0.27 | 5.62E-03 | -0.24 | 1.14E-02 | -0.12 | 2.07E-01 | -0.16 | 1.11E-01 | 0.14 | 1.74E-01 | -0.26 | 6.30E-03 | -0.08 | 3.89E-01 |
| | | Cholesterol in very large VLDL | 0.04 | 7.02E-01 | 0.22 | 2.66E-02 | -0.12 | 2.01E-01 | -0.17 | 7.48E-02 | -0.14 | 1.45E-01 | -0.29 | 3.02E-03 | -0.26 | 6.17E-03 | -0.14 | 1.58E-01 | -0.19 | 6.11E-02 | 0.12 | 2.20E-01 | -0.28 | 3.77E-03 | -0.08 | 3.96E-01 |
| | | CEs in very large VLDL | 0.02 | 8.34E-01 | 0.20 | 4.29E-02 | -0.15 | 1.28E-01 | -0.18 | 6.17E-02 | -0.16 | 1.03E-01 | -0.29 | 2.89E-03 | -0.26 | 7.09E-03 | -0.13 | 1.75E-01 | -0.19 | 5.42E-02 | 0.12 | 2.58E-01 | -0.28 | 3.28E-03 | -0.08 | 4.07E-01 |
| | | Free cholesterol in very large VLDL | 0.06 | 5.53E-01 | 0.24 | 1.52E-02 | -0.09 | 3.47E-01 | -0.16 | 1.00E-01 | -0.12 | 2.30E-01 | -0.28 | 3.66E-03 | -0.26 | 5.93E-03 | -0.14 | 1.45E-01 | -0.18 | 7.63E-02 | 0.13 | 1.89E-01 | -0.27 | 5.13E-03 | -0.09 | 3.87E-01 |
| | | TGs in very large VLDL | 0.08 | 3.93E-01 | 0.24 | 1.76E-02 | -0.05 | 5.89E-01 | -0.04 | 6.57E-01 | -0.06 | 5.63E-01 | -0.19 | 4.93E-02 | -0.18 | 6.37E-02 | -0.07 | 4.85E-01 | -0.06 | 5.36E-01 | 0.12 | 2.33E-01 | -0.24 | 1.12E-02 | -0.08 | 4.00E-01 |
| | Large VLDL (average diameter 53.6 nm) | Conc. of large VLDL particles | 0.05 | 5.95E-01 | 0.22 | 2.61E-02 | -0.12 | 2.15E-01 | -0.09 | 3.63E-01 | -0.14 | 1.45E-01 | -0.23 | 1.64E-02 | -0.23 | 1.78E-02 | -0.08 | 4.05E-01 | -0.16 | 1.16E-01 | 0.15 | 1.47E-01 | -0.27 | 3.85E-03 | -0.06 | 5.20E-01 |
| | | Total lipids in large VLDL | 0.07 | 4.81E-01 | 0.24 | 1.81E-02 | -0.11 | 2.38E-01 | -0.08 | 4.06E-01 | -0.13 | 1.91E-01 | -0.20 | 3.94E-02 | -0.23 | 1.86E-02 | -0.09 | 3.77E-01 | -0.12 | 2.09E-01 | 0.14 | 1.72E-01 | -0.28 | 3.55E-03 | -0.07 | 4.73E-01 |
| | | Phospholipids in large VLDL | 0.03 | 7.61E-01 | 0.21 | 3.38E-02 | -0.11 | 2.43E-01 | -0.07 | 4.92E-01 | -0.15 | 1.32E-01 | -0.23 | 1.60E-02 | -0.21 | 3.11E-02 | -0.07 | 4.86E-01 | -0.14 | 1.47E-01 | 0.16 | 1.19E-01 | -0.26 | 6.10E-03 | -0.06 | 5.31E-01 |
| | | Cholesterol in large VLDL | 0.04 | 7.01E-01 | 0.22 | 3.02E-02 | -0.14 | 1.41E-01 | -0.13 | 1.88E-01 | -0.16 | 8.68E-02 | -0.25 | 9.89E-03 | -0.24 | 1.05E-02 | -0.11 | 2.42E-01 | -0.18 | 7.22E-02 | 0.15 | 1.51E-01 | -0.26 | 5.56E-03 | -0.06 | 5.12E-01 |
| | | CEs in large VLDL | 0.03 | 7.34E-01 | 0.22 | 3.06E-02 | -0.16 | 1.06E-01 | -0.16 | 9.49E-02 | -0.18 | 6.03E-02 | -0.27 | 5.90E-03 | -0.26 | 6.08E-03 | -0.14 | 1.40E-01 | -0.20 | 4.19E-02 | 0.14 | 1.66E-01 | -0.26 | 7.22E-03 | -0.07 | 4.69E-01 |
| | | Free cholesterol in large VLDL | 0.04 | 6.68E-01 | 0.21 | 3.18E-02 | -0.12 | 1.97E-01 | -0.09 | 3.59E-01 | -0.14 | 1.33E-01 | -0.23 | 1.84E-02 | -0.22 | 2.08E-02 | -0.08 | 4.15E-01 | -0.15 | 1.32E-01 | 0.15 | 1.40E-01 | -0.27 | 4.87E-03 | -0.06 | 5.63E-01 |
| | | TGs in large VLDL | 0.11 | 2.84E-01 | 0.25 | 1.38E-02 | -0.09 | 3.54E-01 | -0.05 | 6.04E-01 | -0.08 | 3.83E-01 | -0.14 | 1.67E-01 | -0.21 | 3.04E-02 | -0.07 | 4.69E-01 | -0.07 | 4.82E-01 | 0.12 | 2.59E-01 | -0.27 | 3.81E-03 | -0.07 | 4.51E-01 |
| | Medium VLDL (average diameter 44.5 nm) | Conc. of medium VLDL particles | 0.03 | 7.69E-01 | 0.18 | 7.56E-02 | -0.22 | 2.42E-02 | -0.26 | 6.25E-03 | -0.20 | 3.87E-02 | -0.28 | 3.43E-03 | -0.27 | 4.07E-03 | -0.18 | 6.66E-02 | -0.22 | 2.82E-02 | 0.10 | 3.36E-01 | -0.23 | 1.63E-02 | -0.02 | 8.18E-01 |
| | | Total lipids in medium VLDL | 0.05 | 6.41E-01 | 0.19 | 5.25E-02 | -0.21 | 3.00E-02 | -0.22 | 2.38E-02 | -0.18 | 5.57E-02 | -0.26 | 7.59E-03 | -0.27 | 5.20E-03 | -0.14 | 1.39E-01 | -0.19 | 5.95E-02 | 0.12 | 2.22E-01 | -0.25 | 7.56E-03 | -0.02 | 8.08E-01 |
| | | Phospholipids in medium VLDL | 0.02 | 8.51E-01 | 0.17 | 9.08E-02 | -0.22 | 2.47E-02 | -0.28 | 3.89E-03 | -0.20 | 3.96E-02 | -0.29 | 3.04E-03 | -0.27 | 4.09E-03 | -0.19 | 5.61E-02 | -0.22 | 2.57E-02 | 0.10 | 3.52E-01 | -0.22 | 2.32E-02 | -0.02 | 8.57E-01 |
| | | Cholesterol in medium VLDL | 0.01 | 9.09E-01 | 0.13 | 2.08E-01 | -0.21 | 2.55E-02 | -0.38 | 4.98E-05 | -0.19 | 5.38E-02 | -0.29 | 2.40E-03 | -0.27 | 4.98E-03 | -0.25 | 9.96E-03 | -0.22 | 2.32E-02 | 0.05 | 6.36E-01 | -0.15 | 1.19E-01 | -0.01 | 9.34E-01 |
| | | CEs in medium VLDL | 0.01 | 9.56E-01 | 0.09 | 3.89E-01 | -0.21 | 3.11E-02 | -0.43 | 5.10E-06 | -0.17 | 6.88E-02 | -0.28 | 4.31E-03 | -0.23 | 1.44E-02 | -0.27 | 5.28E-03 | -0.20 | 3.83E-02 | 0.01 | 9.02E-01 | -0.09 | 3.44E-01 | 0.00 | 9.88E-01 |
| | | Free cholesterol in medium VLDL | 0.02 | 8.60E-01 | 0.16 | 1.07E-01 | -0.21 | 3.08E-02 | -0.31 | 1.27E-03 | -0.18 | 5.56E-02 | -0.29 | 2.46E-03 | -0.29 | 2.51E-03 | -0.21 | 3.03E-02 | -0.23 | 1.88E-02 | 0.08 | 4.08E-01 | -0.21 | 3.00E-02 | -0.02 | 8.53E-01 |
| | | TGs in medium VLDL | 0.07 | 5.03E-01 | 0.21 | 3.40E-02 | -0.16 | 8.94E-02 | -0.08 | 4.24E-01 | -0.14 | 1.35E-01 | -0.19 | 4.75E-02 | -0.21 | 2.58E-02 | -0.05 | 5.91E-01 | -0.12 | 2.45E-01 | 0.16 | 1.19E-01 | -0.28 | 3.36E-03 | -0.03 | 7.48E-01 |
| | Small VLDL (average diameter 36.8 nm) | Conc. of small VLDL particles | -0.02 | 8.51E-01 | 0.14 | 1.53E-01 | -0.19 | 5.06E-02 | -0.12 | 2.06E-01 | -0.17 | 6.97E-02 | -0.27 | 4.82E-03 | -0.20 | 4.00E-02 | -0.06 | 5.45E-01 | -0.18 | 7.53E-02 | 0.17 | 8.93E-02 | -0.23 | 1.64E-02 | 0.00 | 9.60E-01 |
| | | Total lipids in small VLDL | -0.01 | 9.36E-01 | 0.16 | 1.20E-01 | -0.20 | 4.08E-02 | -0.13 | 1.82E-01 | -0.18 | 6.05E-02 | -0.27 | 4.38E-03 | -0.20 | 3.26E-02 | -0.06 | 5.23E-01 | -0.18 | 6.75E-02 | 0.18 | 8.19E-02 | -0.24 | 1.32E-02 | 0.00 | 9.89E-01 |
| | | Phospholipids in small VLDL | -0.01 | 9.29E-01 | 0.15 | 1.32E-01 | -0.21 | 2.49E-02 | -0.19 | 4.55E-02 | -0.20 | 3.96E-02 | -0.29 | 2.26E-03 | -0.22 | 2.15E-02 | -0.10 | 3.12E-01 | -0.20 | 4.46E-02 | 0.15 | 1.43E-01 | -0.22 | 1.95E-02 | 0.01 | 9.54E-01 |
| | | Cholesterol in small VLDL | -0.01 | 8.91E-01 | 0.15 | 1.38E-01 | -0.20 | 3.60E-02 | -0.21 | 2.95E-02 | -0.20 | 3.64E-02 | -0.31 | 1.37E-03 | -0.23 | 1.66E-02 | -0.12 | 2.13E-01 | -0.22 | 2.28E-02 | 0.14 | 1.79E-01 | -0.22 | 2.33E-02 | -0.01 | 9.56E-01 |
| | | CEs in small VLDL | -0.02 | 8.47E-01 | 0.15 | 1.37E-01 | -0.19 | 5.09E-02 | -0.19 | 5.61E-02 | -0.20 | 3.86E-02 | -0.31 | 1.32E-03 | -0.22 | 2.10E-02 | -0.11 | 2.62E-01 | -0.23 | 2.18E-02 | 0.15 | 1.52E-01 | -0.22 | 2.18E-02 | -0.01 | 8.88E-01 |
| | | Free cholesterol in small VLDL | 0.00 | 9.82E-01 | 0.15 | 1.48E-01 | -0.23 | 1.85E-02 | -0.26 | 7.34E-03 | -0.20 | 3.46E-02 | -0.30 | 1.82E-03 | -0.24 | 1.13E-02 | -0.14 | 1.42E-01 | -0.22 | 2.69E-02 | 0.12 | 2.58E-01 | -0.21 | 2.86E-02 | 0.01 | 9.01E-01 |
| | | TGs in small VLDL | 0.00 | 9.86E-01 | 0.15 | 1.31E-01 | -0.17 | 8.02E-02 | -0.04 | 7.20E-01 | -0.14 | 1.35E-01 | -0.23 | 1.99E-02 | -0.16 | 8.70E-02 | 0.00 | 9.94E-01 | -0.13 | 2.04E-01 | 0.21 | 4.13E-02 | -0.24 | 1.25E-02 | 0.00 | 9.73E-01 |
| | Very small VLDL (average diameter 31.3 nm) | Conc. of very small VLDL particles | -0.04 | 7.11E-01 | 0.13 | 2.03E-01 | -0.17 | 7.59E-02 | -0.27 | 4.58E-03 | -0.18 | 5.81E-02 | -0.32 | 8.34E-04 | -0.24 | 1.04E-02 | -0.13 | 1.86E-01 | -0.28 | 4.26E-03 | 0.14 | 1.61E-01 | -0.15 | 1.22E-01 | 0.03 | 7.51E-01 |
| | | Total lipids in very small VLDL | -0.04 | 7.23E-01 | 0.13 | 2.13E-01 | -0.17 | 8.16E-02 | -0.27 | 4.93E-03 | -0.17 | 7.39E-02 | -0.32 | 7.88E-04 | -0.25 | 8.86E-03 | -0.14 | 1.59E-01 | -0.27 | 5.10E-03 | 0.15 | 1.41E-01 | -0.15 | 1.13E-01 | 0.03 | 7.68E-01 |
| | | Phospholipids in very small VLDL | -0.04 | 7.07E-01 | 0.14 | 1.53E-01 | -0.15 | 1.28E-01 | -0.25 | 8.62E-03 | -0.17 | 7.84E-02 | -0.34 | 3.22E-04 | -0.25 | 9.12E-03 | -0.13 | 1.87E-01 | -0.27 | 5.60E-03 | 0.15 | 1.35E-01 | -0.16 | 9.39E-02 | 0.00 | 1.00E+00 |
| | | Cholesterol in very small VLDL | -0.01 | 9.04E-01 | 0.09 | 3.74E-01 | -0.14 | 1.42E-01 | -0.38 | 6.09E-05 | -0.14 | 1.38E-01 | -0.26 | 6.31E-03 | -0.24 | 1.04E-02 | -0.23 | 1.74E-02 | -0.27 | 5.79E-03 | 0.05 | 6.40E-01 | -0.08 | 4.12E-01 | 0.03 | 7.24E-01 |
| | | CEs in very small VLDL | 0.00 | 9.81E-01 | 0.05 | 5.97E-01 | -0.13 | 1.88E-01 | -0.39 | 4.17E-05 | -0.13 | 1.94E-01 | -0.22 | 2.67E-02 | -0.22 | 1.93E-02 | -0.24 | 1.14E-02 | -0.25 | 1.03E-02 | 0.01 | 9.40E-01 | -0.04 | 6.49E-01 | 0.04 | 6.97E-01 |
| | | Free cholesterol in very small VLDL | -0.03 | 7.57E-01 | 0.14 | 1.77E-01 | -0.16 | 1.04E-01 | -0.30 | 1.77E-03 | -0.16 | 9.17E-02 | -0.32 | 8.81E-04 | -0.26 | 5.44E-03 | -0.17 | 8.32E-02 | -0.28 | 3.71E-03 | 0.12 | 2.22E-01 | -0.15 | 1.28E-01 | 0.02 | 8.48E-01 |
| | | TGs in very small VLDL | -0.06 | 5.75E-01 | 0.11 | 2.75E-01 | -0.16 | 9.50E-02 | -0.06 | 5.75E-01 | -0.15 | 1.11E-01 | -0.26 | 8.25E-03 | -0.16 | 9.93E-02 | 0.02 | 8.12E-01 | -0.18 | 7.49E-02 | 0.21 | 3.38E-02 | -0.20 | 3.67E-02 | 0.04 | 6.90E-01 |
| | IDL (average diameter 28.6 nm) | Conc. of IDL particles | -0.02 | 8.53E-01 | 0.05 | 6.39E-01 | -0.16 | 9.47E-02 | -0.40 | 2.67E-05 | -0.16 | 9.29E-02 | -0.18 | 6.98E-02 | -0.21 | 2.84E-02 | -0.19 | 5.43E-02 | -0.22 | 2.63E-02 | -0.03 | 7.88E-01 | -0.06 | 5.26E-01 | 0.05 | 5.88E-01 |
| | | Total lipids in IDL | 0.00 | 9.94E-01 | -0.04 | 7.28E-01 | -0.18 | 5.98E-02 | -0.34 | 3.00E-04 | -0.12 | 2.24E-01 | -0.10 | 2.91E-01 | -0.18 | 6.09E-02 | -0.16 | 1.03E-01 | -0.19 | 5.97E-02 | -0.02 | 8.32E-01 | -0.03 | 7.69E-01 | 0.13 | 2.02E-01 |
| | | Phospholipids in IDL | -0.01 | 9.33E-01 | -0.01 | 9.32E-01 | -0.18 | 6.39E-02 | -0.36 | 1.58E-04 | -0.13 | 1.72E-01 | -0.15 | 1.36E-01 | -0.17 | 7.45E-02 | -0.18 | 5.95E-02 | -0.19 | 5.12E-02 | -0.02 | 8.47E-01 | -0.02 | 8.36E-01 | 0.10 | 2.92E-01 |
| | | Cholesterol in IDL | 0.01 | 8.86E-01 | -0.06 | 5.53E-01 | -0.16 | 9.36E-02 | -0.33 | 5.63E-04 | -0.09 | 3.38E-01 | -0.04 | 6.48E-01 | -0.17 | 8.55E-02 | -0.16 | 9.88E-02 | -0.16 | 1.05E-01 | -0.06 | 5.73E-01 | -0.01 | 9.59E-01 | 0.12 | 2.28E-01 |
| | | CEs in IDL | 0.02 | 8.65E-01 | -0.07 | 4.88E-01 | -0.16 | 9.68E-02 | -0.32 | 8.10E-04 | -0.10 | 3.24E-01 | -0.03 | 7.55E-01 | -0.17 | 8.56E-02 | -0.15 | 1.16E-01 | -0.16 | 1.11E-01 | -0.06 | 5.66E-01 | -0.01 | 9.52E-01 | 0.12 | 2.21E-01 |
| | | Free cholesterol in IDL | 0.00 | 9.65E-01 | -0.03 | 7.67E-01 | -0.16 | 9.48E-02 | -0.35 | 2.09E-04 | -0.08 | 3.93E-01 | -0.09 | 3.73E-01 | -0.17 | 8.32E-02 | -0.18 | 6.27E-02 | -0.17 | 8.61E-02 | -0.05 | 6.14E-01 | 0.00 | 9.75E-01 | 0.11 | 2.62E-01 |
| | | TGs in IDL | -0.07 | 4.58E-01 | 0.08 | 4.11E-01 | -0.17 | 6.96E-02 | -0.08 | 3.87E-01 | -0.16 | 1.07E-01 | -0.24 | 1.15E-02 | -0.16 | 1.04E-01 | 0.03 | 7.38E-01 | -0.20 | 4.35E-02 | 0.19 | 5.94E-02 | -0.17 | 8.22E-02 | 0.08 | 4.25E-01 |
| | Large LDL (average diameter 25.5 nm) | Conc. of large LDL particles | -0.03 | 7.88E-01 | 0.05 | 6.54E-01 | -0.22 | 1.92E-02 | -0.36 | 1.77E-04 | -0.18 | 5.81E-02 | -0.29 | 2.80E-03 | -0.20 | 3.44E-02 | -0.21 | 3.46E-02 | -0.21 | 3.55E-02 | -0.02 | 8.78E-01 | -0.11 | 2.68E-01 | 0.07 | 4.98E-01 |
| | | Total lipids in large LDL | -0.01 | 9.29E-01 | -0.02 | 8.39E-01 | -0.26 | 6.96E-03 | -0.28 | 3.85E-03 | -0.16 | 9.88E-02 | -0.08 | 4.31E-01 | -0.12 | 2.04E-01 | -0.10 | 3.16E-01 | -0.13 | 1.82E-01 | -0.01 | 9.15E-01 | -0.04 | 6.73E-01 | 0.17 | 8.83E-02 |
| | | Phospholipids in large LDL | -0.01 | 9.52E-01 | -0.03 | 7.97E-01 | -0.25 | 8.86E-03 | -0.29 | 2.82E-03 | -0.16 | 1.03E-01 | -0.07 | 5.01E-01 | -0.14 | 1.60E-01 | -0.10 | 2.89E-01 | -0.14 | 1.54E-01 | -0.02 | 8.69E-01 | -0.05 | 6.14E-01 | 0.16 | 1.06E-01 |
| | | Cholesterol in large LDL | 0.00 | 9.95E-01 | -0.03 | 7.77E-01 | -0.25 | 8.28E-03 | -0.28 | 3.76E-03 | -0.15 | 1.15E-01 | -0.05 | 5.79E-01 | -0.11 | 2.54E-01 | -0.11 | 2.71E-01 | -0.12 | 2.37E-01 | -0.03 | 7.60E-01 | -0.02 | 8.08E-01 | 0.16 | 9.35E-02 |
| | | CEs in large LDL | 0.00 | 9.91E-01 | -0.02 | 8.77E-01 | -0.26 | 5.80E-03 | -0.27 | 4.64E-03 | -0.16 | 9.07E-02 | -0.07 | 4.81E-01 | -0.11 | 2.45E-01 | -0.10 | 2.97E-01 | -0.12 | 2.34E-01 | -0.02 | 8.54E-01 | -0.03 | 7.31E-01 | 0.16 | 9.38E-02 |
| | | Free cholesterol in large LDL | 0.00 | 9.87E-01 | -0.06 | 5.36E-01 | -0.21 | 2.52E-02 | -0.29 | 3.06E-03 | -0.12 | 2.20E-01 | -0.01 | 8.98E-01 | -0.10 | 2.92E-01 | -0.12 | 2.10E-01 | -0.11 | 2.58E-01 | -0.07 | 5.24E-01 | 0.00 | 9.83E-01 | 0.16 | 1.10E-01 |
| | | TGs in large LDL | -0.07 | 4.69E-01 | 0.06 | 5.75E-01 | -0.20 | 3.98E-02 | -0.09 | 3.69E-01 | -0.15 | 1.21E-01 | -0.21 | 3.03E-02 | -0.14 | 1.41E-01 | 0.05 | 6.30E-01 | -0.18 | 6.20E-02 | 0.16 | 1.17E-01 | -0.15 | 1.31E-01 | 0.11 | 2.54E-01 |
| | Medium LDL (average diameter 23 nm) | Conc. of medium LDL particles | 0.02 | 8.77E-01 | 0.12 | 2.16E-01 | -0.24 | 1.13E-02 | -0.28 | 3.29E-03 | -0.17 | 7.37E-02 | -0.17 | 7.59E-02 | -0.20 | 3.28E-02 | -0.13 | 1.93E-01 | -0.18 | 6.99E-02 | 0.08 | 4.26E-01 | -0.16 | 1.02E-01 | 0.07 | 4.68E-01 |
| | | Total lipids in medium LDL | -0.02 | 8.51E-01 | 0.04 | 7.01E-01 | -0.29 | 2.51E-03 | -0.23 | 1.77E-02 | -0.20 | 3.39E-02 | -0.14 | 1.65E-01 | -0.15 | 1.30E-01 | -0.06 | 5.15E-01 | -0.14 | 1.73E-01 | 0.05 | 6.45E-01 | -0.13 | 1.70E-01 | 0.14 | 1.59E-01 |
| | | Phospholipids in medium LDL | -0.02 | 8.19E-01 | 0.02 | 8.44E-01 | -0.29 | 2.57E-03 | -0.23 | 1.78E-02 | -0.21 | 3.01E-02 | -0.13 | 1.84E-01 | -0.14 | 1.50E-01 | -0.06 | 5.58E-01 | -0.13 | 1.82E-01 | 0.03 | 7.68E-01 | -0.13 | 1.76E-01 | 0.14 | 1.49E-01 |
| | | Cholesterol in medium LDL | -0.01 | 9.03E-01 | 0.04 | 6.98E-01 | -0.29 | 2.61E-03 | -0.24 | 1.37E-02 | -0.20 | 3.54E-02 | -0.12 | 2.19E-01 | -0.14 | 1.42E-01 | -0.08 | 4.25E-01 | -0.13 | 2.02E-01 | 0.03 | 7.44E-01 | -0.12 | 2.04E-01 | 0.14 | 1.68E-01 |
| | | CEs in medium LDL | -0.01 | 9.11E-01 | 0.07 | 5.21E-01 | -0.28 | 2.78E-03 | -0.22 | 2.05E-02 | -0.21 | 2.86E-02 | -0.14 | 1.43E-01 | -0.15 | 1.10E-01 | -0.07 | 4.58E-01 | -0.13 | 1.79E-01 | 0.06 | 5.51E-01 | -0.15 | 1.29E-01 | 0.12 | 2.30E-01 |
| | | Free cholesterol in medium LDL | -0.01 | 8.81E-01 | -0.03 | 7.42E-01 | -0.27 | 4.74E-03 | -0.25 | 8.42E-03 | -0.17 | 8.44E-02 | -0.05 | 6.35E-01 | -0.10 | 3.07E-01 | -0.08 | 3.96E-01 | -0.10 | 3.15E-01 | -0.04 | 7.24E-01 | -0.06 | 5.68E-01 | 0.17 | 8.48E-02 |
| | | TGs in medium LDL | -0.05 | 6.00E-01 | 0.07 | 4.69E-01 | -0.21 | 2.91E-02 | -0.09 | 3.44E-01 | -0.14 | 1.42E-01 | -0.22 | 2.05E-02 | -0.16 | 1.03E-01 | 0.04 | 7.08E-01 | -0.17 | 8.62E-02 | 0.16 | 1.06E-01 | -0.17 | 7.21E-02 | 0.10 | 3.00E-01 |
| | Small LDL (average diameter 18.7 nm) | Conc. of small LDL particles | 0.00 | 9.77E-01 | 0.14 | 1.77E-01 | -0.19 | 4.64E-02 | -0.34 | 4.13E-04 | -0.17 | 8.07E-02 | -0.25 | 1.13E-02 | -0.24 | 1.03E-02 | -0.20 | 4.49E-02 | -0.23 | 1.69E-02 | 0.03 | 7.66E-01 | -0.15 | 1.13E-01 | 0.03 | 7.64E-01 |
| | | Total lipids in small LDL | -0.01 | 9.30E-01 | 0.05 | 6.46E-01 | -0.24 | 1.28E-02 | -0.33 | 6.62E-04 | -0.17 | 8.24E-02 | -0.21 | 3.43E-02 | -0.19 | 4.77E-02 | -0.15 | 1.18E-01 | -0.17 | 8.91E-02 | -0.02 | 8.44E-01 | -0.13 | 1.66E-01 | 0.10 | 3.17E-01 |
| | | Phospholipids in small LDL | -0.02 | 8.33E-01 | 0.02 | 8.77E-01 | -0.16 | 9.01E-02 | -0.34 | 2.98E-04 | -0.14 | 1.54E-01 | -0.21 | 2.95E-02 | -0.19 | 4.53E-02 | -0.21 | 2.84E-02 | -0.18 | 6.32E-02 | -0.10 | 3.10E-01 | -0.04 | 6.73E-01 | 0.06 | 5.15E-01 |
| | | Cholesterol in small LDL | 0.00 | 9.62E-01 | 0.04 | 6.90E-01 | -0.25 | 9.06E-03 | -0.31 | 1.13E-03 | -0.17 | 8.25E-02 | -0.17 | 8.95E-02 | -0.17 | 7.91E-02 | -0.13 | 1.81E-01 | -0.15 | 1.40E-01 | -0.01 | 8.91E-01 | -0.14 | 1.36E-01 | 0.11 | 2.51E-01 |
| | | CEs in small LDL | 0.01 | 9.46E-01 | 0.07 | 4.97E-01 | -0.26 | 6.01E-03 | -0.31 | 1.26E-03 | -0.18 | 6.69E-02 | -0.19 | 4.95E-02 | -0.19 | 4.90E-02 | -0.13 | 2.00E-01 | -0.16 | 1.14E-01 | 0.02 | 8.42E-01 | -0.16 | 9.80E-02 | 0.10 | 2.93E-01 |
| | | Free cholesterol in small LDL | -0.03 | 7.28E-01 | -0.05 | 6.20E-01 | -0.20 | 3.96E-02 | -0.29 | 2.92E-03 | -0.13 | 1.75E-01 | -0.07 | 5.02E-01 | -0.10 | 2.95E-01 | -0.13 | 1.76E-01 | -0.11 | 2.73E-01 | -0.12 | 2.53E-01 | -0.09 | 3.57E-01 | 0.13 | 1.85E-01 |
| | | TGs in small LDL | 0.00 | 9.96E-01 | 0.14 | 1.65E-01 | -0.19 | 4.38E-02 | -0.15 | 1.38E-01 | -0.13 | 1.90E-01 | -0.29 | 2.20E-03 | -0.20 | 3.55E-02 | -0.02 | 8.40E-01 | -0.17 | 8.45E-02 | 0.17 | 8.86E-02 | -0.22 | 2.15E-02 | 0.04 | 6.53E-01 |
| | Very large HDL (average diameter 14.3 nm) | Conc. of very large HDL particles | 0.02 | 8.43E-01 | -0.07 | 4.63E-01 | -0.02 | 8.25E-01 | -0.24 | 1.40E-02 | 0.10 | 2.80E-01 | 0.00 | 9.99E-01 | -0.12 | 1.97E-01 | -0.15 | 1.31E-01 | -0.09 | 3.63E-01 | -0.11 | 2.93E-01 | 0.08 | 3.82E-01 | 0.08 | 3.97E-01 |
| | | Total lipids in very large HDL | 0.02 | 8.45E-01 | -0.08 | 4.46E-01 | 0.01 | 9.20E-01 | -0.22 | 2.07E-02 | 0.12 | 2.19E-01 | 0.01 | 8.88E-01 | -0.11 | 2.51E-01 | -0.15 | 1.33E-01 | -0.08 | 4.36E-01 | -0.12 | 2.44E-01 | 0.10 | 2.95E-01 | 0.07 | 4.95E-01 |
| | | Phospholipids in very large HDL | 0.02 | 8.28E-01 | -0.08 | 4.49E-01 | 0.02 | 8.07E-01 | -0.22 | 2.32E-02 | 0.12 | 2.01E-01 | 0.01 | 8.94E-01 | -0.11 | 2.50E-01 | -0.15 | 1.31E-01 | -0.08 | 4.40E-01 | -0.11 | 2.65E-01 | 0.10 | 2.91E-01 | 0.06 | 5.47E-01 |
| | | Cholesterol in very large HDL | 0.02 | 8.40E-01 | -0.08 | 4.07E-01 | 0.00 | 9.75E-01 | -0.22 | 2.37E-02 | 0.12 | 2.18E-01 | 0.03 | 7.73E-01 | -0.10 | 3.08E-01 | -0.14 | 1.49E-01 | -0.07 | 5.02E-01 | -0.13 | 2.00E-01 | 0.11 | 2.56E-01 | 0.07 | 4.61E-01 |
| | | CEs in very large HDL | 0.02 | 8.37E-01 | -0.09 | 3.97E-01 | -0.01 | 9.21E-01 | -0.23 | 1.83E-02 | 0.12 | 2.24E-01 | 0.02 | 8.34E-01 | -0.10 | 2.81E-01 | -0.14 | 1.60E-01 | -0.06 | 5.20E-01 | -0.12 | 2.30E-01 | 0.10 | 2.86E-01 | 0.08 | 4.06E-01 |
| | | Free cholesterol in very large HDL | 0.02 | 8.71E-01 | -0.08 | 4.40E-01 | 0.04 | 6.81E-01 | -0.19 | 5.15E-02 | 0.12 | 2.13E-01 | 0.05 | 6.01E-01 | -0.08 | 3.97E-01 | -0.15 | 1.17E-01 | -0.07 | 4.54E-01 | -0.16 | 1.25E-01 | 0.12 | 1.95E-01 | 0.04 | 6.72E-01 |
| | | TGs in very large HDL | 0.02 | 8.37E-01 | 0.09 | 3.48E-01 | -0.19 | 4.99E-02 | -0.27 | 5.47E-03 | -0.06 | 5.69E-01 | -0.27 | 4.42E-03 | -0.26 | 6.09E-03 | -0.13 | 1.85E-01 | -0.23 | 1.83E-02 | 0.12 | 2.38E-01 | -0.14 | 1.48E-01 | 0.08 | 4.06E-01 |
| | Large HDL (average diameter 12.1 nm) | Conc. of large HDL particles | 0.03 | 7.29E-01 | -0.11 | 2.95E-01 | -0.08 | 4.23E-01 | -0.21 | 3.01E-02 | 0.09 | 3.56E-01 | 0.05 | 6.27E-01 | -0.10 | 3.04E-01 | -0.09 | 3.44E-01 | -0.05 | 6.18E-01 | -0.07 | 5.23E-01 | 0.09 | 3.57E-01 | 0.13 | 1.75E-01 |
| | | Total lipids in large HDL | 0.03 | 7.32E-01 | -0.11 | 2.71E-01 | -0.07 | 4.50E-01 | -0.20 | 3.83E-02 | 0.09 | 3.67E-01 | 0.06 | 5.41E-01 | -0.09 | 3.62E-01 | -0.08 | 4.13E-01 | -0.04 | 6.78E-01 | -0.07 | 5.18E-01 | 0.10 | 3.17E-01 | 0.14 | 1.67E-01 |
| | | Phospholipids in large HDL | 0.03 | 7.29E-01 | -0.12 | 2.48E-01 | -0.08 | 4.13E-01 | -0.19 | 4.96E-02 | 0.08 | 4.13E-01 | 0.07 | 4.78E-01 | -0.08 | 3.92E-01 | -0.06 | 5.28E-01 | -0.04 | 7.06E-01 | -0.06 | 5.75E-01 | 0.10 | 3.15E-01 | 0.15 | 1.39E-01 |
| | | Cholesterol in large HDL | 0.03 | 7.44E-01 | -0.11 | 2.70E-01 | -0.06 | 5.46E-01 | -0.20 | 3.65E-02 | 0.10 | 2.91E-01 | 0.06 | 5.30E-01 | -0.08 | 3.98E-01 | -0.09 | 3.41E-01 | -0.03 | 7.43E-01 | -0.08 | 4.07E-01 | 0.11 | 2.70E-01 | 0.12 | 2.07E-01 |
| | | CEs in large HDL | 0.03 | 7.34E-01 | -0.11 | 2.65E-01 | -0.06 | 5.29E-01 | -0.20 | 3.79E-02 | 0.10 | 2.80E-01 | 0.06 | 5.08E-01 | -0.08 | 4.12E-01 | -0.09 | 3.51E-01 | -0.03 | 7.77E-01 | -0.09 | 4.01E-01 | 0.11 | 2.66E-01 | 0.13 | 2.01E-01 |
| | | Free cholesterol in large HDL | 0.03 | 7.76E-01 | 0.04 | 7.02E-01 | -0.05 | 6.07E-01 | -0.21 | 3.10E-02 | 0.09 | 3.29E-01 | -0.14 | 1.53E-01 | -0.09 | 3.52E-01 | -0.09 | 3.47E-01 | -0.05 | 6.16E-01 | 0.15 | 1.53E-01 | 0.10 | 2.90E-01 | 0.12 | 2.06E-01 |
| | | TGs in large HDL | 0.04 | 7.00E-01 | -0.12 | 2.22E-01 | -0.20 | 3.75E-02 | -0.13 | 2.00E-01 | -0.07 | 4.74E-01 | 0.12 | 2.26E-01 | -0.24 | 1.25E-02 | 0.06 | 5.48E-01 | -0.22 | 2.61E-02 | 0.06 | 5.70E-01 | -0.11 | 2.77E-01 | 0.24 | 1.19E-02 |
| | Medium HDL (average diameter 10.9 nm) | Conc. of medium HDL particles | 0.06 | 5.72E-01 | -0.12 | 2.23E-01 | -0.26 | 6.25E-03 | -0.11 | 2.73E-01 | -0.05 | 6.32E-01 | 0.13 | 1.97E-01 | -0.06 | 5.02E-01 | 0.08 | 4.15E-01 | 0.00 | 9.97E-01 | 0.07 | 5.00E-01 | 0.02 | 8.31E-01 | 0.25 | 1.01E-02 |
| | | Total lipids in medium HDL | 0.06 | 5.74E-01 | -0.12 | 2.53E-01 | -0.27 | 4.86E-03 | -0.10 | 3.20E-01 | -0.06 | 5.57E-01 | 0.13 | 1.92E-01 | -0.05 | 5.75E-01 | 0.09 | 3.81E-01 | 0.00 | 9.60E-01 | 0.09 | 3.99E-01 | 0.02 | 8.43E-01 | 0.25 | 1.10E-02 |
| | | Phospholipids in medium HDL | 0.06 | 5.33E-01 | -0.15 | 1.47E-01 | -0.27 | 4.42E-03 | -0.11 | 2.65E-01 | -0.07 | 4.58E-01 | 0.15 | 1.23E-01 | -0.05 | 5.73E-01 | 0.07 | 4.53E-01 | 0.00 | 9.83E-01 | 0.02 | 8.28E-01 | 0.01 | 8.79E-01 | 0.25 | 1.12E-02 |
| | | Cholesterol in medium HDL | 0.05 | 6.16E-01 | -0.15 | 1.37E-01 | -0.24 | 1.09E-02 | -0.10 | 3.09E-01 | -0.02 | 8.37E-01 | 0.16 | 1.01E-01 | -0.03 | 7.49E-01 | 0.08 | 4.02E-01 | 0.03 | 7.50E-01 | 0.02 | 8.45E-01 | 0.05 | 5.93E-01 | 0.25 | 1.06E-02 |
| | | CEs in medium HDL | 0.05 | 6.16E-01 | -0.13 | 2.09E-01 | -0.25 | 9.98E-03 | -0.14 | 1.41E-01 | -0.02 | 8.52E-01 | 0.11 | 2.56E-01 | -0.02 | 8.24E-01 | 0.04 | 6.97E-01 | 0.04 | 6.56E-01 | 0.03 | 7.59E-01 | 0.05 | 5.71E-01 | 0.24 | 1.53E-02 |
| | | Free cholesterol in medium HDL | 0.04 | 6.72E-01 | 0.12 | 2.49E-01 | -0.23 | 1.52E-02 | -0.06 | 5.71E-01 | -0.03 | 7.73E-01 | -0.17 | 8.20E-02 | -0.07 | 4.90E-01 | 0.02 | 8.17E-01 | -0.01 | 8.87E-01 | 0.26 | 1.00E-02 | 0.04 | 6.89E-01 | 0.07 | 4.76E-01 |
| | Small HDL (average diameter 8.7 nm) | Conc. of small HDL particles | 0.03 | 7.82E-01 | -0.04 | 6.71E-01 | -0.35 | 2.00E-04 | 0.02 | 8.63E-01 | -0.20 | 3.31E-02 | 0.09 | 3.62E-01 | 0.01 | 9.24E-01 | -0.10 | 3.06E-01 | -0.21 | 3.15E-02 | -0.08 | 4.33E-01 | -0.24 | 1.35E-02 | 0.12 | 2.25E-01 |
| | | TGs in medium HDL | 0.03 | 7.72E-01 | -0.11 | 2.91E-01 | -0.24 | 1.21E-02 | -0.21 | 3.05E-02 | -0.21 | 2.61E-02 | 0.05 | 6.06E-01 | -0.21 | 3.01E-02 | 0.16 | 1.05E-01 | 0.05 | 6.24E-01 | 0.13 | 1.97E-01 | -0.10 | 3.10E-01 | 0.21 | 3.21E-02 |
| | | Total lipids in small HDL | 0.03 | 7.44E-01 | -0.03 | 7.80E-01 | -0.35 | 1.59E-04 | 0.01 | 9.19E-01 | -0.22 | 2.25E-02 | 0.07 | 5.04E-01 | -0.03 | 7.43E-01 | 0.15 | 1.19E-01 | 0.01 | 9.08E-01 | 0.16 | 1.08E-01 | -0.13 | 1.83E-01 | 0.21 | 3.57E-02 |
| | | Phospholipids in small HDL | 0.05 | 6.29E-01 | -0.03 | 7.42E-01 | -0.35 | 1.70E-04 | 0.00 | 9.64E-01 | -0.22 | 1.98E-02 | 0.08 | 3.95E-01 | -0.05 | 6.11E-01 | 0.14 | 1.44E-01 | 0.00 | 9.65E-01 | 0.17 | 9.92E-02 | -0.13 | 1.85E-01 | 0.20 | 3.82E-02 |
| | | Cholesterol in small HDL | 0.02 | 8.25E-01 | -0.06 | 5.41E-01 | -0.35 | 1.58E-04 | 0.01 | 8.85E-01 | -0.19 | 5.01E-02 | 0.10 | 2.91E-01 | 0.02 | 8.13E-01 | 0.17 | 7.54E-02 | 0.06 | 5.55E-01 | 0.12 | 2.32E-01 | -0.08 | 4.11E-01 | 0.23 | 1.66E-02 |

| Variable Group | Subgroup | Variable | TSH | | | | Apgar score, 1 min | | | | Apgar score, 5 min | | | | Child weight | | | | Head circumstance | | | | Placenta weight | | | |
|---|---|---|---|---|---|---|---|---|---|---|---|---|---|---|---|---|---|---|---|---|---|---|---|---|---|---|
| | | | Controls | | Levothyroxine | | Controls | | Levothyroxine | | Controls | | Levothyroxine | | Controls | | Levothyroxine | | Controls | | Levothyroxine | | Controls | | Levothyroxine | |
| | | | β | p | β | p | β | p | β | p | β | p | β | p | β | p | β | p | β | p | β | p | β | p | β | p |
| | | CEs in small HDL | 0.02 | 8.05E-01 | -0.07 | 4.99E-01 | -0.34 | 3.65E-04 | 0.05 | 6.01E-01 | -0.18 | 5.76E-02 | 0.14 | 1.55E-01 | 0.07 | 4.92E-01 | 0.19 | 4.77E-02 | 0.10 | 2.99E-01 | 0.11 | 2.79E-01 | -0.06 | 5.03E-01 | 0.22 | 2.35E-02 |
| | | Free cholesterol in small HDL | 0.01 | 9.45E-01 | -0.05 | 6.56E-01 | -0.35 | 2.05E-04 | -0.08 | 3.89E-01 | -0.17 | 7.26E-02 | 0.00 | 9.85E-01 | -0.09 | 3.69E-01 | 0.11 | 2.84E-01 | -0.06 | 5.69E-01 | 0.14 | 1.68E-01 | -0.11 | 2.73E-01 | 0.25 | 1.18E-02 |
| | | TGs in small HDL | -0.01 | 8.94E-01 | 0.13 | 2.02E-01 | -0.17 | 7.43E-02 | 0.02 | 8.73E-01 | -0.19 | 4.74E-02 | -0.19 | 5.45E-02 | -0.13 | 1.88E-01 | 0.05 | 5.78E-01 | -0.14 | 1.64E-01 | 0.22 | 3.01E-02 | -0.24 | 1.31E-02 | 0.02 | 8.31E-01 |
| Relative lipoprotein lipid concentrations | CMs and extremely large VLDL ratios | Phospholipids to total lipids ratio in CMs and extremely large VLDL | 0.06 | 6.18E-01 | 0.15 | 1.92E-01 | -0.13 | 2.46E-01 | 0.03 | 7.72E-01 | -0.12 | 2.65E-01 | -0.13 | 2.54E-01 | -0.06 | 5.49E-01 | 0.13 | 2.66E-01 | -0.14 | 2.20E-01 | 0.26 | 2.92E-02 | -0.15 | 1.76E-01 | 0.10 | 3.93E-01 |
| | | Cholesterol to total lipids ratio in CMs and extremely large VLDL | 0.01 | 9.16E-01 | 0.09 | 4.43E-01 | -0.22 | 4.35E-02 | -0.02 | 8.28E-01 | -0.23 | 3.49E-02 | -0.11 | 3.53E-01 | -0.15 | 1.58E-01 | 0.18 | 1.13E-01 | -0.28 | 1.15E-02 | 0.20 | 9.63E-02 | -0.17 | 1.13E-01 | 0.11 | 3.19E-01 |
| | | CEs to total lipids ratio in CMs and extremely large VLDL | 0.01 | 9.16E-01 | 0.08 | 4.85E-01 | -0.20 | 6.25E-02 | -0.06 | 6.24E-01 | -0.21 | 4.55E-02 | -0.14 | 2.19E-01 | -0.20 | 6.29E-02 | 0.14 | 2.17E-01 | -0.33 | 2.60E-03 | 0.10 | 3.86E-01 | -0.23 | 3.15E-02 | 0.07 | 5.23E-01 |
| | | Free cholesterol to total lipids ratio in CMs and extremely large VLDL | 0.01 | 9.22E-01 | 0.09 | 4.28E-01 | -0.22 | 4.33E-02 | 0.02 | 8.63E-01 | -0.22 | 4.16E-02 | -0.05 | 6.60E-01 | -0.06 | 5.52E-01 | 0.21 | 5.56E-02 | -0.17 | 1.22E-01 | 0.31 | 8.38E-03 | -0.06 | 5.66E-01 | 0.16 | 1.61E-01 |
| | | TGs to total lipids ratio in CMs and extremely large VLDL | -0.03 | 7.97E-01 | -0.12 | 3.14E-01 | 0.19 | 7.73E-02 | 0.00 | 9.79E-01 | 0.19 | 7.06E-02 | 0.12 | 2.96E-01 | 0.12 | 2.47E-01 | -0.16 | 1.48E-01 | 0.23 | 3.38E-02 | -0.23 | 5.48E-02 | 0.17 | 1.20E-01 | -0.11 | 3.30E-01 |
| | Very large VLDL ratios | Phospholipids to total lipids ratio in very large VLDL | 0.03 | 7.46E-01 | 0.20 | 5.13E-02 | -0.07 | 5.10E-01 | -0.14 | 1.75E-01 | -0.12 | 2.44E-01 | -0.14 | 1.62E-01 | -0.22 | 2.31E-02 | -0.06 | 5.89E-01 | -0.22 | 2.98E-02 | 0.23 | 2.80E-02 | -0.23 | 2.00E-02 | -0.04 | 7.17E-01 |
| | | Cholesterol to total lipids ratio in very large VLDL | -0.08 | 4.38E-01 | -0.08 | 4.52E-01 | -0.18 | 7.60E-02 | -0.23 | 2.42E-02 | -0.20 | 4.45E-02 | 0.03 | 7.67E-01 | -0.10 | 2.97E-01 | -0.06 | 5.43E-01 | -0.18 | 7.46E-02 | 0.01 | 9.25E-01 | -0.12 | 2.19E-01 | 0.07 | 5.24E-01 |
| | | CEs to total lipids ratio in very large VLDL | -0.10 | 3.12E-01 | -0.12 | 2.70E-01 | -0.19 | 5.91E-02 | -0.19 | 5.77E-02 | -0.21 | 3.74E-02 | 0.06 | 5.74E-01 | -0.06 | 5.51E-01 | 0.00 | 9.74E-01 | -0.15 | 1.45E-01 | 0.00 | 9.96E-01 | -0.12 | 2.29E-01 | 0.09 | 3.84E-01 |
| | | Free cholesterol to total lipids ratio in very large VLDL | -0.02 | 8.29E-01 | 0.03 | 7.52E-01 | -0.13 | 1.93E-01 | -0.26 | 9.20E-03 | -0.16 | 1.10E-01 | -0.05 | 6.58E-01 | -0.18 | 7.29E-02 | -0.21 | 4.28E-02 | -0.22 | 3.04E-02 | 0.04 | 7.22E-01 | -0.11 | 2.75E-01 | -0.01 | 9.43E-01 |
| | | TGs to total lipids ratio in very large VLDL | 0.04 | 7.05E-01 | -0.04 | 6.97E-01 | 0.15 | 1.39E-01 | 0.23 | 2.70E-02 | 0.18 | 6.34E-02 | 0.04 | 6.66E-01 | 0.17 | 9.57E-02 | 0.07 | 5.03E-01 | 0.22 | 3.44E-02 | -0.11 | 2.88E-01 | 0.18 | 7.05E-02 | -0.03 | 7.76E-01 |
| | Large VLDL ratios | Phospholipids to total lipids ratio in large VLDL | 0.00 | 9.66E-01 | 0.16 | 1.01E-01 | -0.10 | 3.26E-01 | 0.00 | 1.00E+00 | -0.16 | 8.80E-02 | -0.16 | 9.70E-02 | -0.13 | 1.85E-01 | 0.05 | 5.93E-01 | -0.16 | 1.11E-01 | 0.16 | 1.09E-01 | -0.23 | 1.41E-02 | -0.01 | 9.08E-01 |
| | | Cholesterol to total lipids ratio in large VLDL | -0.03 | 7.94E-01 | 0.07 | 4.83E-01 | -0.17 | 8.34E-02 | -0.19 | 5.11E-02 | -0.18 | 5.55E-02 | -0.15 | 1.33E-01 | -0.15 | 1.25E-01 | -0.10 | 3.29E-01 | -0.20 | 4.04E-02 | 0.03 | 7.41E-01 | -0.12 | 1.96E-01 | 0.01 | 9.40E-01 |
| | | CEs to total lipids ratio in large VLDL | -0.02 | 8.05E-01 | 0.05 | 6.46E-01 | -0.18 | 6.43E-02 | -0.26 | 8.16E-03 | -0.19 | 4.69E-02 | -0.15 | 1.21E-01 | -0.16 | 8.71E-02 | -0.17 | 7.90E-02 | -0.23 | 2.17E-02 | -0.01 | 9.02E-01 | -0.10 | 3.22E-01 | -0.02 | 8.65E-01 |
| | | Free cholesterol to total lipids ratio in large VLDL | -0.03 | 7.88E-01 | 0.10 | 3.43E-01 | -0.13 | 1.78E-01 | -0.05 | 5.85E-01 | -0.15 | 1.16E-01 | -0.11 | 2.55E-01 | -0.10 | 2.91E-01 | 0.04 | 6.82E-01 | -0.14 | 1.64E-01 | 0.10 | 3.40E-01 | -0.16 | 1.02E-01 | 0.04 | 6.53E-01 |
| | | TGs to total lipids ratio in large VLDL | 0.01 | 9.00E-01 | -0.13 | 2.12E-01 | 0.14 | 1.44E-01 | 0.11 | 2.79E-01 | 0.18 | 5.42E-02 | 0.17 | 8.70E-02 | 0.15 | 1.27E-01 | 0.03 | 7.94E-01 | 0.19 | 5.30E-02 | -0.11 | 2.97E-01 | 0.19 | 5.18E-02 | 0.00 | 9.89E-01 |
| | Medium VLDL ratios | Phospholipids to total lipids ratio in medium VLDL | 0.06 | 5.29E-01 | 0.10 | 3.24E-01 | -0.22 | 2.39E-02 | -0.25 | 9.71E-03 | -0.19 | 4.32E-02 | -0.11 | 2.48E-01 | -0.15 | 1.24E-01 | -0.11 | 2.67E-01 | -0.18 | 7.26E-02 | -0.06 | 5.81E-01 | -0.12 | 2.14E-01 | 0.02 | 8.23E-01 |
| | | Cholesterol to total lipids ratio in medium VLDL | 0.03 | 7.74E-01 | -0.02 | 8.78E-01 | -0.18 | 6.40E-02 | -0.33 | 5.33E-04 | -0.13 | 1.64E-01 | -0.10 | 3.09E-01 | -0.11 | 2.41E-01 | -0.18 | 7.08E-02 | -0.13 | 2.04E-01 | -0.11 | 2.84E-01 | 0.03 | 7.80E-01 | 0.02 | 8.70E-01 |
| | | CEs to total lipids ratio in medium VLDL | 0.01 | 8.87E-01 | -0.05 | 5.94E-01 | -0.16 | 9.43E-02 | -0.33 | 5.52E-04 | -0.12 | 2.10E-01 | -0.08 | 4.42E-01 | -0.07 | 4.81E-01 | -0.18 | 6.36E-02 | -0.08 | 4.06E-01 | -0.13 | 2.11E-01 | 0.09 | 3.30E-01 | 0.02 | 8.54E-01 |
| | | Free cholesterol to total lipids ratio in medium VLDL | 0.05 | 6.20E-01 | 0.05 | 5.93E-01 | -0.19 | 4.94E-02 | -0.30 | 1.74E-03 | -0.14 | 1.40E-01 | -0.13 | 1.75E-01 | -0.18 | 6.20E-02 | -0.15 | 1.24E-01 | -0.19 | 5.83E-02 | -0.07 | 5.17E-01 | -0.09 | 3.64E-01 | 0.01 | 9.10E-01 |
| | | TGs to total lipids ratio in medium VLDL | -0.04 | 6.67E-01 | -0.03 | 7.82E-01 | 0.20 | 3.76E-02 | 0.31 | 1.11E-03 | 0.16 | 9.19E-02 | 0.11 | 2.65E-01 | 0.13 | 1.73E-01 | 0.16 | 1.09E-01 | 0.15 | 1.28E-01 | 0.09 | 3.67E-01 | 0.03 | 7.56E-01 | -0.02 | 8.49E-01 |
| | Small VLDL ratios | Phospholipids to total lipids ratio in small VLDL | 0.02 | 8.36E-01 | 0.01 | 9.26E-01 | -0.17 | 6.99E-02 | -0.34 | 4.26E-04 | -0.14 | 1.42E-01 | -0.07 | 4.77E-01 | -0.09 | 3.66E-01 | -0.14 | 1.62E-01 | -0.09 | 3.77E-01 | -0.12 | 2.48E-01 | 0.00 | 9.69E-01 | 0.08 | 4.12E-01 |
| | | Cholesterol to total lipids ratio in small VLDL | -0.02 | 8.47E-01 | -0.03 | 7.33E-01 | -0.05 | 5.84E-01 | -0.29 | 2.48E-03 | -0.08 | 3.94E-01 | -0.05 | 6.07E-01 | -0.06 | 5.63E-01 | -0.20 | 3.96E-02 | -0.10 | 3.24E-01 | -0.20 | 4.75E-02 | 0.08 | 4.31E-01 | -0.01 | 9.25E-01 |
| | | CEs to total lipids ratio in small VLDL | -0.07 | 5.02E-01 | -0.05 | 5.90E-01 | 0.04 | 6.67E-01 | -0.19 | 5.21E-02 | -0.05 | 6.10E-01 | -0.04 | 6.81E-01 | 0.00 | 9.82E-01 | -0.20 | 4.33E-02 | -0.09 | 3.90E-01 | -0.22 | 3.33E-02 | 0.11 | 2.45E-01 | -0.09 | 3.70E-01 |
| | | Free cholesterol to total lipids ratio in small VLDL | 0.05 | 6.43E-01 | -0.01 | 9.49E-01 | -0.15 | 1.08E-01 | -0.35 | 2.57E-04 | -0.10 | 2.82E-01 | -0.05 | 5.96E-01 | -0.11 | 2.65E-01 | -0.17 | 8.43E-02 | -0.09 | 3.51E-01 | -0.15 | 1.49E-01 | 0.02 | 8.62E-01 | 0.08 | 4.31E-01 |
| | | TGs to total lipids ratio in small VLDL | 0.01 | 9.50E-01 | 0.02 | 8.34E-01 | 0.10 | 3.24E-01 | 0.31 | 1.01E-03 | 0.10 | 2.79E-01 | 0.06 | 5.49E-01 | 0.07 | 4.88E-01 | 0.18 | 5.85E-02 | 0.10 | 3.27E-01 | 0.18 | 7.76E-02 | -0.05 | 5.85E-01 | -0.02 | 8.34E-01 |
| | Very small VLDL ratios | Phospholipids to total lipids ratio in very small VLDL | -0.03 | 7.71E-01 | 0.13 | 1.83E-01 | 0.18 | 5.59E-02 | 0.13 | 1.94E-01 | 0.06 | 5.39E-01 | -0.11 | 2.46E-01 | 0.05 | 5.93E-01 | 0.05 | 6.22E-01 | 0.07 | 4.93E-01 | 0.02 | 8.74E-01 | -0.03 | 7.32E-01 | -0.20 | 3.69E-02 |
| | | Cholesterol to total lipids ratio in very small VLDL | 0.08 | 4.17E-01 | -0.10 | 3.25E-01 | 0.01 | 8.95E-01 | -0.19 | 5.49E-02 | 0.04 | 6.47E-01 | 0.10 | 3.16E-01 | -0.01 | 9.15E-01 | -0.17 | 8.17E-02 | -0.01 | 9.59E-01 | -0.19 | 6.14E-02 | 0.17 | 8.28E-02 | 0.02 | 8.49E-01 |
| | | CEs to total lipids ratio in very small VLDL | 0.08 | 4.42E-01 | -0.11 | 2.67E-01 | 0.01 | 9.29E-01 | -0.18 | 6.45E-02 | 0.04 | 6.79E-01 | 0.10 | 3.00E-01 | 0.00 | 9.83E-01 | -0.16 | 9.84E-02 | 0.00 | 9.88E-01 | -0.18 | 7.12E-02 | 0.17 | 7.53E-02 | 0.03 | 7.88E-01 |
| | | Free cholesterol to total lipids ratio in very small VLDL | 0.09 | 3.67E-01 | 0.08 | 4.16E-01 | 0.07 | 4.57E-01 | -0.24 | 1.25E-02 | 0.10 | 3.12E-01 | 0.03 | 7.49E-01 | -0.10 | 3.05E-01 | -0.26 | 8.25E-03 | -0.05 | 5.90E-01 | -0.22 | 2.67E-02 | 0.11 | 2.55E-01 | -0.09 | 3.72E-01 |
| | | TGs to total lipids ratio in very small VLDL | -0.08 | 3.96E-01 | 0.08 | 4.19E-01 | -0.06 | 5.19E-01 | 0.18 | 6.01E-02 | -0.07 | 4.86E-01 | -0.09 | 3.83E-01 | 0.00 | 9.85E-01 | 0.18 | 5.98E-02 | -0.01 | 9.04E-01 | 0.21 | 3.57E-02 | -0.19 | 5.16E-02 | 0.03 | 7.71E-01 |
| | IDL ratios | Phospholipids to total lipids ratio in IDL | -0.08 | 4.27E-01 | 0.13 | 1.90E-01 | 0.13 | 1.65E-01 | 0.17 | 9.06E-02 | 0.01 | 9.09E-01 | -0.09 | 3.69E-01 | 0.15 | 1.13E-01 | 0.02 | 8.02E-01 | 0.10 | 3.03E-01 | 0.04 | 6.80E-01 | 0.06 | 5.24E-01 | -0.15 | 1.39E-01 |
| | | Cholesterol to total lipids ratio in IDL | 0.10 | 3.03E-01 | -0.12 | 2.54E-01 | -0.06 | 5.52E-01 | -0.16 | 1.08E-01 | 0.01 | 8.93E-01 | 0.11 | 2.67E-01 | -0.05 | 6.38E-01 | -0.13 | 1.99E-01 | -0.03 | 7.81E-01 | -0.16 | 1.11E-01 | 0.09 | 3.57E-01 | 0.03 | 7.38E-01 |
| | | CEs to total lipids ratio in IDL | 0.10 | 2.90E-01 | -0.13 | 1.92E-01 | -0.07 | 4.49E-01 | -0.15 | 1.19E-01 | -0.02 | 8.50E-01 | 0.11 | 2.41E-01 | -0.06 | 5.40E-01 | -0.11 | 2.52E-01 | -0.04 | 6.82E-01 | -0.15 | 1.45E-01 | 0.06 | 5.32E-01 | 0.04 | 6.53E-01 |
| | | Free cholesterol to total lipids ratio in IDL | 0.04 | 6.75E-01 | 0.01 | 9.01E-01 | 0.06 | 5.33E-01 | -0.14 | 1.63E-01 | 0.19 | 4.45E-02 | 0.04 | 6.94E-01 | 0.04 | 6.43E-01 | -0.16 | 1.01E-01 | 0.05 | 6.28E-01 | -0.20 | 4.87E-02 | 0.21 | 2.94E-02 | -0.05 | 6.04E-01 |
| | | TGs to total lipids ratio in IDL | -0.10 | 3.28E-01 | 0.10 | 3.20E-01 | 0.02 | 8.28E-01 | 0.14 | 1.42E-01 | -0.02 | 8.14E-01 | -0.11 | 2.76E-01 | 0.00 | 9.69E-01 | 0.15 | 1.35E-01 | 0.00 | 9.94E-01 | 0.19 | 6.32E-02 | -0.13 | 1.69E-01 | 0.01 | 9.48E-01 |
| | Large LDL ratios | Phospholipids to total lipids ratio in large LDL | 0.02 | 8.57E-01 | -0.04 | 6.77E-01 | 0.16 | 9.28E-02 | 0.11 | 2.54E-01 | 0.12 | 2.33E-01 | 0.04 | 6.75E-01 | -0.04 | 6.97E-01 | 0.00 | 9.85E-01 | 0.01 | 8.81E-01 | -0.03 | 7.40E-01 | 0.02 | 8.08E-01 | -0.16 | 1.01E-01 |
| | | CEs to total lipids ratio in large LDL | -0.05 | 5.80E-01 | 0.06 | 5.80E-01 | 0.12 | 2.32E-01 | 0.16 | 1.02E-01 | 0.09 | 3.77E-01 | -0.08 | 4.07E-01 | 0.02 | 8.06E-01 | -0.05 | 5.80E-01 | -0.03 | 7.30E-01 | 0.00 | 1.00E+00 | -0.12 | 1.98E-01 | 0.12 | 2.37E-01 |
| | Medium LDL ratios | CEs to total lipids ratio in medium LDL | -0.02 | 8.29E-01 | -0.16 | 1.19E-01 | 0.18 | 6.84E-02 | 0.13 | 1.85E-01 | 0.12 | 2.22E-01 | 0.06 | 5.13E-01 | 0.06 | 5.68E-01 | 0.12 | 2.30E-01 | 0.00 | 9.61E-01 | 0.14 | 1.59E-01 | 0.10 | 3.21E-01 | -0.06 | 5.56E-01 |
| | | Cholesterol to total lipids ratio in large LDL | 0.04 | 6.76E-01 | 0.04 | 6.88E-01 | -0.15 | 1.17E-01 | -0.17 | 8.19E-02 | -0.11 | 2.76E-01 | 0.00 | 9.87E-01 | -0.04 | 6.74E-01 | -0.08 | 4.01E-01 | 0.00 | 9.82E-01 | -0.07 | 4.79E-01 | 0.02 | 8.67E-01 | 0.13 | 2.00E-01 |
| | | Cholesterol to total lipids ratio in medium LDL | 0.00 | 9.80E-01 | 0.03 | 7.61E-01 | -0.19 | 4.31E-02 | -0.14 | 1.64E-01 | -0.15 | 1.23E-01 | -0.02 | 8.16E-01 | 0.05 | 6.34E-01 | 0.14 | 1.42E-01 | -0.01 | 8.85E-01 | 0.17 | 9.43E-02 | -0.06 | 5.15E-01 | -0.04 | 6.55E-01 |
| | | Free cholesterol to total lipids ratio in large LDL | 0.01 | 8.94E-01 | -0.04 | 6.60E-01 | -0.16 | 8.65E-02 | -0.16 | 1.10E-01 | -0.11 | 2.35E-01 | 0.03 | 7.94E-01 | 0.04 | 6.44E-01 | 0.00 | 9.96E-01 | -0.07 | 4.60E-01 | 0.10 | 3.08E-01 | -0.19 | 4.31E-02 | 0.10 | 3.19E-01 |
| | | Free cholesterol to total lipids ratio in medium LDL | 0.04 | 6.78E-01 | -0.14 | 1.53E-01 | -0.04 | 6.97E-01 | -0.06 | 5.46E-01 | 0.02 | 8.72E-01 | 0.15 | 1.32E-01 | 0.03 | 7.60E-01 | -0.04 | 6.63E-01 | 0.00 | 9.98E-01 | -0.01 | 9.29E-01 | 0.01 | 8.81E-01 | 0.14 | 1.60E-01 |
| | | Phospholipids to total lipids ratio in medium LDL | 0.05 | 6.30E-01 | 0.16 | 1.16E-01 | -0.21 | 2.65E-02 | -0.14 | 1.57E-01 | -0.18 | 5.42E-02 | -0.08 | 4.17E-01 | -0.09 | 3.51E-01 | -0.02 | 8.78E-01 | 0.06 | 5.31E-01 | -0.12 | 2.54E-01 | 0.14 | 1.37E-01 | -0.13 | 1.70E-01 |
| | | TGs to total lipids ratio in large LDL | -0.05 | 6.08E-01 | 0.08 | 4.51E-01 | 0.15 | 1.23E-01 | 0.15 | 1.21E-01 | 0.10 | 3.09E-01 | -0.09 | 3.60E-01 | -0.05 | 6.27E-01 | -0.12 | 2.30E-01 | -0.01 | 9.38E-01 | -0.20 | 4.44E-02 | 0.02 | 8.61E-01 | -0.04 | 7.13E-01 |
| | | TGs to total lipids ratio in medium LDL | 0.00 | 9.75E-01 | -0.20 | 4.17E-02 | 0.05 | 6.10E-01 | 0.03 | 7.65E-01 | 0.10 | 3.08E-01 | 0.17 | 8.66E-02 | 0.07 | 4.81E-01 | -0.09 | 3.86E-01 | 0.05 | 6.10E-01 | -0.23 | 2.53E-02 | 0.07 | 4.51E-01 | -0.02 | 8.00E-01 |
| | Small LDL ratios | Phospholipids to total lipids ratio in small LDL | -0.02 | 8.23E-01 | -0.10 | 3.37E-01 | 0.16 | 9.19E-02 | 0.16 | 9.33E-02 | 0.09 | 3.66E-01 | 0.09 | 3.41E-01 | 0.08 | 3.90E-01 | -0.04 | 6.58E-01 | 0.06 | 5.34E-01 | -0.11 | 2.77E-01 | 0.23 | 1.62E-02 | -0.14 | 1.68E-01 |
| | | Cholesterol to total lipids ratio in small LDL | 0.02 | 8.30E-01 | 0.01 | 8.94E-01 | -0.14 | 1.35E-01 | -0.18 | 5.96E-02 | -0.08 | 4.31E-01 | 0.00 | 9.70E-01 | -0.05 | 5.73E-01 | 0.00 | 9.63E-01 | -0.04 | 6.93E-01 | 0.01 | 9.01E-01 | -0.19 | 5.38E-02 | 0.14 | 1.46E-01 |
| | | CEs to total lipids ratio in small LDL | 0.05 | 6.31E-01 | 0.11 | 2.73E-01 | -0.21 | 2.95E-02 | -0.21 | 3.20E-02 | -0.13 | 1.91E-01 | -0.10 | 2.85E-01 | -0.11 | 2.40E-01 | -0.01 | 8.94E-01 | -0.08 | 4.31E-01 | 0.09 | 3.94E-01 | -0.22 | 2.32E-02 | 0.11 | 2.72E-01 |
| | | Free cholesterol to total lipids ratio in small LDL | -0.04 | 6.87E-01 | -0.22 | 3.10E-02 | 0.04 | 6.71E-01 | 0.12 | 2.23E-01 | 0.05 | 5.83E-01 | 0.26 | 6.96E-03 | 0.09 | 3.68E-01 | 0.04 | 7.17E-01 | 0.06 | 5.72E-01 | -0.19 | 5.56E-02 | -0.05 | 5.98E-01 | 0.03 | 7.30E-01 |
| | | TGs to total lipids ratio in small LDL | 0.00 | 9.60E-01 | 0.16 | 1.04E-01 | -0.07 | 4.88E-01 | 0.02 | 8.59E-01 | -0.04 | 6.88E-01 | -0.21 | 3.17E-02 | -0.12 | 2.12E-01 | 0.08 | 3.90E-01 | -0.10 | 3.13E-01 | 0.22 | 2.69E-02 | -0.18 | 6.32E-02 | 0.01 | 9.50E-01 |
| | Very large HDL ratios | Phospholipids to total lipids ratio in very large HDL | 0.01 | 9.26E-01 | -0.04 | 6.86E-01 | 0.17 | 7.16E-02 | -0.09 | 3.46E-01 | 0.16 | 8.66E-02 | 0.00 | 9.76E-01 | -0.07 | 4.82E-01 | -0.04 | 7.15E-01 | -0.05 | 6.11E-01 | -0.08 | 4.06E-01 | 0.11 | 2.71E-01 | 0.00 | 9.69E-01 |
| | | Cholesterol to total lipids ratio in very large HDL | -0.04 | 7.07E-01 | -0.03 | 7.81E-01 | -0.08 | 3.86E-01 | 0.20 | 4.36E-02 | -0.10 | 3.17E-01 | 0.16 | 9.61E-02 | 0.20 | 3.64E-02 | 0.14 | 1.43E-01 | 0.17 | 8.96E-02 | -0.06 | 5.53E-01 | 0.00 | 9.99E-01 | 0.00 | 9.79E-01 |
| | | CEs to total lipids ratio in very large HDL | 0.00 | 9.93E-01 | -0.11 | 2.89E-01 | -0.22 | 2.41E-02 | -0.12 | 2.02E-01 | 0.03 | 7.61E-01 | 0.05 | 6.09E-01 | 0.01 | 9.44E-01 | 0.04 | 7.18E-01 | 0.06 | 5.37E-01 | -0.13 | 2.01E-01 | 0.12 | 2.28E-01 | 0.13 | 1.87E-01 |
| | | Free cholesterol to total lipids ratio in very large HDL | -0.02 | 8.02E-01 | 0.06 | 5.33E-01 | 0.09 | 3.56E-01 | 0.19 | 4.82E-02 | -0.09 | 3.58E-01 | 0.05 | 5.88E-01 | 0.14 | 1.61E-01 | 0.05 | 5.97E-01 | 0.08 | 4.32E-01 | 0.06 | 5.56E-01 | -0.08 | 4.16E-01 | -0.09 | 3.44E-01 |
| | | TGs to total lipids ratio in very large HDL | 0.02 | 8.13E-01 | 0.07 | 4.80E-01 | -0.20 | 3.90E-02 | 0.00 | 9.84E-01 | -0.17 | 8.16E-02 | -0.11 | 2.79E-01 | -0.12 | 2.17E-01 | -0.04 | 6.72E-01 | -0.12 | 2.25E-01 | 0.15 | 1.36E-01 | -0.18 | 6.38E-02 | 0.00 | 1.00E+00 |
| | Large HDL ratios | Phospholipids to total lipids ratio in large HDL | 0.00 | 9.88E-01 | 0.01 | 8.90E-01 | 0.07 | 4.60E-01 | 0.15 | 1.37E-01 | -0.09 | 3.37E-01 | 0.03 | 7.34E-01 | 0.06 | 5.29E-01 | 0.24 | 1.14E-02 | 0.07 | 4.98E-01 | 0.07 | 5.12E-01 | -0.13 | 1.86E-01 | 0.04 | 6.60E-01 |
| | | Cholesterol to total lipids ratio in large HDL | -0.02 | 8.65E-01 | -0.05 | 6.54E-01 | 0.00 | 9.75E-01 | -0.10 | 3.08E-01 | 0.13 | 1.92E-01 | 0.02 | 8.30E-01 | 0.01 | 9.43E-01 | -0.05 | 5.94E-01 | 0.02 | 8.57E-01 | -0.16 | 1.08E-01 | 0.16 | 1.04E-01 | 0.00 | 9.74E-01 |
| | | CEs to total lipids ratio in large HDL | -0.02 | 8.74E-01 | -0.05 | 6.53E-01 | 0.00 | 9.69E-01 | -0.08 | 4.04E-01 | 0.14 | 1.40E-01 | 0.03 | 7.43E-01 | 0.02 | 8.06E-01 | -0.04 | 6.94E-01 | 0.05 | 6.41E-01 | -0.16 | 1.07E-01 | 0.15 | 1.08E-01 | -0.01 | 9.50E-01 |
| | | Free cholesterol to total lipids ratio in large HDL | -0.02 | 8.41E-01 | -0.04 | 6.62E-01 | 0.00 | 9.92E-01 | -0.15 | 1.32E-01 | 0.08 | 3.95E-01 | -0.01 | 8.87E-01 | -0.03 | 7.63E-01 | -0.09 | 3.65E-01 | -0.05 | 6.42E-01 | -0.15 | 1.38E-01 | 0.15 | 1.24E-01 | 0.01 | 9.44E-01 |
| | | TGs to total lipids ratio in large HDL | 0.03 | 7.26E-01 | 0.04 | 6.99E-01 | -0.09 | 3.70E-01 | 0.03 | 7.53E-01 | -0.14 | 1.40E-01 | -0.04 | 7.05E-01 | -0.09 | 3.28E-01 | -0.07 | 4.93E-01 | -0.14 | 1.71E-01 | 0.13 | 2.14E-01 | -0.16 | 9.24E-02 | -0.02 | 8.52E-01 |
| | Medium HDL ratios | Phospholipids to total lipids ratio in medium HDL | -0.03 | 7.28E-01 | 0.17 | 8.49E-02 | 0.21 | 2.77E-02 | 0.12 | 2.02E-01 | -0.01 | 9.33E-01 | -0.10 | 3.13E-01 | 0.00 | 9.92E-01 | -0.03 | 7.93E-01 | -0.01 | 9.31E-01 | 0.08 | 4.56E-01 | -0.05 | 5.79E-01 | -0.21 | 3.59E-02 |
| | | Cholesterol to total lipids ratio in medium HDL | 0.02 | 8.22E-01 | -0.19 | 5.42E-02 | -0.09 | 3.57E-01 | -0.04 | 6.71E-01 | 0.09 | 3.63E-01 | 0.20 | 3.86E-02 | 0.11 | 2.68E-01 | 0.03 | 7.76E-01 | 0.12 | 2.28E-01 | -0.17 | 9.61E-02 | 0.18 | 6.18E-02 | 0.13 | 1.90E-01 |
| | | CEs to total lipids ratio in medium HDL | -0.01 | 9.43E-01 | -0.18 | 6.82E-02 | 0.03 | 7.88E-01 | 0.04 | 6.86E-01 | 0.17 | 7.16E-02 | 0.22 | 2.14E-02 | 0.19 | 5.29E-02 | 0.05 | 5.77E-01 | 0.21 | 3.08E-02 | -0.21 | 4.26E-02 | 0.22 | 2.42E-02 | 0.06 | 5.27E-01 |
| | | Free cholesterol to total lipids ratio in medium HDL | 0.05 | 6.26E-01 | -0.15 | 1.49E-01 | -0.20 | 4.15E-02 | -0.16 | 1.08E-01 | -0.05 | 6.15E-01 | 0.09 | 3.71E-01 | -0.03 | 7.76E-01 | -0.03 | 7.66E-01 | -0.04 | 7.03E-01 | -0.03 | 7.34E-01 | 0.07 | 4.96E-01 | 0.19 | 5.58E-02 |
| | | TGs to total lipids ratio in medium HDL | 0.00 | 9.61E-01 | 0.19 | 6.51E-02 | -0.05 | 6.37E-01 | -0.04 | 7.16E-01 | -0.14 | 1.50E-01 | -0.25 | 9.30E-03 | -0.18 | 5.92E-02 | -0.03 | 7.63E-01 | -0.19 | 5.56E-02 | 0.21 | 3.92E-02 | -0.25 | 8.60E-03 | -0.05 | 6.21E-01 |
| | Small HDL ratios | Phospholipids to total lipids ratio in small HDL | 0.09 | 3.52E-01 | -0.05 | 6.24E-01 | 0.12 | 2.18E-01 | -0.03 | 7.41E-01 | 0.04 | 6.72E-01 | 0.14 | 1.40E-01 | -0.14 | 1.54E-01 | -0.14 | 1.67E-01 | -0.07 | 4.65E-01 | -0.03 | 7.76E-01 | 0.05 | 6.13E-01 | -0.09 | 3.57E-01 |
| | | Cholesterol to total lipids ratio in small HDL | -0.04 | 6.66E-01 | -0.17 | 9.27E-02 | -0.09 | 3.70E-01 | 0.04 | 6.67E-01 | 0.07 | 4.72E-01 | 0.21 | 3.07E-02 | 0.27 | 4.57E-03 | 0.12 | 2.38E-01 | 0.23 | 1.71E-02 | -0.19 | 5.60E-02 | 0.20 | 3.63E-02 | 0.16 | 1.09E-01 |
| | | CEs to total lipids ratio in small HDL | -0.02 | 8.60E-01 | -0.13 | 1.99E-01 | 0.01 | 9.51E-01 | 0.16 | 1.03E-01 | 0.07 | 4.78E-01 | 0.25 | 9.72E-03 | 0.29 | 2.03E-03 | 0.13 | 1.79E-01 | 0.27 | 5.50E-03 | -0.17 | 9.74E-02 | 0.17 | 7.47E-02 | 0.05 | 6.40E-01 |
| | | Free cholesterol to total lipids ratio in small HDL | -0.04 | 7.04E-01 | -0.04 | 6.59E-01 | -0.15 | 1.08E-01 | -0.27 | 4.61E-03 | -0.02 | 8.72E-01 | -0.16 | 1.08E-01 | -0.13 | 1.82E-01 | -0.07 | 4.51E-01 | -0.14 | 1.56E-01 | 0.00 | 9.89E-01 | -0.01 | 9.07E-01 | 0.18 | 6.06E-02 |
| | | TGs to total lipids ratio in small HDL | -0.04 | 7.24E-01 | 0.18 | 6.96E-02 | -0.02 | 8.03E-01 | -0.01 | 9.18E-01 | -0.11 | 2.61E-01 | -0.28 | 3.25E-03 | -0.14 | 1.49E-01 | 0.00 | 9.83E-01 | -0.16 | 1.04E-01 | 0.20 | 4.88E-02 | -0.23 | 1.51E-02 | -0.06 | 5.23E-01 |
